# B-Cell Activation Reflects Psychiatric Symptom Severity Amid Multi-Lineage Peripheral Immune Activation in Pediatric Acute-Onset Neuropsychiatric Syndrome

**DOI:** 10.64898/2026.11.24.26368791

**Authors:** Samantha Leonard, Imran Khan Mohammad, Natalia Sigal, Noelle Schlenk, Bahare Farhadian, Mei Ma, Melissa Silverman, Margo Thienemann, Paula Tran, Yuhuan Xie, Sana Ahmed, Theresa Willett, Joseph D Hernandez, Tanya Saini, Avis Chan, Laurie Columbo, Jacquelyn Horgan, M Karen Newell-Rogers, Catherine A Blish, David B Lewis, Holden Maecker, Jennifer Frankovich

**Author notes:** Shared first or shared last authors (Samantha Leonard, Imran Khan Mohammad, Holden Maecker, Jennifer Frankovich).

## Abstract

**Importance:** Pediatric acute-onset neuropsychiatric syndrome (PANS) lacks validated disease-activity biomarkers. Relationships between peripheral immune-cell activation and psychiatric severity remain incompletely characterized.

**Objective:** To compare peripheral immune-cell phenotypes during PANS flare and recovery with those of healthy controls and evaluate associations with psychiatric severity and functional impairment.

**Design, Setting, and Participants:** In this prospective single-center cohort study, peripheral blood and psychiatric symptom ratings were collected from 20 patients meeting PANS criteria and 20 matched healthy controls at Stanford University Immune Behavioral Health Program between November 2016 to January 2020.

**Main Predictors:** Clinical group (PANS flare, PANS recovery, or healthy control) and peripheral B-cell, CD4+ T-cell, monocyte, and NK-cell ratios.

**Main Outcomes and Measures:** Mass cytometry quantified ratios of activated (CD19+CD20+CD69+) to naive (CD19+CD20+IgD+CD27−) B-cells; activated (CD3^+^CD4^+^CD69^+^) to naive (CD3^+^CD4^+^CD45RA^+^CD27^+^) T-cells; classical (CD14+CD16−CCR2+) to nonclassical/patrolling (CD14+CD16+CX3CR1+) monocytes; and activated (CD56-bright CD69+) to resting (CD56-dim) NK cells. Primary clinical measures were the clinician-rated Children’s Global Assessment Scale (CGAS), parent-rated PANS Global Impairment Scale, and PANS-31 Symptom Rating Scale (30 available items). Linear mixed-effects models accounted for repeated measurements; false discovery rate (FDR) correction addressed multiple testing.

**Results:** Analyses included 19 flare samples, 15 recovery samples, and 20 healthy control samples. B-cell, CD4+ T-cell, monocyte, and NK-cell activation ratios were higher during flare than in healthy controls (all *P* < .01) or recovery (all *P* < .03). Recovery and control ratios did not differ significantly. Each one unit increase in the B-cell activation ratio was associated with lower CGAS scores, indicating worse functioning (β = −14.89; FDR-adjusted *P* < .001), higher Global Impairment scores (β = 12.81; FDR-adjusted *P* = .004), and higher PANS-31 scores (β = 8.37; FDR-adjusted *P* = .012). CD4+ T-cell, monocyte, and NK-cell activation ratios were not significantly associated with psychiatric scores. Adjustment for sex or immunomodulatory medication exposure did not materially alter B-cell associations. Adjustment for disease state attenuated these associations, although the CGAS association remained significant.

**Conclusions and Relevance:** In this longitudinal observational study, PANS flares were associated with higher peripheral immune-cell activation ratios across all cell subsets studies (B, CD4+ T, monocyte, and NK). Only B-cell activation ratios were associated with greater psychiatric severity and impairment. The activated-to-naive B-cell ratio warrants independent validation as a candidate disease-activity marker for the psychiatric aspects of PANS; these findings do not establish causality or clinical utility.

## Introduction

Pediatric Acute-Onset Neuropsychiatric Syndrome (PANS) is defined by abrupt-onset obsessive-compulsive symptoms and/or severe eating restriction, accompanied by at least two additional neuropsychiatric symptoms^1,2^). Pediatric Autoimmune Neuropsychiatric Disorders Associated with Streptococcal infections (PANDAS) represent the subset associated with confirmed preceding Group A streptococcal (GAS) infection^2^. PANS/PANDAS often follows a relapsing–remitting course, but its pathophysiology remains incompletely understood, and validated biomarkers of disease activity are lacking.

Sydenham chorea (SC), the neurological manifestation of acute rheumatic fever (ARF), provides a clinical framework for considering post-streptococcal neuropsychiatric disease. ARF is a prototypic post-streptococcal inflammatory disease whose manifestations include chorea, arthritis, carditis and skin involvement^3,4^. Delayed presentation can impede confirmation of a preceding GAS infection in SC; a challenge also encountered in PANDAS. Emotional lability, anxiety, irritability and obsessive–compulsive symptoms are frequent in SC and overlap with the psychiatric features of PANS/PANDAS ^5–7^. Neuropsychiatric symptoms have also been reported in patients with ARF without chorea^8^, These parallels motivate investigation of both neural and systemic immune responses in PANS/PANDAS.

Postinfectious neuroinflammation and autoimmunity are proposed mechanisms in PANDAS and potentially a subset of the etiologically heterogeneous PANS population^9,10^. Studies of patient-derived antibodies support streptococcal–neuronal antigen cross-reactivity and altered neuronal signalling^11–16^. Complementary rodent studies demonstrate brain-reactive antibodies and behavioral and motor abnormalities following streptococcal-antigen immunization or passive antibody transfer ^14,17–19^. However, antibody findings vary across cohorts and assays; Dale and colleagues detected surface dopamine D2 receptor antibodies in SC and basal ganglia encephalitis, but not in their PANDAS cohort^20^. Collectively, these studies support biological plausibility rather than a shared causal mechanism, and neuronal binding or functional effects alone do not establish streptococcal molecular mimicry^21–24^.

Convergent evidence implicates basal ganglia circuitry in PANS/PANDAS. Neuroimaging studies report basal ganglia enlargement in acute presentations, positron emission tomography (PET) findings consistent with microglial activation, and microstructural changes in the basal ganglia and other subcortical regions^25–28^, alongside neurological soft signs associated with basal ganglia dysfunction^29^. Polysomnography studies report excessive motor activity and reduced atonia during REM sleep reminiscent of the during rapid eye movement (REM) sleep motor disinhibition seen in Parkinson disease, another disorder of basal ganglia–brainstem motor circuitry^30–33^. Patient-derived antibodies also bind striatal cholinergic interneurons and alter their activity^23,24,34^. In mice, repeated intranasal GAS infection induces Th17-cell infiltration into the central nervous system, blood–brain barrier disruption, microglial activation and synaptic loss^35,36^.

Evidence of systemic immune involvement includes elevated circulating inflammatory mediators during acute PANS/PANDAS illness^37^, including TNF, GM-CSF and CXCL10, which are also implicated in ARF^38–40^. Reported rheumatological findings include enthesitis, subclinical joint inflammation on imaging and subtle signs of vasculopathy^41,42^. These features resemble the articular, subclinical cardiac and vascular involvement of ARF, but the pathology is not identical^3,43^. In observational studies, early anti-inflammatory treatment with non-steroidal anti-inflammatory drugs (NSAIDs) and corticosteroids has been associated with shorter PANS/PANDAS flares-indirectly supporting an inflammatory contribution but not establishing efficacy^44,45^. Similarly, ARF arthritis responds rapidly to NSAIDS^46,47^, and corticosteroids are associated with shorter SC flare duration^7,48,49^. Peripheral acute-phase reactants do not reliably track neuropsychiatric involvement in SC^49^. Nonetheless, immunotherapy is recommended as first-line treatment for ARF and moderate to severe SC^7^, whereas its role in PANS/PANDAS remains controversial. In both PANS/PANDAS and SC, neuroinflammation may occur without abnormal routine peripheral markers of systemic inflammation, which shows the need for more sensitive measures of peripheral immune activity.

Autoantibody targets in PANS/PANDAS also extend beyond neuronal surface antigens to include tubulin^16^ and antigens linked to systemic and organ-specific autoimmune diseases^50^. A subset of patients harbors autoantibodies against interferon-λ, a key mediator of host defense at barrier surfaces, that neutralizes its signaling in vitro^50^. This breadth echoes the heterogeneous autoantigen reactivity and epitope spreading described in ARF^51,52^. Broader immune dysregulation may also help explain why arthritis and other autoimmune diseases are more common in this population ^41,42^. Whereas ESR and CRP are typically elevated in ARF ^43^, PANS/PANDAS has no reliable peripheral marker of inflammation. Defining the peripheral immune response could therefore yield more sensitive biomarkers of systemic inflammation and disease activity and clarify their relationship to neuropsychiatric symptoms.

Recent immunophenotyping studies in PANS identify disease-state-dependent changes in circulating myeloid cells. Rahman et al. (2025) reported increased M1-like monocytes during flare and increased M2-like populations during recovery, together with expansion of CD11c-positive migratory monocytes in males and monocyte-derived dendritic cells in both sexes during flare.^53^ PANS flare plasma also induced the CNS-trafficking-associated markers VLA-4 and CD166 on healthy donor monocytes, suggesting that circulating factors can modulate leukocyte trafficking phenotypes. Circulating monocyte activation has similarly been described in ARF^40^. In an open-label PANS study, intravenous immunoglobulin (IVIG) treatment was associated with reductions in pro-inflammatory monocyte and dendritic cell populations alongside neuropsychiatric improvement.^54^ Whether coordinated disease-state-dependent activation extends across innate and adaptive immune populations in PANS, and how these phenotypes relate to psychiatric symptom profiles and severity, remains unresolved. Defining these relationships could inform peripheral biomarkers of disease activity.

This study used mass cytometry to characterize peripheral immune-cell activation in a longitudinal PANS cohort during flare and recovery, alongside age-matched healthy controls. Ratios of activated B-cell, CD4+ T-cells, natural killer-cell and monocyte subsets to their corresponding naïve, inactive or patrolling populations were examined across disease states and in relation to neuropsychiatric symptom severity. The primary hypothesis was that greater immune-cell activation would accompany active disease and greater symptom severity.

## Materials and Methods

### Study population

This study was conducted at the Human Immune Monitoring Center (HIMC) at Stanford University. Samples were collected in the Stanford Children’s, Immune Behavioral Health (IBH) Clinic, a multidisciplinary specialty clinic providing psychiatric, psychological, pediatric, rheumatological, and immunological care for patients with PANS since 2012. Prior to IBH clinic admission, patients were screened for a symptom presentation aligning with PANS through clinician review of medical records, consultation with referring providers, and, more recently, parent-completed screening questionnaires (see Supplementary Methods for screening protocol).

PANS diagnoses were established by the Stanford IBH medical-psychiatric team using evaluation criteria established at the 2013 PANS Consensus Conference^75^. Disease state definitions followed Masterson et al. (2025)^76^. Flare was defined as a period of escalating neuropsychiatric symptoms with associated functional impairment relative to the patient’s baseline. Minor symptom escalations without substantial functional impairment were not classified as flares, and worsening of pain, fatigue, or arthritis without neuropsychiatric deterioration was excluded. Recovery was defined as a significant improvement in symptoms and functioning compared with the peak of the most recent flare. It was classified as partial if impairment remained above baseline, or full if impairment returned to or near baseline Additional details regarding cohort characterization and disease-state classification have been published previously^76^.

This study was approved by the Stanford Institutional Review Board under the Stanford Human Research Protection Program (protocol 26922). Informed consent was obtained from parents or legal guardians. Child assent was obtained when appropriate. The study was conducted in accordance with the principles of the Declaration of Helsinki.

### Sample collection, processing, and storage

Whole blood samples were prospectively collected from patients with PANS and healthy pediatric controls as part of the ongoing Stanford IBH/PANS research study. Peripheral venous blood (from both patients and controls) was drawn into green top tubes at scheduled visits and processed within 1 hour of collection. fresh whole blood was mixed with Proteomic Stabilizer PROT1 (Smart Tube Inc., San Carlos, CA) at a 1:1.4 blood-to-PROT1 ratio, incubated for 10 minutes at room temperature, and stored at −80 °C. All specimens were processed by the Stanford Clinical Translational Research Unit and maintained by the Stanford Medicine Biobank under standardized chain-of-custody conditions, and the time from collection, to fixation, to cryopreservation was recorded for each sample.

For the present study consecutive samples were chosen by AC among patients who were classified as meeting PANS criteria and satisfied the following criteria: clear flare state and flare trajectory data (documented in clinical records and/or clinician encounter forms), psychometric scores, and available matched control. To begin to explore the heterogeneity of flare-state, AC was directed to select 10 flare samples obtained during an improving trajectory and 10 samples obtained during a worsening trajectory or at the peak of flare symptoms. Recovery samples were available for 15/20 patients. Healthy controls were recruited from the same community as patients, were processed and stored using the same pipeline as patients, and matched forage (± 1 year), sex, and race/ethnicity. Median storage time was 15.5 months (range, 0.4–37.8) overall: 4.1 months (range, 0.5–25.2) for healthy controls, 9.2 months (range, 1.5–19.4) for recovery samples, and 20.6 months (range, 0.4–37.8) for flare samples.

### Mass cytometry

A 37-parameter panel was designed to resolve activation and differentiation states across B-cell, CD4 T-cell, NK and monocyte cell lineages. Antibodies were purchased pre-conjugated from Standard BioTools (South San Francisco, CA) or conjugated in-house using the Maxpar X8 Antibody Labeling Kit per manufacturer instructions. Conjugate yield was determined by absorbance at 280 nm, and conjugates were stabilized in Candor PBS-based antibody stabilization buffer and stored at 4°C. Each conjugate was titrated on healthy donor PBMC before inclusion. Clone, isotope, vendor, and final concentration for all antibodies are listed in Supplementary Table S2. Lineage and activation markers included in the panel were previously established by the Stanford HIMC^55^ and included: CD3, CD4, CD8, CD45RA, CD27, CD69, CD19, CD20, IgD, CD14, CD16, CCR2, CX3CR1, CD56, and HLA-DR. Intracellular signaling was assessed using eight phospho-specific antibodies: pSTAT1, pSTAT3, pSTAT4, pSTAT5, pERK1/2, pp38, pCREB, and pS6.

Samples were thawed once and analyzed in 3 batches in April 2020. Samples were assigned to assay batches so that each batch contained a balanced number of patients and healthy controls. Samples were thawed then washed twice with Smart Tube Thaw-Lyse buffer and once with CyFACS buffer (PBS supplemented with 2% BSA, 2 mM EDTA, and 0.1% sodium azide). Cells were stained with 20 µL of surface antibody cocktail for 30 minutes at room temperature, washed twice with CyFACS, permeabilized with 100% methanol, and stored at −80 °C overnight. The following day, cells were washed with CyFACS, stained with 20 µL of intracellular antibody cocktail for 30 minutes at room temperature, and washed twice with CyFACS. Cells were then incubated in 100 µL of iridium DNA intercalator (1:2,000 in 2% PFA in PBS) for 20 minutes at room temperature, washed once with CyFACS and twice with MilliQ water, and resuspended at 7.5 × 10□ cells/mL in MilliQ water containing a 1:10 dilution of EQ Four Element Calibration Beads.

Samples were acquired on a Helios mass cytometer at the Stanford Human Immune Monitoring Center at an event rate not exceeding 300 events per second, with a minimum of 200,000 events collected per sample. A shared reference control was acquired with each batch to assess inter-batch technical variability. One patient sample was excluded because of insufficient T-cell recovery (fewer than the prespecified minimum of 200,000 events). All analyses, including phospho-signaling, were performed on unstimulated samples under basal conditions. Data were analyzed in Cytobank. Calibration beads were excluded, and intact single cells were identified using Gaussian discrimination parameters and the iridium DNA intercalator (191Ir/193Ir). CD45+ leukocytes were then selected, CD66b+ granulocytes were excluded, and cell subsets were identified by lineage-specific gating (Supplementary Figure 3; Fernandez and Maecker, 2015).^56^.

### Data processing and gating

Data processing and analyses were performed from December 2025 to September 2026. Raw .fcs files were normalized to EQ bead signal using the Standard BioTools normalization algorithm. Normalized files were analyzed in Cytobank. Sequential manual gating identified intact, singlet leukocytes: Gaussian discrimination parameters, followed by DNA-positive 191Ir/193Ir events, and CD45-positive leukocytes. The full gating strategy is shown in Supplementary Figure S1.

Within the CD45^+^ singlet gate, activated CD4^+^ T-cells were defined as CD3^+^CD4^+^CD69^+^ and naive CD4^+^ T-cells as CD3^+^CD4^+^CD45RA^+^CD27^+^. Activated B-cells were defined as CD19+CD20+CD69+ and naive B-cells as CD19+CD20+IgD+CD27-. Classical monocytes were defined as CD14+CD16-CCR2+ and non-classical monocytes as CD14+CD16+CX3CR1+. Activated NK cells were defined as CD56 bright CD69+ and resting NK cells as CD56 dim.

For each lineage, an activation ratio was calculated as the frequency of the activated population divided by the frequency of the corresponding resting or naive population within the same sample. Ratios were used in place of absolute frequencies to normalize for inter-individual variation in total lineage abundance. Gates were applied consistently across all samples within a batch by a single analyst blinded to disease state.

### Clinical assessment

Per our protocol, psychometric scores are collected at each clinical encounter, including but not limited to encounters in which a research blood draw is obtained. Three psychometric scales were selected a *priori* as primary outcomes to provide complementary assessments of functioning, impairment, and symptom burden: the Children’s Global Assessment Scale (CGAS), the PANS Global Impairment Scale (GIS), and the PANS 31-Item Symptom Rating Scale (PANS-31). CGAS is an established clinician-rated measure of overall functioning^57^. GIS is a caregiver-rated measure of overall PANS-related impairment^58^. PANS-31 is a newer caregiver-rated measure of PANS-specific symptom burden ^59^. As the aggression item of the PANS-31 was unavailable for all samples, total scores were calculated by summing the remaining 30 items. Exploratory secondary analyses included the Children’s Yale-Brown Obsessive-Compulsive Scale (CY-BOCS), Yale Global Tic Severity Scale (YGTSS), Modified Overt Aggression Scale (MOAS), and Columbia Impairment Scale (CIS) to assess consistency of associations across measures. CY-BOCS, YGTSS, and CIS assess specific symptom domains, whereas CIS was included as a sensitivity analysis for functional impairment score. Higher scores indicate greater symptom severity on all scales except CGAS, for which higher scores indicate better functioning. All psychometrics were completed before immunophenotyping experiments. Immunophenotyping and analyses of flow cytometry data were performed blinded to patient group, disease state, and psychometric scores.

Medication exposure was assessed relative to the date of blood collection, including nonsteroidal anti-inflammatory drug (NSAID) use at time of sampling, intravenous or oral corticosteroid use within the preceding 6 weeks; intravenous immunoglobulin (IVIG) within 3 months, maintenance immunomodulatory therapy (hydroxychloroquine, methotrexate, mycophenolate mofetil, or rituximab) within 6 months; and azithromycin within 7 days. A composite variable indicating any immunomodulatory exposure across these categories was also derived. Medication exposures at the time of sampling and within the specified preceding windows are summarized in Appendix Table 2.

## Statistical analysis

### Primary analysis of B-cell, CD4 T-cell, NK-cell and monocyte phenotypes

Analyses were performed in Python (3.14.0); statsmodels (0.15.0), SciPy (1.18.1). In figures, boxes denote median and interquartile range, whiskers extend to 1.5 times the interquartile range, and overlaid points represent individual samples. Differences across the three groups were assessed by Kruskal-Wallis test. Where the omnibus test was significant, pairwise comparisons were performed using two-tailed Mann-Whitney U tests. Paired flare and recovery samples from the same individual were compared using the Wilcoxon signed-rank test.

To evaluate whether sample storage duration influenced immune-cell measurements, storage time (months from blood draw to measurement) was compared with each activation ratio using Spearman correlation within each group (healthy control, recovery, flare), Wilcoxon rank-sum (Mann-Whitney U) tests comparing samples above and below the median storage time, and a rank-based linear model adjusting for group.

Among flare samples, differences in B-cell, CD4+ T-cell, monocyte, and NK-cell activation ratios between improving and worsening flares were assessed using separate linear regression models, with ratios as the dependent variable, improving flare as the reference group, and FDR multiple testing correction applied.

### Analysis of B-cell, CD4 T-cell, monocyte, and NK-cell subset ratios with psychiatric symptoms and functional impairment

Associations between immune activation ratios and psychiatric symptom severity were evaluated by linear mixed-effects models with participant-level random intercepts to account for repeated flare and recovery measurements (clinical severity ∼ immune activation ratio (1 | participant)). Complete case analysis was conducted for individual clinical scores. Model assumptions were assessed using residual diagnostics, and intraclass correlation coefficients (ICCs) quantified within-participant clustering. False discovery rate (FDR) correction was applied across the four immune-cell ratios separately within each primary clinical outcome. Random intercepts were retained in models with near-zero estimated between-participant variance to reflect the repeated-measures design. The same methods were applied to the secondary clinical outcomes (CY-BOCS, YGTSS, MOAS, and CI).

Sensitivity analyses assessed outlier exclusion and separate adjustment for potential confounders, including sex, clinical status (flare versus recovery), and immunomodulatory medication exposure. Exploratory interaction models evaluated potential effect modification by these variables. Interactions involving clinical status were interpreted cautiously because disease-state classification partly reflects symptom severity. Additional sensitivity analyses of MOAS scores were conducted using linear-mixed effects models with log-transformed scores and logistic mixed-effects models assessing the presence versus absence of aggression (MOAS > 0).

Exploratory symptom-specific analyses examined associations between four standardized immune activation ratios and eight clinician-selected symptom domains using linear mixed-effects models. Obsessive-compulsive symptoms and anxiety were defined by the maximum score of their respective items. Nominal p-values were used for hypothesis generation, with FDR-adjusted results also presented for transparency.

Post hoc analyses examined activated and naïve B-cell contributions to the ratio-severity associations. Observations exceeding the upper outlier threshold (Q3 +1.5 x IQR) for either component were excluded. Standardized components and their ratios were evaluated using identical observations within each outcome, with model fit compared using the Akaike information criterion (AIC) and Bayesian information criterion (BIC). FDR correction applied to the component analyses.

All Aim 2 analyses were conducted in R version 4.6.0 using packages dplyr, lmerTest, lme4, purrr, tibble, ggplot2, scales, gt, and patchwork.

### Exploratory analyses

Phospho-signaling was quantified as the median intensity of each phospho-specific marker within manually gated B-cell, CD4+ T-cell, and monocyte populations. Flare and recovery samples from the same patient were compared using the Wilcoxon signed-rank test, and independent groups were compared using the Mann-Whitney U test. Given the exploratory nature of these analyses, nominal p-values are reported without correction for multiple comparisons.

## Results

Demographic and clinical characteristics at the time of blood collection are presented in Appendix Table 1. The study included 19 flare samples, 15 recovery samples, and 20 healthy controls, with comparable age and sex distributions across groups. Compared with recovery, flare samples showed greater neuropsychiatric severity, reflected by lower CGAS scores (56.1 vs 82.8) and higher Global Impairment (45.3 vs 13.2) and PANS-31 scores (22.9 vs. 10.3). Immunomodulatory treatment and NSAID use were common in both disease states. Prior to flare blood draw, 3/19 patients were exposed to azithromycin which were used to treat infection. Prior to flare blood draw, 13/19 patients were exposed to at least one immunomodulatory medication. Prior to recovery blood draw, 11/15 patients were exposed to at least one immunomodulatory medication. The mean interval between flare onset and blood collection was 10.8 weeks (7.0 SD). Additional medication exposures are shown in Appendix Table 2. Additional clinical measures, including YGTSS, CY-BOCS, MOAS, and CIS, are summarized in Appendix Table 3. Participant characteristics at initial IBH are presentated in Supplementary Table S1.

### Primary analysis of B-cell, CD4 T-cell, NK-cell and monocyte phenotypes

All four immune-cell activation ratios (ratio of activated vs naive or resting for B-cells, CD4 T-cells, NK cells and Monocytes) were higher during PANS flare state (Figure 1). Compared with healthy controls, flare samples had higher immune-cell activation ratios (all four subsets p < 0.01). Compared with recovery, flare samples had higher immune-cell activation ratios (all four subsets p < 0.03). No significant differences were observed between immune-cell activation ratios between recovery and healthy controls.

**Figure 1.**
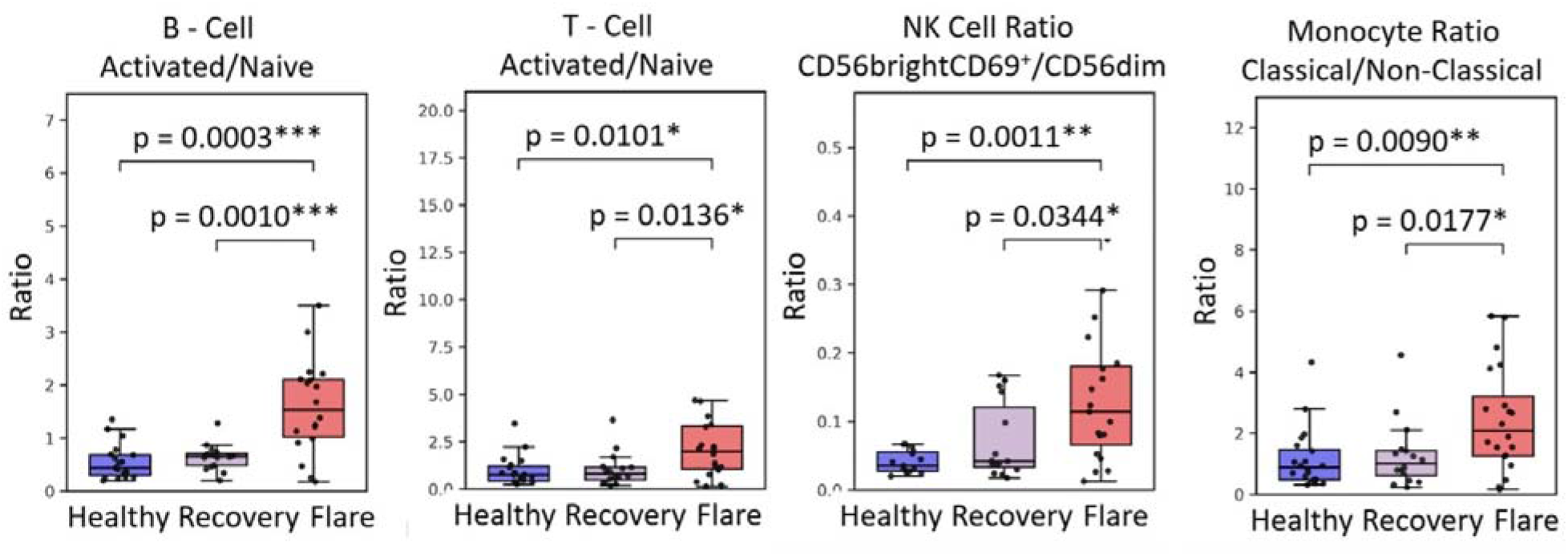
Peripheral immune-cell phenotypes in healthy controls, PANS recovery and PANS flare. Frequencies of activated B-cells, and naïve and activated CD4+ T-cells, CD56 bright and CD56 dim (NK) cells, non-classical and classical monocytes are shown alongside their corresponding ratios. Phenotypic markers are indicated above each panel. Boxes show medians and interquartile ranges, whiskers show the most extreme values within 1.5 times the interquartile range, and points represent individual specimens. Group comparisons used Kruskal-Wallis tests with post hoc Mann-Whitney U tests following significant omnibus tests. Paired flare-recovery comparisons used Wilcoxon signed-rank tests. All pairwise tests are two-sided.

Storage-time sensitivity analysis was conducted. Storage time was not associated with B-cell, CD4+ T-cell, NK-cell, or monocyte activation ratios within any group (all p > 0.11) or after adjustment for group (all p > 0.05) (Supplementary Figure S3)

Within the flare group, immune-cell activation ratios differed between samples obtained during an improving trajectory (henceforth called “improving flares”) (n=10) and those with a worsening trajectory or at peak of symptoms (henceforth these are called “worsening flares”) (n=9). Worsening flares were significantly associated with higher B-cell activation ratios (β=1.05, FDR p = 0.020) and lower monocyte classical/non-classical ratios (β= -2.46, FDR p = 0.002) compared with improving flares (Figure 2). CD4+ T-cell activation ratios were higher in the improving flare, although the difference was not significant after multiple testing corrections. NK-cell CD56 bright/CD56 dim ratios did not differ between groups.

**Figure 2.**
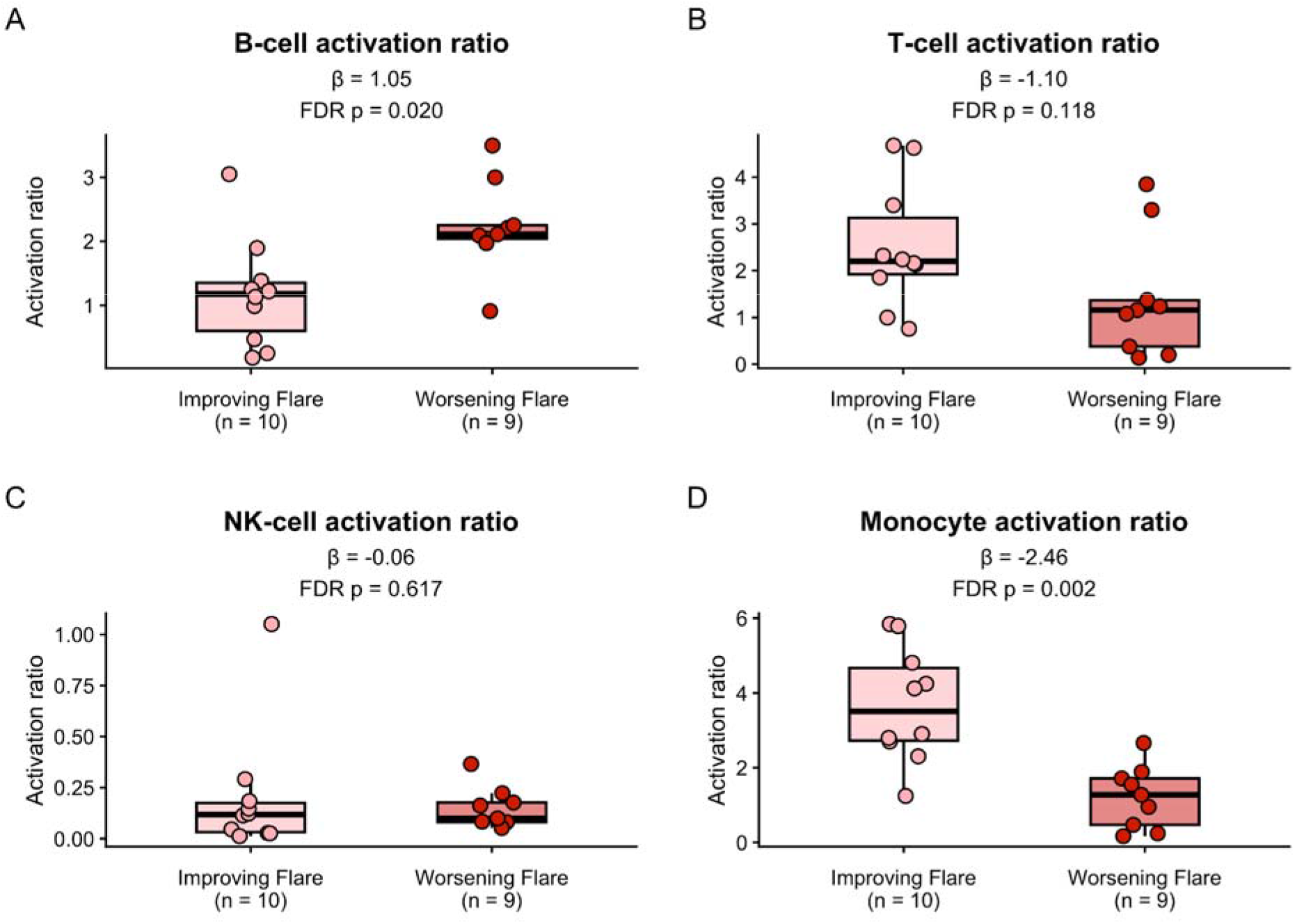
Comparison of immune-cell subset ratios across all four cell types (B, T, Monocyte, and Natural Killer (NK) cells): improving flare (n=10) vs worsening flare/at peak of symptoms (n=9). Points represent individual observations. Boxes span the interquartile range, and horizontal lines mark the median. Beta (β) is the estimated difference in immune-cell subset ratio between improving flare v worsening flare. False Discovery Rate (FDR) multiple testing corrections were applied.

### Analysis of B-cell, CD4 T-cell, monocyte, and NK-cell subset ratios with psychiatric symptoms and functional impairment

Higher B-cell activation ratios were consistently associated with greater psychiatric symptom severity and functional impairment across all three primary clinical measures after FDR correction (Figure 3). Each one-unit increase in the B-cell activation ratio was associated with lower clinician rated function (CGAS scores) (β= -14.89, FDR p < 0.001), higher Global Impairment (parent rating of psychiatric symptom severity) (β=12.81, FDR p=0.004), and high PANS-31 scores (β=8.37, FDR p= 0.012). In contrast, CD4+ T-cell, monocyte, and NK-cell activation ratios were not significantly associated with any of the three primary clinical measures (all p > 0.05). Associations with additional clinical measures and MOAS sensitivity results are presented in Supplementary Table S3. Residual diagnostics indicated no substantial departures from model assumptions.

**Figure 3.**
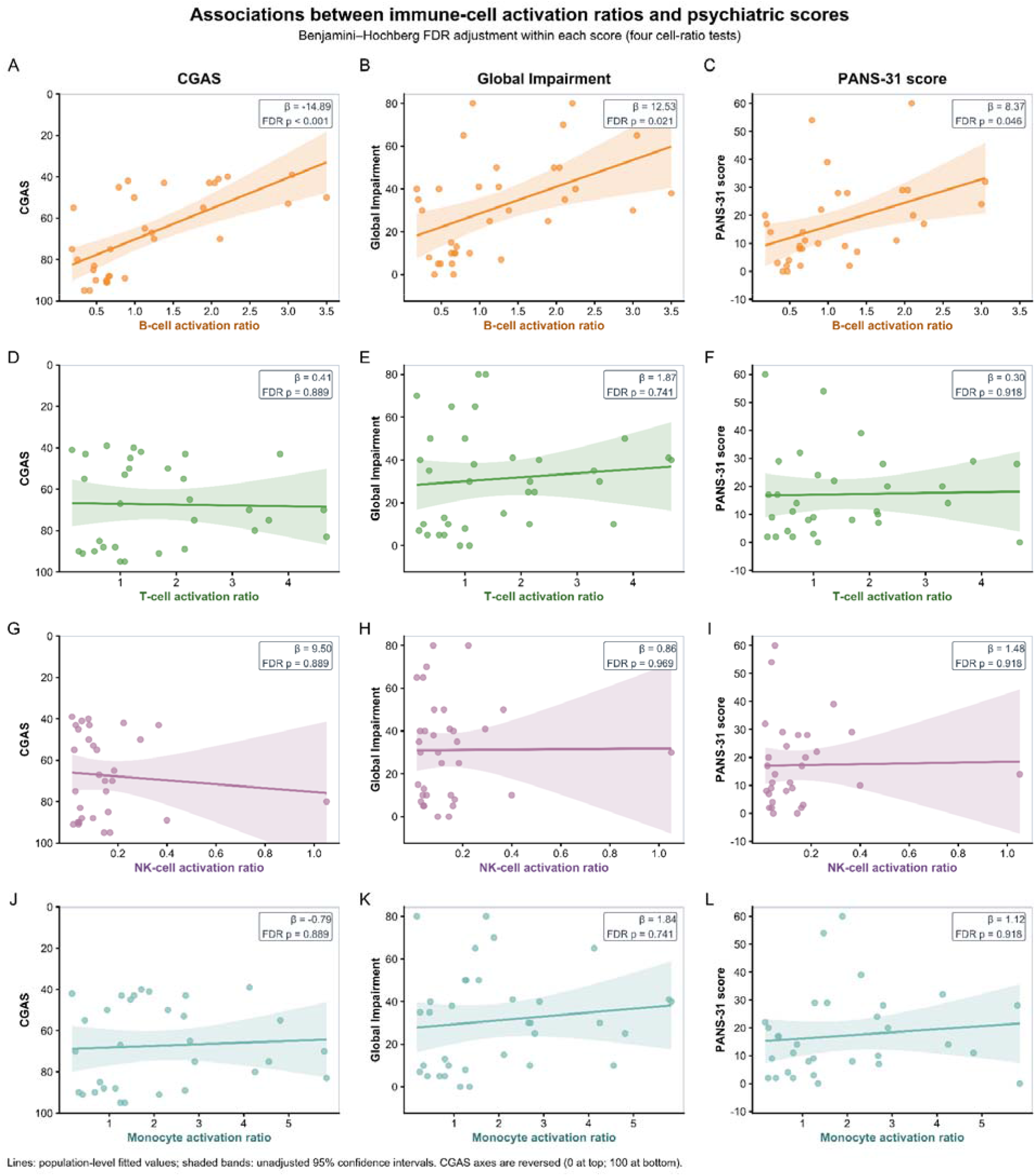
Associations between immune-cell activation ratios and psychiatric scores. Columns show Clinician’s Global Assessment Scale (CGAS), Global Impairment, and PANS-31 score. Rows show B-cell (A-C), CD4+ T-cell (D-F), NK-cell (G-I) and monocyte (J-L) activation ratios. Lines and shaded bands represent population-level fitted values and 95% confidence intervals from linear mixed-effect models with a participant-level random intercept to account for repeated flare and recovery measurements. The CGAS y-axis is reversed so that greater impairment is displayed higher in all panels. β denotes the estimated score change per one-unit increase in the corresponding activation ratio. P-value were adjusted using the Benjamini-Hochberg false discovery rate (FDR) separately within each psychiatric score across the four immune-cell ratio tests.

#### Sensitivity Analysis

Exclusion of the sample with an outlying NK-cell activation ratio did not materially alter the results, with associations remaining nonsignificant (Supplementary Table S4. Additional sensitivity analyses separately adjusted for clinical status (flare versus recovery), immunomodulator medication exposure, and sex. Adjustment for medication exposure or sex did not materially alter the primary associations. As expected, adjustment for clinical status (flare vs recovery) attenuated associations between B-cell activation and psychiatric symptom severity since the measures are interdependent (flare status determination is based on assessment of psychiatric symptom severity; psychiatric symptom severity increases during disease flares); however, the association with CGAS remained significant (Supplementary Table S5).

#### Exploratory Analyses

##### Interaction Analysis

Exploratory analyses assessed potential effect modification by clinical status, sex, and immunomodulatory medication exposure (Supplementary Figures S20-S28). A nominally significant interaction between CD4+ T-cell activation ratio and clinical status was observed for CGAS (p for interaction = 0.026), with higher CD4+ T-cell activation ratios associated with higher predicted CGAS scores during flare. Monocyte activation ratios exhibited a similar directional pattern, although the interaction was not statistically significant (p for interaction = 0.12).

No significant interactions were observed with sex or immunomodulatory medication exposure. However, the inverse association between B-cell activation ratio and CGAS appeared stronger among patients exposed to immunomodulatory medications (p for interaction = 0.054) possibly driven by a confounder (severity of psychiatric symptoms may influence probability of pursuing immunomodulation). Given the small sample size and limited statistical power, these exploratory findings require validation in larger cohorts.

##### Symptom-specific analyses

Exploratory symptom-specific analyses identified nominal positive associations between B-cell activation ratios and anxiety, mood, oppositional behavior, attention difficulties, and sleep disturbances (β=0.40-0.48), and between monocyte activation ratios and obsessive-compulsive symptoms (β=0.63; Figure 4).

**Figure 4.**
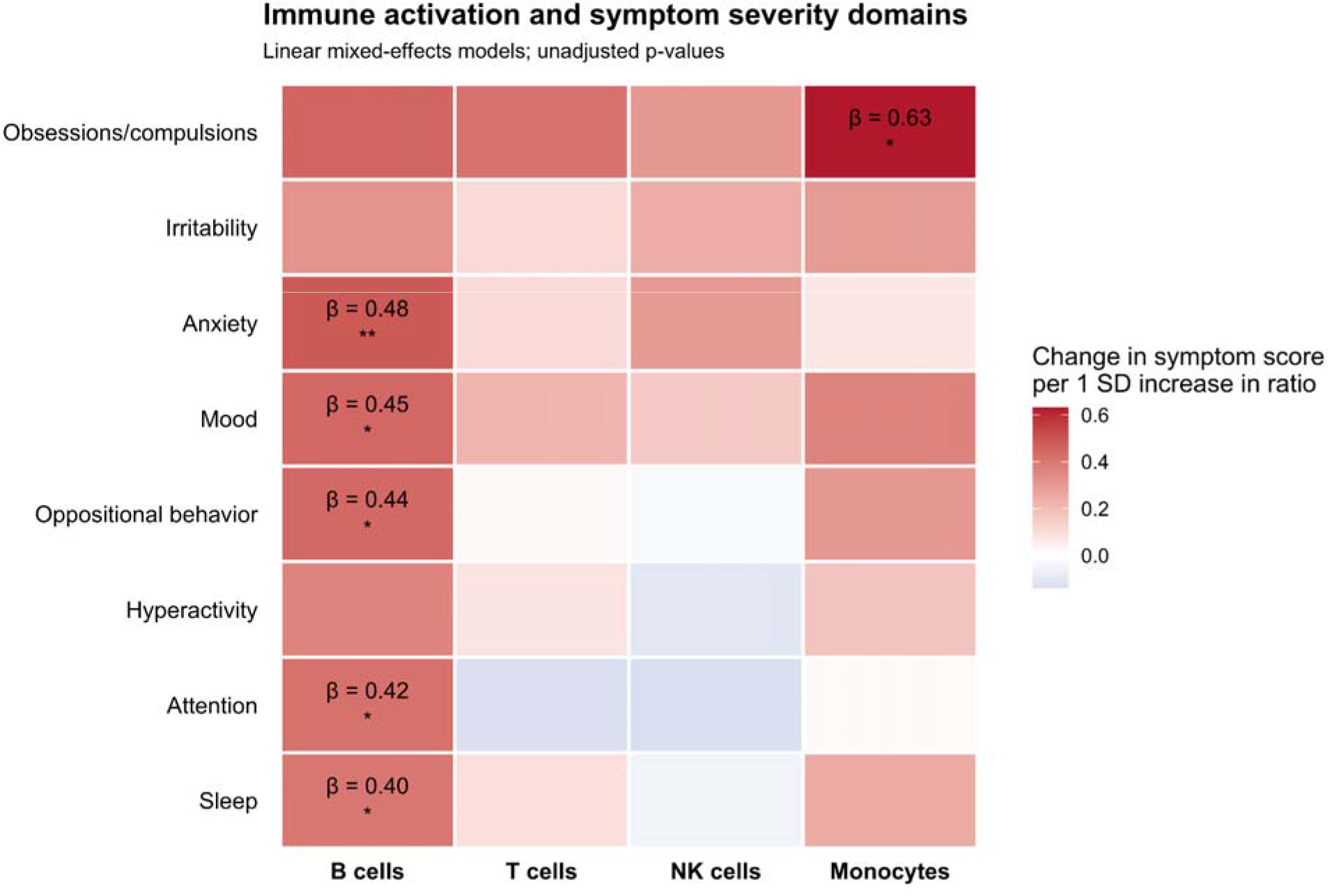
Associations between immune-cell activation and symptom severity. Each tile shows the estimated difference in symptom severity (0-4, higher scores indicate greater severity) per 1-SD higher activation ratio. Ratios were standardized within each cell type to make estimates comparable across cell types. Linear mixed-effects models include a participant random intercept. Red and blue indicate positiv and negative associations, respectively. Estimates are shown where unadjusted *p < 0.05 and **p<0.01.

##### B-cell ratio component analyses

After exclusion of 5 extreme observations, the B-cell activation ratio remained associated with greater clinical severity across all three primary psychometrics after FDR correction and exhibited larger standardized coefficients than either component alone (Figure 5). Higher activated B-cell levels were significantly associated with lower CGAS and higher Global Impairment scores, with a similar direction of association for PANS-31. Naïve B-cell levels showed nonsignificant associations in the opposite direction, consistent with lower impairment at higher levels. Across all three outcomes, models including the B-cell activation ratio had lower AIC and BIC than models including either component alone. Results retaining extreme observations are presented in Supplementary Table S6.

**Figure 5.**
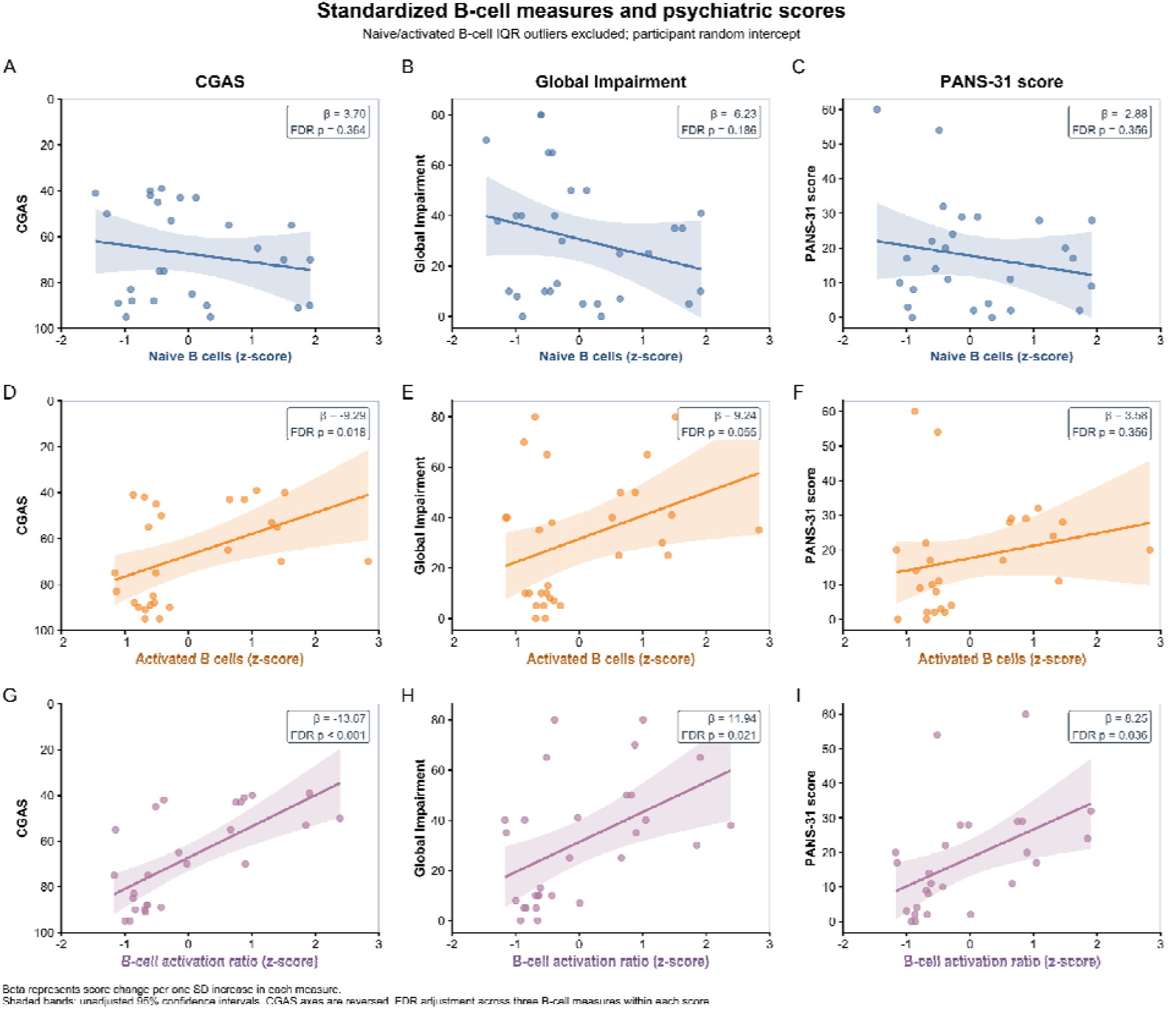
Associations of naïve B-cells (CD19^+^ CD20^+^ IgD^+^ CD27^-^), activated B-cells (CD19^+^ CD20^+^ CD69^+^), and the B-cell activation ratio (activated/naïve) with clinical severity scales. Associations with Children’s Global Assessment Scale (CGAS), Global Impairment scores, and PANS-31 are shown for standardized naïve B-cell frequencies (A-C), activated B-cell frequencies (D-F), and activated-to-naïve B-cell ratios (G-I). Separate linear mixed-effects models included a participant random intercept and used flare and recovery observations. Points represent individual specimens, lines indicate population-level fitted values, and shaded bands show 95% confidence intervals. β estimates represent score difference per 1-SD higher B-cell measure. Observations exceeding the upper outlier threshold (Q3 + 1.5 x IQR) for either naïve or activated B-cell frequency were excluded. Models used identical observations across th three B-cell measures within each clinical score. P-values were adjusted using FDR correction across the three B-cell measures within each score. CGAS axes are reversed, with lower scores indicating greater impairment.

##### Phospho-signaling analyses

Exploratory analyses identified nominal differences in intracellular signaling across B-cell, CD4+ T-cell, and monocyte populations (Supplementary Figure S2). pSTAT5 was higher during recovery than flare in activated B-cells (p = 0.029) and activated T-cells (p = 0.0003). Compared with healthy controls, flare samples showed higher pS6 in activated T-cells (p = 0.0001) and lower pSTAT3 (p = 0.0003) and pCREB (p = 0.0016) in classical monocytes. These findings were not corrected for multiple comparisons and are considered hypothesis-generating. Across the remaining marker–population combinations, phospho-signaling did not differ significantly among healthy controls, recovery, and flare in either the improving-or worsening-flare cohorts (Supplementary Figures S1–S16). Isolated nominal differences (5 of 112 comparisons; p < 0.05) were consistent with the number expected by chance, and more detailed analyses with larger cohorts will be needed to characterize disease-state–specific signaling.

## Discussion

In this study, activation ratios of B-cells, CD4+ T-cells, monocytes, and NK cells were higher during PANS flares than during recovery or in healthy controls, indicating broad peripheral immune activation linked to disease state. However, only the B-cell activation ratio was associated with greater psychiatric symptom severity and functional impairment, and this held across all clinical psychiatric measures. Flare trajectory also mattered. Compared with samples collected during improving flares, samples collected during worsening flares or at peak symptoms showed higher B-cell activation but lower monocyte activation. The increases in activated CD4+ T-cells, activated NK cells, and pro-inflammatory monocytes fit with the peripheral inflammation previously described in this population, including enthesitis-related arthritis (ERA) ^41,42^. All three cell subsets have been implicated in the pathogenesis of pediatric ERA^60–62^. Together, these findings suggest that peripheral immune activation is a feature of PANS flares in general. The immune features most closely tied to the severity of psychiatric symptoms may be distinct, with B-cell activation emerging as especially relevant to psychiatric disease expression.

The broad elevation of immune-cell ratios during flare, followed by their attenuation during recovery, suggests that peripheral immune alterations in PANS are dynamic and potentially reversible. These changes may reflect inflammatory stimulation accompanied by shifts in immune-cell differentiation, redistribution, and regulatory function. Consistent with this interpretation, Rahman et al. (2025) reported enrichment of pro-inflammatory M1-like monocytes during PANS flare and anti-inflammatory M2-like monocytes during recovery^53^. Rahman et al. also identified CNS trafficking-associated monocyte phenotypes expressing regulatory cytokines, including IL-10 and TGF-β, which suggests that clinical improvement may involve active immunoregulatory processes rather than simply diminished inflammatory stimulation. Similar patterns of immune-cell remodeling have been described in relapsing-remitting autoimmune diseases. Longitudinal studies of systemic lupus erythematosus (SLE) identified distinct B-cell, CD4+ T-cell, and innate immune-cell profiles associated with persistent disease activity and subsequent quiescence, illustrating how immune-cell populations may exhibit different relationships with disease progression and resolution ^63^. In our study, elevated ratios during flare were primarily accompanied by higher frequencies of activated or inflammatory populations (across all subsets), whereas naïve, inactive and nonclassical populations tended to be more abundant in healthy controls than during flare or recovery, although these differences were not all statistically significant. This pattern may indicate that immune-cell remodeling in PANS involves both reversible changes in activation-associated phenotypes and more persistent alterations in cellular composition. Accordingly, clinical recovery may reflect restoration of aspects of immune regulation without complete normalization of the peripheral immune-cell landscape.

The B-cell findings provide more specific insight into the immunological processes associated with neuropsychiatric manifestations. Across all three primary clinical outcomes, the activated-to-naïve B-cell ratio exhibited consistent associations with symptom burden and functional impairment, than either population alone. These converging results suggest that the relative balance of activated and naïve B-cells may capture clinically relevant immune dysregulation more effectively than either component alone. B-cell homeostasis is maintained through tightly regulated processes governing activation, differentiation, and peripheral tolerance. In systemic autoimmunity, defective tolerance checkpoints and sustained stimulation through B-cell receptor, B-cell activating factor (BAFF), and Toll-like receptor signaling can disrupt this balance, promoting the persistence and differentiation of activated B-cell populations^64^. CD69, the activation-associated marker used to define our activated B-cell population, is involved in lymphocyte activation and the regulation of cellular trafficking, providing a potential functional link between altered activation phenotypes and immune-cell behavior. Dysregulated B-cell activation and differentiation are central to SLE and multiple sclerosis (MS)^65,66^, while changes in CD69-expressing B-cell populations following immunomodulatory treatment in MS further highlight the clinical relevance of this phenotype^67^. In rheumatoid arthritis, B-cell phenotypes have long been investigated as markers of disease activity, clinical heterogeneity, and therapeutic response^68–70^. These studies provide a biological framework in which an increased activated-to-naïve B-cell ratio may reflect altered activation and differentiation processes associated with greater neuropsychiatric severity in PANS. The better model fit of the ratio further suggests its potential as a candidate biomarker.

One mechanism through which altered B-cell activity may contribute to neuropsychiatric manifestations involves the generation of neuronal-reactive antibodies. In a Yale-Stanford study, IgG isolated from plasma collected during PANS flares bound to striatal cholinergic interneurons and altered neuronal activity, effects not observed with remission plasma^24^. Striatal cholinergic interneurons modulate neural circuits involved in motor control, behavioral regulation, and cognitive function, providing a biologically relevant connection between humoral immune responses and the characteristic neuropsychiatric manifestations of PANS. B-cell activation and differentiation are integral to the generation and maintenance of antibody responses, raising the possibility that the altered peripheral B-cell phenotype identified in our study accompanies changes in neuronal-reactive humoral immunity. This relationship is particularly relevant given the association between B-cell activation and clinical severity, suggesting that differences in peripheral B-cell homeostasis may reflect immunological processes linked to neuronal dysfunction.

Beyond antibody production, B-cells contribute to immune regulation through antigen presentation, cytokine secretion, CD4+ T-cell co-stimulation, and interactions with other immune populations. In autoimmune CNS disorders, including MS and autoimmune encephalitis, these functions contribute to neuroinflammatory processes alongside antibody-mediated mechanisms ^71^. Experimental evidence further implicates B-cells in neurological and behavioral dysfunction beyond classical CNS autoimmunity. In mouse models of Alzheimer’s disease, activated B-cells accumulated in the brain and contributed to neuropathology and cognitive impairment, whereas B-cell depletion attenuated these abnormalities^72^. Emerging literature on stress-related and depressive disorders also suggest that B-cell associated immune responses influence neurobehavioral regulation through peripheral inflammatory signaling and interactions with CNS immune pathways^73^. These mechanisms may be particularly relevant to PANS, in which neuropsychiatric manifestations have been linked to altered basal ganglia circuitry and immune-mediated neuronal dysfunction^23,24^. B-cell antigen presentation and cytokine signaling could influence neuroinflammatory pathways involving T-cells and the neurovascular interface. Experimental models of post-streptococcal neuroinflammation have demonstrated Th17-associated blood-brain barrier disruption and CNS inflammation^35^, providing a plausible route through which peripheral adaptive immune responses may influence neural function. These findings raise the possibility that the association between peripheral B-cell activation and neuropsychiatric severity in PANS may reflect broader B-cell-associated neuroimmune processes affecting CNS function.

Exploratory phospho-signaling analyses suggested that recovery may be accompanied by increased STAT5 signaling in activated B and T-cells, consistent with IL-2–dependent homeostatic signaling, whereas flare was associated with increased mTOR activity (pS6) in activated T-cells relative to healthy controls. Flare was also associated with reduced STAT3 and CREB signaling in classical monocytes relative to healthy controls. Given the number of comparisons and modest effect sizes, these findings should be considered hypothesis-generating and require confirmation in larger cohorts. Most phospho-signaling comparisons were not significant (Supplementary Figures (S4–S19), and the limited sample size precluded robust assessment. Deeper analyses, including stimulation-based phospho-flow and larger longitudinal cohorts, will be needed to define signaling changes associated with PANS disease activity.

Collectively, our findings add to the growing evidence supporting an immune component in the pathophysiology of PANS. Previous studies have demonstrated systemic inflammatory and autoimmune abnormalities^41^, disease-state-dependent myeloid-cell phenotypes ^53^, altered immune-related transcriptional and functional responses ^74^, and neuronal-reactive antibodies capable of modifying neuronal activity^24^. These complementary studies span cellular phenotyping, molecular immune responses, and experimental evidence of neuronal dysfunction. By demonstrating broad changes in peripheral immune phenotypes during flare alongside a selective relationship between B-cell activation and neuropsychiatric symptom severity, our study adds a clinically relevant dimension to this evidence. Together, this convergent evidence supports a model in which immune dysregulation constitutes an important biological component of PANS, with distinct immune subsets potentially contributing to disease activity and the severity of neuropsychiatric manifestations.

Strengths of this study include the clinically well-characterized Stanford Immune Behavioral Health (IBH) PANS cohort, longitudinal samples collected during flare and recovery, and comparison with matched healthy controls from the same community as the patients who had samples processed using the same pipeline, and assessment of multiple immune populations alongside complementary measures of neuropsychiatric severity and functioning. The consistency of B-cell associations across clinical outcomes, supported by component analyses, further strengthens the findings. Nevertheless, the relatively small sample has limited precision, particularly for covariate adjustment and exploratory analyses, while the observational design precludes causal inference. Prolonged storage and PROT1 fixation may reduce activation-marker detection, potentially underestimating activation levels. However, although flare samples were stored longer than controls, sensitivity analyses did not indicate a storage-related effect. Standardized processing minimized procedural variability, while activation-to-naive cell ratios may be less sensitive to uniform signal loss. Adjustment for clinical status attenuated the B-cell associations with Global Impairment and PANS-31, so we cannot fully separate effects of disease state from those of symptom severity. Medication classification did not capture treatment dose, duration, or indication, leaving potential residual confounding. Near-zero random-intercept variance estimates in several models also limited characterization of participant-level variability. Activation ratios were based on a limited set of predefined populations. Flare samples were collected a mean of 10.8 weeks after flare onset thus the initial phase of the illness was missed. Lastly, findings were from a single-center cohort thus requiring future external validation.

In conclusion, our findings show increased activated immune-cell phenotypes during PANS flare and identify the activated-to-naïve B-cell ratio as a candidate biomarker of neuropsychiatric severity that warrants validation. Its consistent clinical associations and better model fit relative to the individual B-cell populations suggest the potential importance of B-cell homeostasis in PANS pathophysiology. These observations also point to B-cell-associated immune pathways as candidates for therapeutic investigation. Larger longitudinal studies integrating single-cell RNA sequencing, B-cell receptor repertoire analysis, further subset delineation, and neuronal autoantibody profiling could elucidate the mechanisms linking B-cell activation to neuropsychiatric dysfunction and determine whether these phenotypes track clinical deterioration, recovery, and treatment response.

## Funders

Lucile Packard Foundation for Children’s Health. The funding sources had no role in the design and conduct of the study; collection, management, analysis, and interpretation of the data; preparation, review, or approval of the manuscript; and decision to submit the manuscript for publication.

## Acknowledgements

Past and present funding for the infrastructure of our research program, training, advocacy, sample collection and overlapping projects came from: 1) Lucile Packard Foundation for Children’s Health; 2) National Institute of Mental Health-Pediatrics and Developmental Neuroscience Branch which supported the initial creation of the Stanford University IBH Research Program; 3) The Neuroimmune Foundation for education/training, microbial sequencing, and medication analyses; 4) The Brain Foundation and O’Sullivan Foundation for Autism research; 5) The Tara & Dave Dollinger PANS Biomarker Discovery Core for patient sample collection; 6) Stanford Maternal and Child Health Research Institute (MCHRI) for HLA research; 7) Stanford SPARK, SPARK Pisa, and International OCD Foundation for immunophenotyping research; 8) Oxnard Foundation for imaging research; 9) Caudwell Children’s Foundation for genetics and immunology research; 10) The Louisa Adelynn Johnson Fund for Complex Disease for expanding the Illuminate PANS project; 11) PPN for funding our healthy control project; 12) Stanford COMET, DRIVE, and HBREX programs for student research in our program; 13) LPCH Volunteer Program. We also wish to thank our dedicated staff, collaborating clinicians, and the many additional donors who have contributed to our work. Most importantly, we extend our deepest gratitude to the patients, families, and healthy control volunteers; this research would not be possible without their trust and participation.

## Author Approval

All authors have reviewed and approved the final version of the manuscript and agree to its submission for publication.

## Competing interests

The authors declare that they have no competing interests.

## Declarations

This study was approved by the Stanford University Institutional Review Board (Protocol No. 26922). Written informed consent was obtained from parents or legal guardians, with participant assent obtained as appropriate.

## Data Availability Statement

The datasets generated and analyzed during the current study are available from the corresponding author upon reasonable request, subject to institutional and ethical restrictions.

## Appendix

**Appendix Figure 1:**
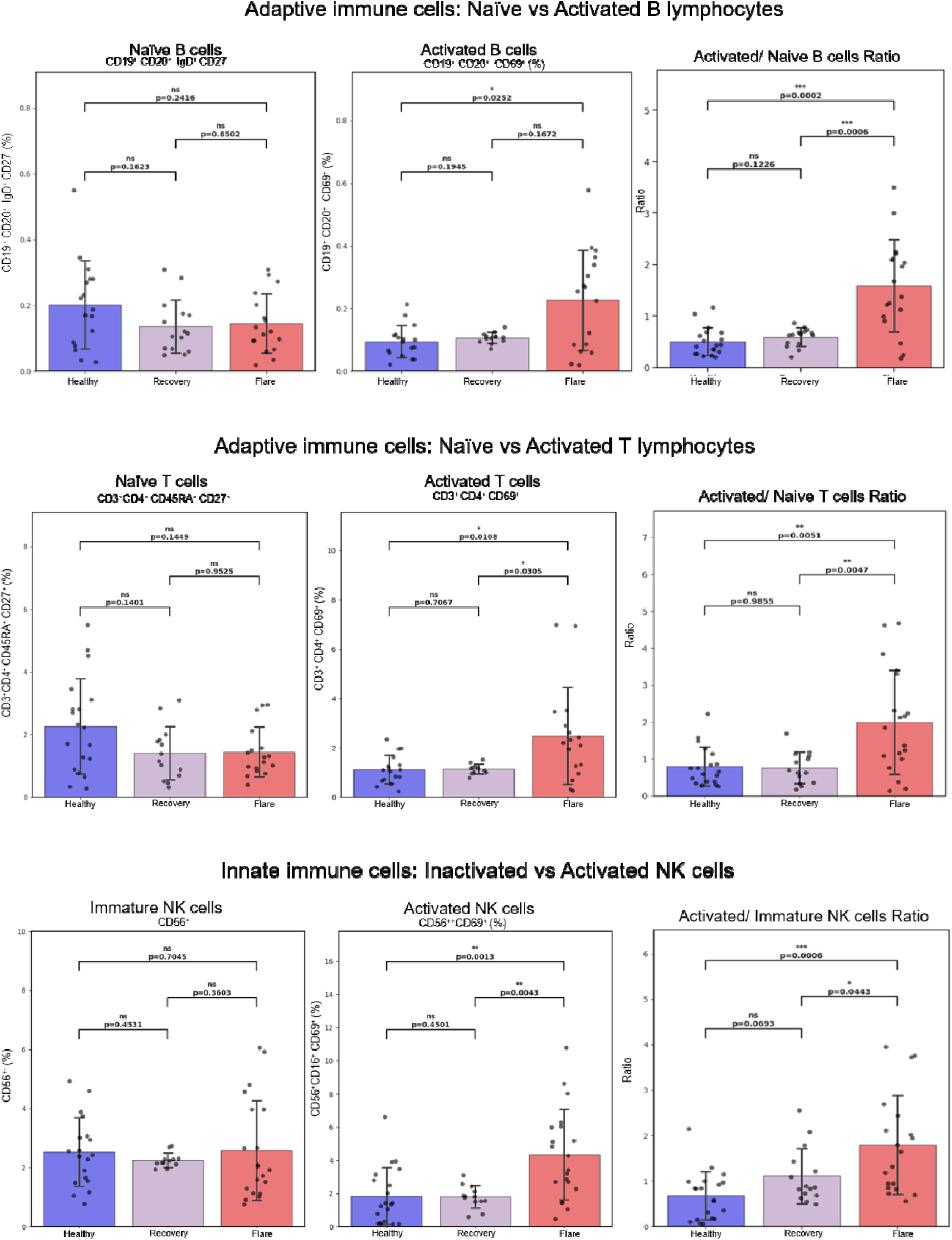
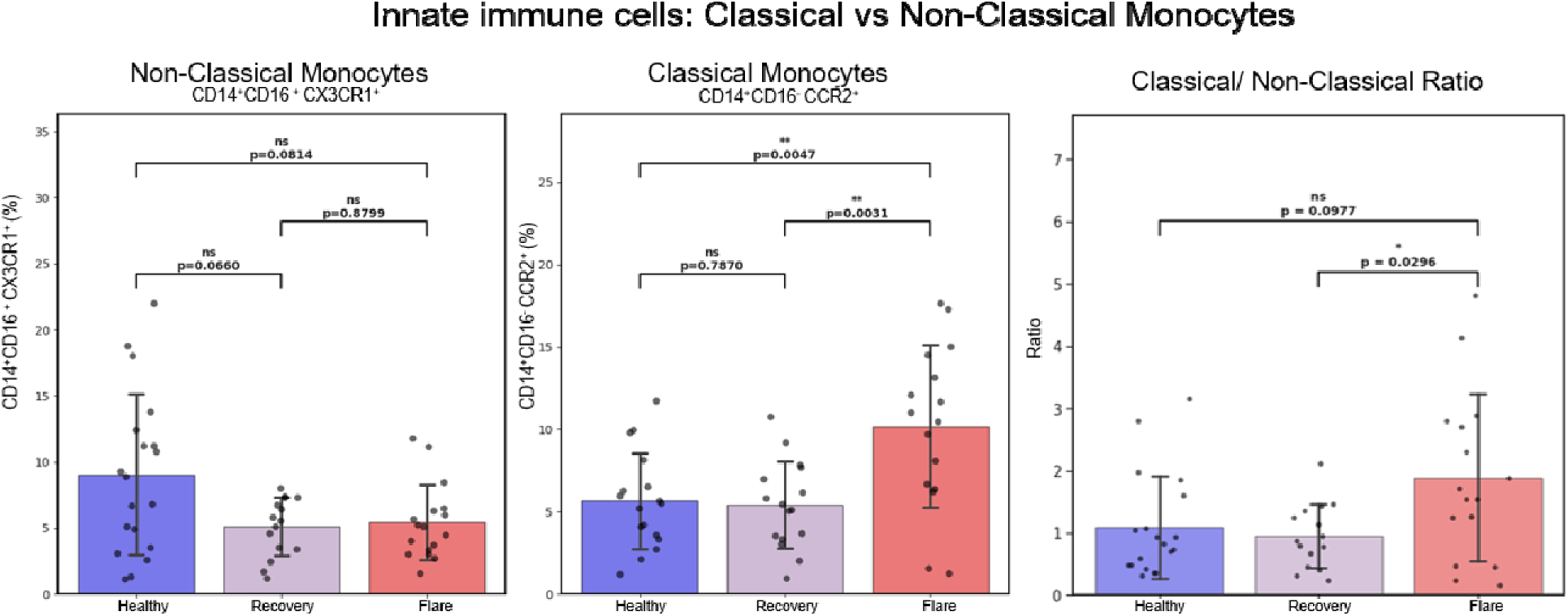
Peripheral immune-cell phenotypes in healthy controls, PANS recovery and PANS flare. Frequencies of naïve and activated B-cells, and naïve and activated T-cells, inactive and activated natural killer (NK) cells, non-classical and classical monocytes are shown alongside their corresponding ratios. Phenotypic markers are indicated above each panel. Boxes show medians and interquartile ranges, whiskers show the most extreme values within 1.5 times the interquartile range, and points represent individual specimens. Group comparisons used Kruskal-Wallis tests with post hoc Mann-Whitney U tests following significant omnibus tests. Paired flare-recovery comparisons used Wilcoxon signed-rank tests. All pairwise tests are two-sided.

**Appendix Figure 2.**
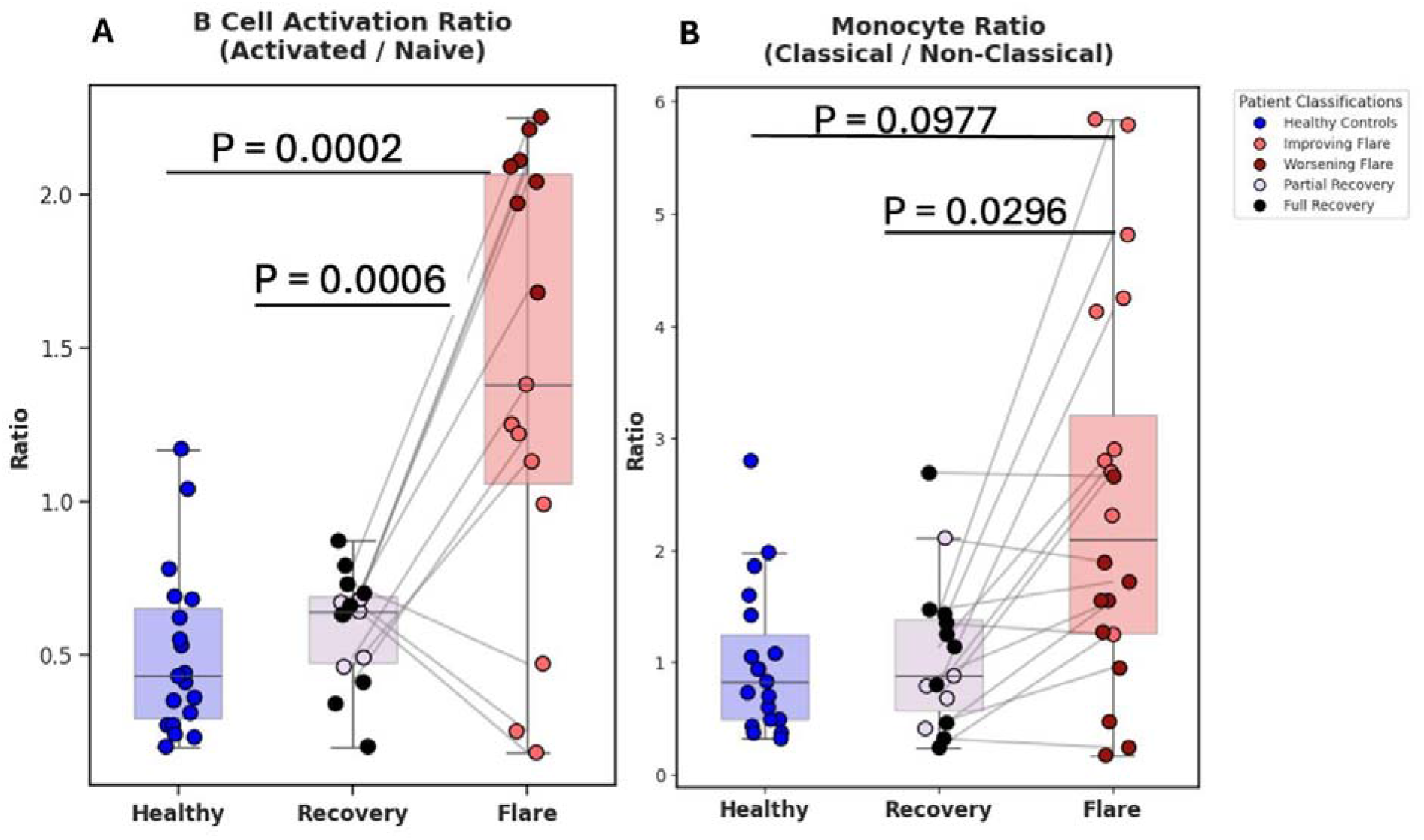
B-cell and monocyte activation ratios in healthy controls and in patients with PANS during recovery and flare. (A) B-cell activation ratio, defined as the frequency of activated (CD19+CD20+CD69+) B-cells divided by the frequency of naïve (CD19+CD20+IgD+CD27+) B-cells. (B) Monocyte ratio, defined as the frequency of classical (CD14+CD16+CCR2+) monocytes divided by the frequency of non-classical (CD14+CD16+CX3CR1+) monocytes. Samples were obtained from healthy controls (n = 20), patients with PANS during recovery (n = 15), and patients with PANS during flare (n = 19). In panel A, five observations exceeding the upper outlier threshold (Q3 + 1.5 × IQR) were excluded after confirming that they did not reflect data-entry or acquisition errors ([3] healthy control, [1] recovery, [1] flare). Flare trajectory (improving vs. worsening or at peak of symptoms) was determined from clinical notes before sample selection. Laboratory staff (NS, HM) and the analyst (IK) were blinded to disease state and flare trajectory. Each point represents one sample and is colored by clinical status: healthy control (blue), improving flare (light red), worsening flare (dark red), partial recovery (lavender), and full recovery (black). Grey lines connect paired flare and recovery samples from the same patient. Boxes show the median and interquartile range, and whiskers extend to the most extreme values within 1.5 × IQR. P-values are from two-sided Mann-Whitney U tests performed after a significant Kruskal-Wallis test, and only significant comparisons are shown.

**Appendix Table 1.** Demographic and Clinical Characteristics of the Study Population.

| **Characteristic** | **Flare**<br>N = 19<sup>*1*</sup> | **Recovery**<br>N = 15<sup>*1*</sup> | **Healthy Controls**<br>N = 20<sup>*1*</sup> |
|---|---|---|---|
| **Samples, n** | 19 | 15 | 20 |
| **Age at blood draw, years** | 8.9 (2.3) | 9.5 (2.4) | 9.3 (2.4) |
| **Sex** | — | — | — |
| Female | 8 (42%) | 5 (38%) | 8 (40%) |
| Male | 11 (58%) | 8 (62%) | 12 (60%) |
| **CGAS** | 56.1 (15.0) | 82.8 (15.5) | — |
| **Missing, n** | 1 | 2 | — |
| **Global Impairment** | 45.3 (17.1) | 13.2 (16.5) | — |
| **Missing, n** | 0 | 0 | — |
| **PANS-31 total score** | 22.9 (13.9) | 10.3 (13.6) | — |
| **Missing, n** | 2 | 1 | — |
| **Flare duration, weeks** | 10.8 (7.0) | — | — |
| **Any immunomodulatory treatment** | 13 (68%) | 11 (85%) | — |
| **NSAID use** | 11 (58%) | 9 (69%) | — |

<sup>*1*</sup> Mean (SD); n (%)
Continuous variables are presented as mean (SD); categorical variables as n (%). Data are summarized at the sample/visit level. Some PANS participants contributed both flare and recovery samples. Any immunomodulatory treatment is a composite encompassing the medication categories listed in methods. Missing counts are provided for psychiatric outcomes; these outcomes were not assessed in healthy controls.

**Appendix Table 2.**
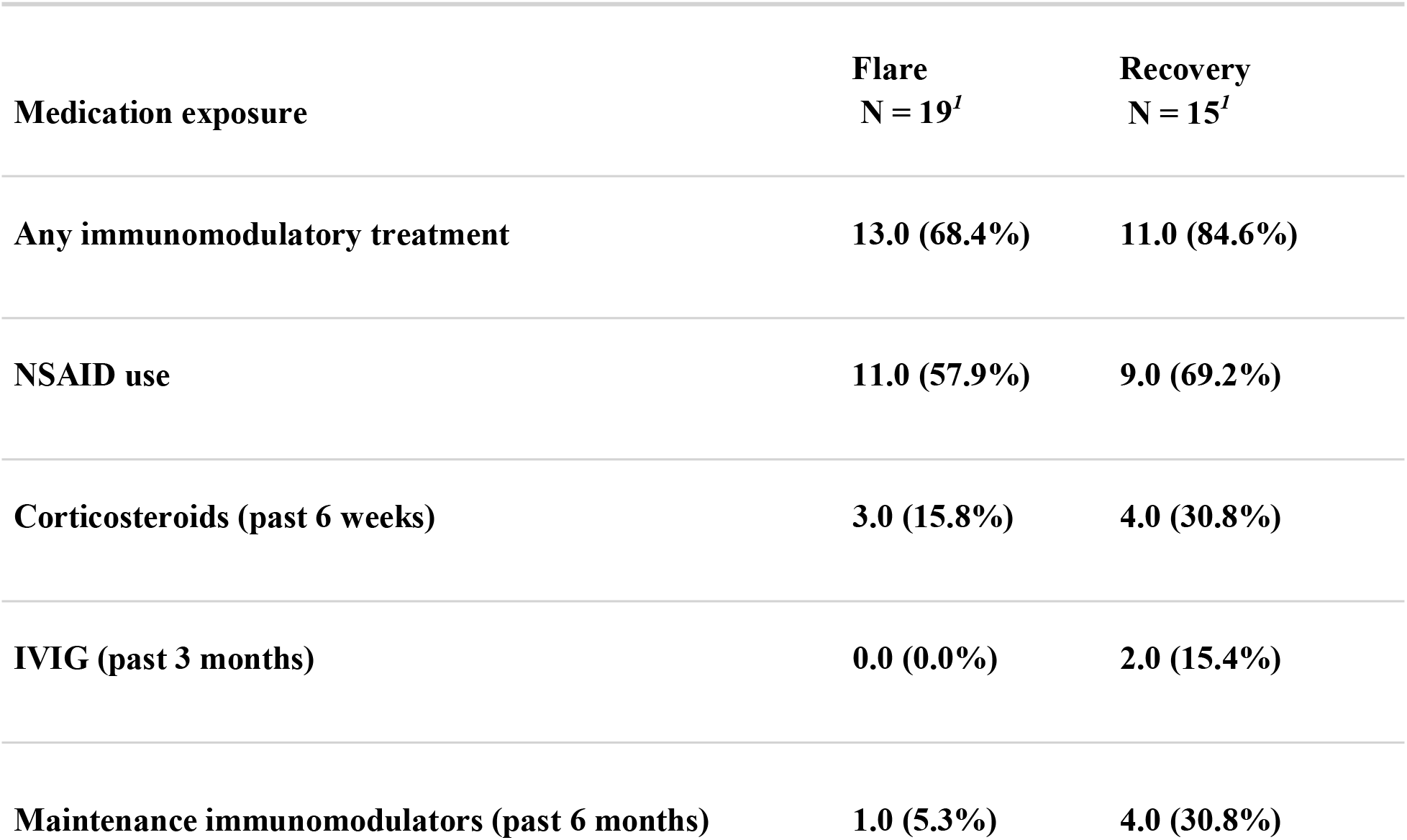
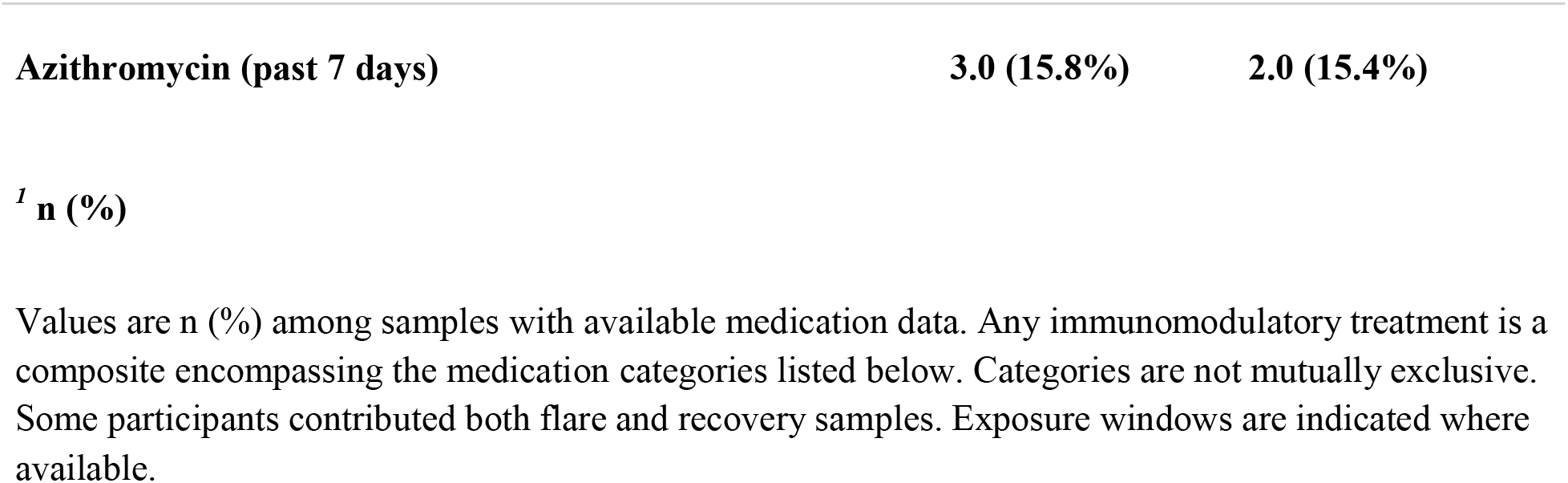
Medication Exposure by PANS Disease State.

| **Medication exposure** | **Flare**<br>**N = 19*<sup>1</sup>*** | **Recovery**<br>**N = 15*<sup>1</sup>*** |
|---|---|---|
| **Any immunomodulatory treatment** | **13.0 (68.4%)** | **11.0 (84.6%)** |
| **NSAID use** | **11.0 (57.9%)** | **9.0 (69.2%)** |
| **Corticosteroids (past 6 weeks)** | **3.0 (15.8%)** | **4.0 (30.8%)** |
| **IVIG (past 3 months)** | **0.0 (0.0%)** | **2.0 (15.4%)** |
| **Maintenance immunomodulators (past 6 months)** | **1.0 (5.3%)** | **4.0 (30.8%)** |
| **Azithromycin (past 7 days)** | **3.0 (15.8%)** | **2.0 (15.4%)** |

***<sup>1</sup>*** **n (%)**
Values are n (%) among samples with available medication data. Any immunomodulatory treatment is a composite encompassing the medication categories listed below. Categories are not mutually exclusive. Some participants contributed both flare and recovery samples. Exposure windows are indicated where available.

**Appendix Table 3.** Additional Clinical Outcomes by PANS Disease State.

| **Clinical outcome** | **Flare**<br>N = 19<sup>*1*</sup> | **Recovery**<br>N = 15<sup>*1*</sup> |
|---|---|---|
| **CY-BOCS total score** | 16.2 (6.7) | 7.4 (9.2) |
| Missing, n | 4 | 7 |
| **YGTSS total score** | 16.3 (14.9) | 7.5 (10.6) |
| Missing, n | 8 | 13 |
| **MOAS total score** | 7.2 (9.2) | 1.9 (3.9) |
| **Columbia Impairment total** | 18.3 (9.7) | 7.7 (9.3) |
| Missing, n | 4 | 2 |

<sup>*1*</sup> Mean (SD)
Values are mean (SD). Missing values are reported as counts. Characteristics are summarized at the sample/visit level; some participants contributed observations during both flare and recovery.

## Supplementary Material

### Supplementary Methods

#### Stanford University Immune Behavioral Health (IBH) Clinic Entry Screening Protocol

To ethically manage an overwhelming number of patient referrals that had become unsustainable, we sought guidance from the Stanford Ethics Committee. In consultation with them, we established a standardized screening protocol in September 2014, which is administered by a single nurse practitioner (BF). This protocol provides a fair and consistent method for managing clinic capacity and fulfilling our research mission by prioritizing local patients and individuals who facilitate research on early, untreated disease flares.

Priority is given to patients who meet the following criteria:

**1. Geographic Location:** The patient is a resident of one of 13 specified counties in Northern California surrounding the clinic location.
**2. Referral Source:** The patient has a referral from a board-certified pediatrician, family practice physician, developmental pediatrician, neurologist, psychiatrist, psychologist, or equivalent.
**3. Medical Records:** A minimum of three years of medical records preceding the symptomatic deterioration are available during the screening process. If the patient is accepted, all medical records from birth to the present are required.
**4. Primary Medical Doctor (PMD) Relationship:** The patient has an established relationship with a local PMD who has clearly documented a stable functional baseline prior to the deterioration. Preference is given to patients who were followed by the same clinic for at least 3 years prior to the deterioration so that the prior baseline is known to the primary practice.
**5. Record Review Clarification:** In cases where a review of records is inconclusive, the PMD is available for a case consultation with the screening nurse practitioner to assess the likelihood of Pediatric Acute-onset Neuropsychiatric Syndrome (PANS).
**6. Comorbid Conditions:** Preexisting, stable diagnoses such as Autism Spectrum Disorder (ASD), Attention-Deficit/Hyperactivity Disorder (ADHD), Obsessive-Compulsive Disorder (OCD), or an anxiety disorder are permissible, provided the patient demonstrates stable baseline functioning (e.g., attending school) prior to the sudden onset of new psychiatric/behavioral symptoms.
**7. Symptom Onset:** The symptom onset is hyperacute (a transition from a stable baseline to severe symptoms in less than 72 hours) or acute (a transition from a stable baseline to severe symptoms within 3-7 days).
**8. Clinical Course and Timing:** Preference is given to patients with a single episode or a relapsing-remitting course who are in a flare for less than four months. As clinic capacity permits, patients up to nine months from the start of a flare are considered.
**9. Diagnostic Probability:** Based on a record review and potential discussion with the PMD, there is a high likelihood that the patient fulfills PANS criteria. The PMD, referring clinician(s), and the screening nurse practitioner must all have a strong clinical suspicion for PANS.
**10. Prior Treatment Status:** The patient is relatively untreated. The following prior treatments are permitted: a course of antibiotics for an infection (e.g., streptococcal pharyngitis) and standard-of-care medications for psychiatric symptoms.
**11. Vaccination Status:** The patient has completed all age-appropriate vaccinations as recommended by the Centers for Disease Control and Prevention (CDC) and the American Academy of Pediatrics (AAP). This prerequisite mitigates risks associated with possible immunomodulatory therapies and prevents exposure to other immunocompromised patients in the shared waiting room.

#### Notes on Protocol Implementation

##### Flexibility

During periods of low referral volume (such as summer months or the 2020-2021 quarantine period), the protocol allows for the inclusion of patients who are missing one or two of the above criteria.

##### Education

To improve the referral pipeline, the program provides educational outreach on PANS to local PMDs via grand rounds, lectures, and telephone consultations.

##### Protocol History

This standardized screening protocol has been in effect since September 2014. Between September 2012 and September 2014, all referred patients were accepted into the clinic. Prior to June 15, 2022, the geographic location criterion was restricted to 8 surrounding counties.

**Supplementary Figure S1.**
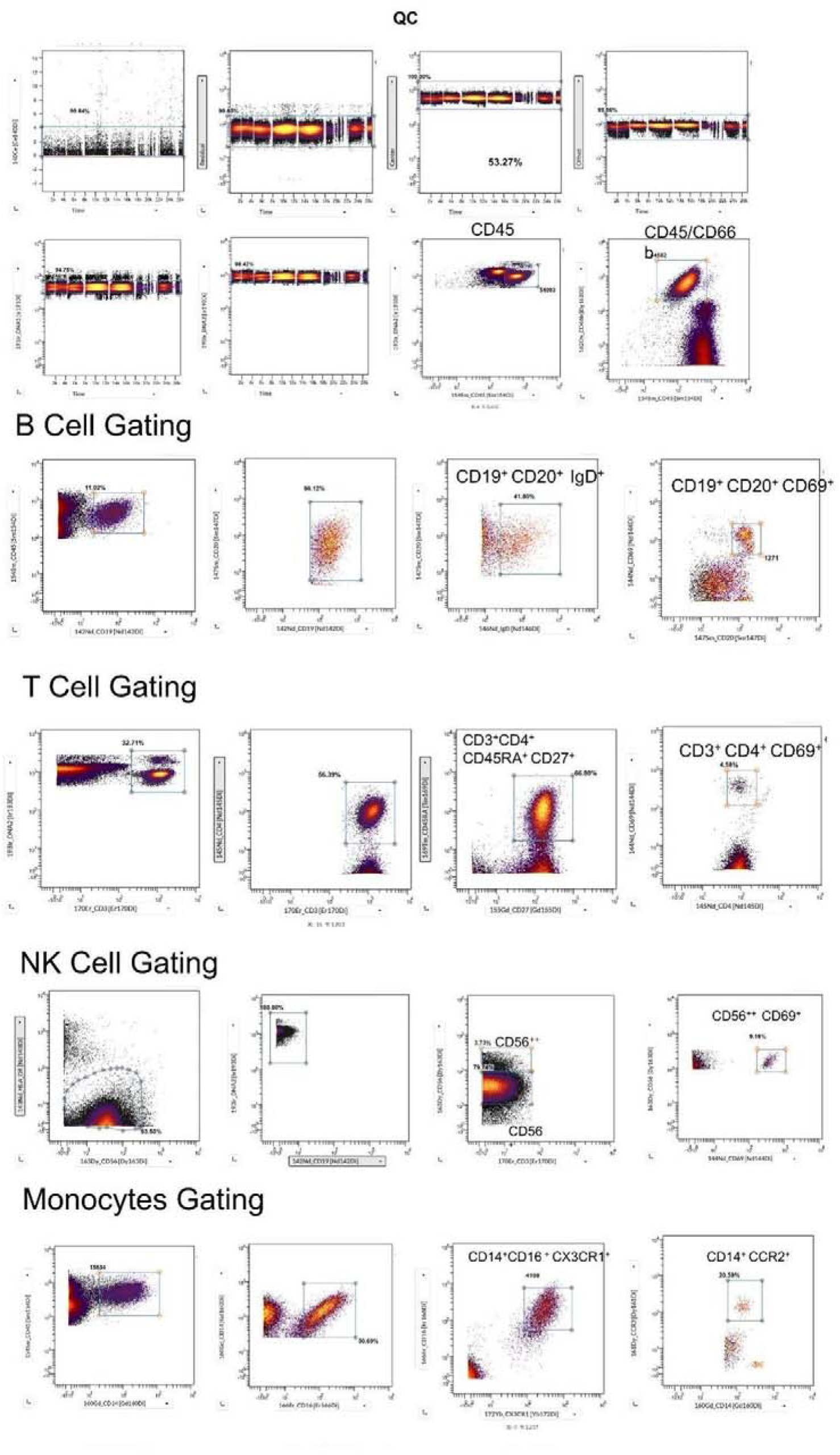
Gating strategy for mass cytometry data. Representative gating from one sample, done in Cytobank. (A) Quality control. Normalization beads were excluded at 140Ce versus time. Single intacT-cells were then selected using the Gaussian discrimination parameters (Residual, Center, and Offset) versus time, followed by DNA-positive events on the iridium intercalator channels (191Ir and 193Ir). CD45+ leukocytes were then selected, followed by the CD45/CD66b gate. (B) B-cells. CD19+ cells were gated from CD45^+^ leukocytes, and CD19+CD20+ B-cells from those. Naïve B-cells were defined as CD19+CD20+IgD+ and activated B-cells as CD19+CD20+CD69+. (C) CD4+ T-cells. CD3+ T-cells were gated, followed by CD3+CD4+ T-cells. Naïve CD4+ T-cells were defined as CD3+CD4+CD45RA+CD27− and activated CD4+ T-cells as CD3+CD4+CD69+. (D) NK cells. HLA-DR+/low and CD19+ cells were selected, followed by CD3+ cells. These were divided into CD56^bright^ (CD56++) and CD56^dim^ (CD56+) NK cells. Activated NK cells were defined as CD56^bright^CD69+. (E) Monocytes. CD14+ monocytes were gated from CD45+ leukocytes and resolved by CD14 and CD16 expression. Non-classical monocytes were defined as CD14+CD16+CX3CR1+ and classical monocytes as CD14+CCR2+. Numbers on the plots show the percentage of the parent population or the number of events in each gate. Axes show arcsinh-transformed signal intensity.

**Supplementary Figure S2.**
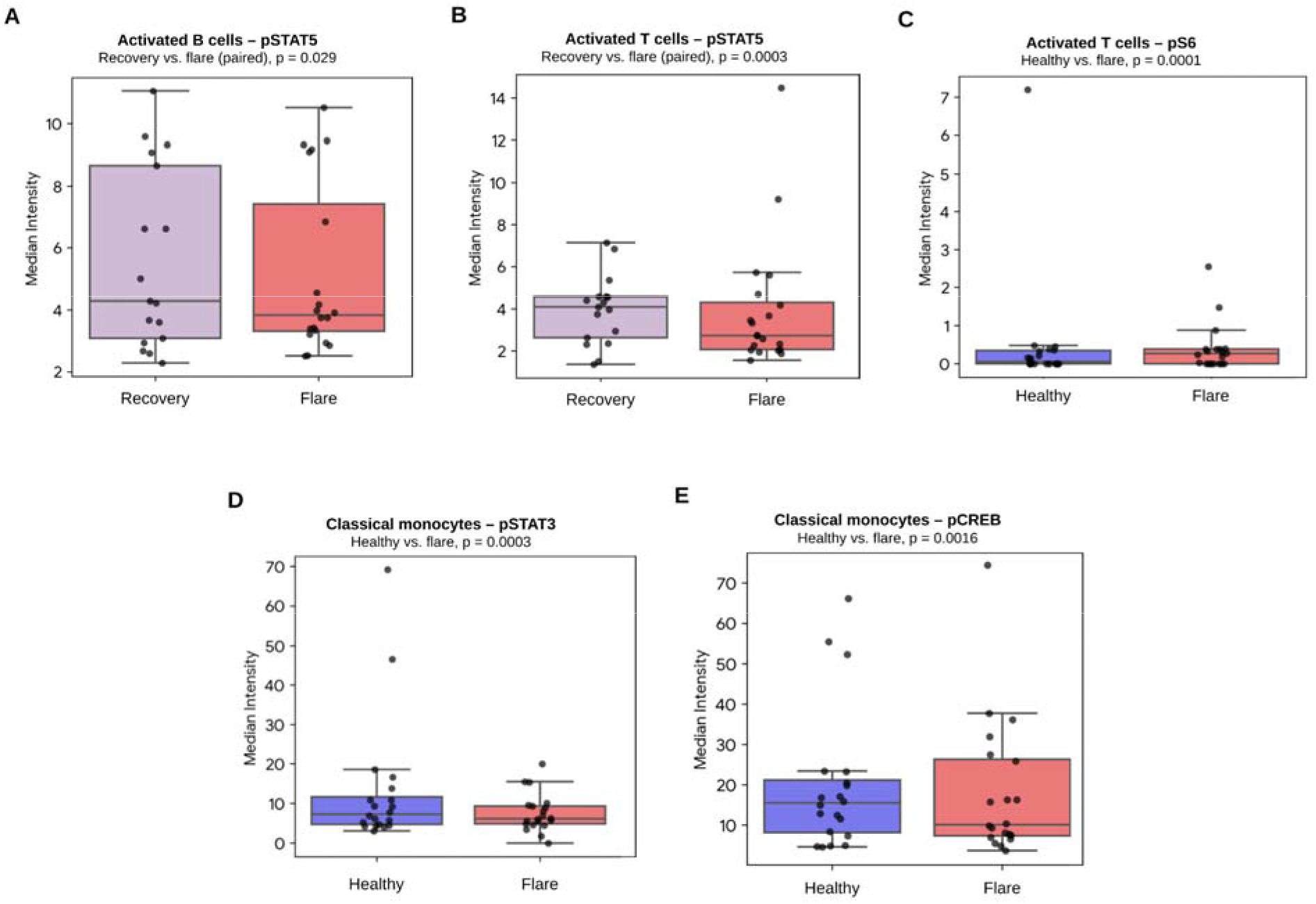
Exploratory phospho-signaling differences across immune-cell populations in PANS. Median intensity of selected phospho-proteins in (A) activated B-cells (pSTAT5; recovery vs. flare), activated T-cells showing (B) pSTAT5 (recovery vs. flare) and (C) pS6 (healthy vs. flare), classical monocytes showing (D) pSTAT3 and (E) pCREB (healthy vs. flare); Healthy controls are shown in blue, recovery in lavender, and flare in red. Points represent individual samples. Boxes show the median and interquartile range, and whiskers extend to 1.5 times the interquartile range. Paired comparisons used the Wilcoxon signed-rank test, and unpaired comparisons used the Mann-Whitney U test. P-values are nominal and not corrected for multiple comparisons.

**Supplementary Figure S3.**
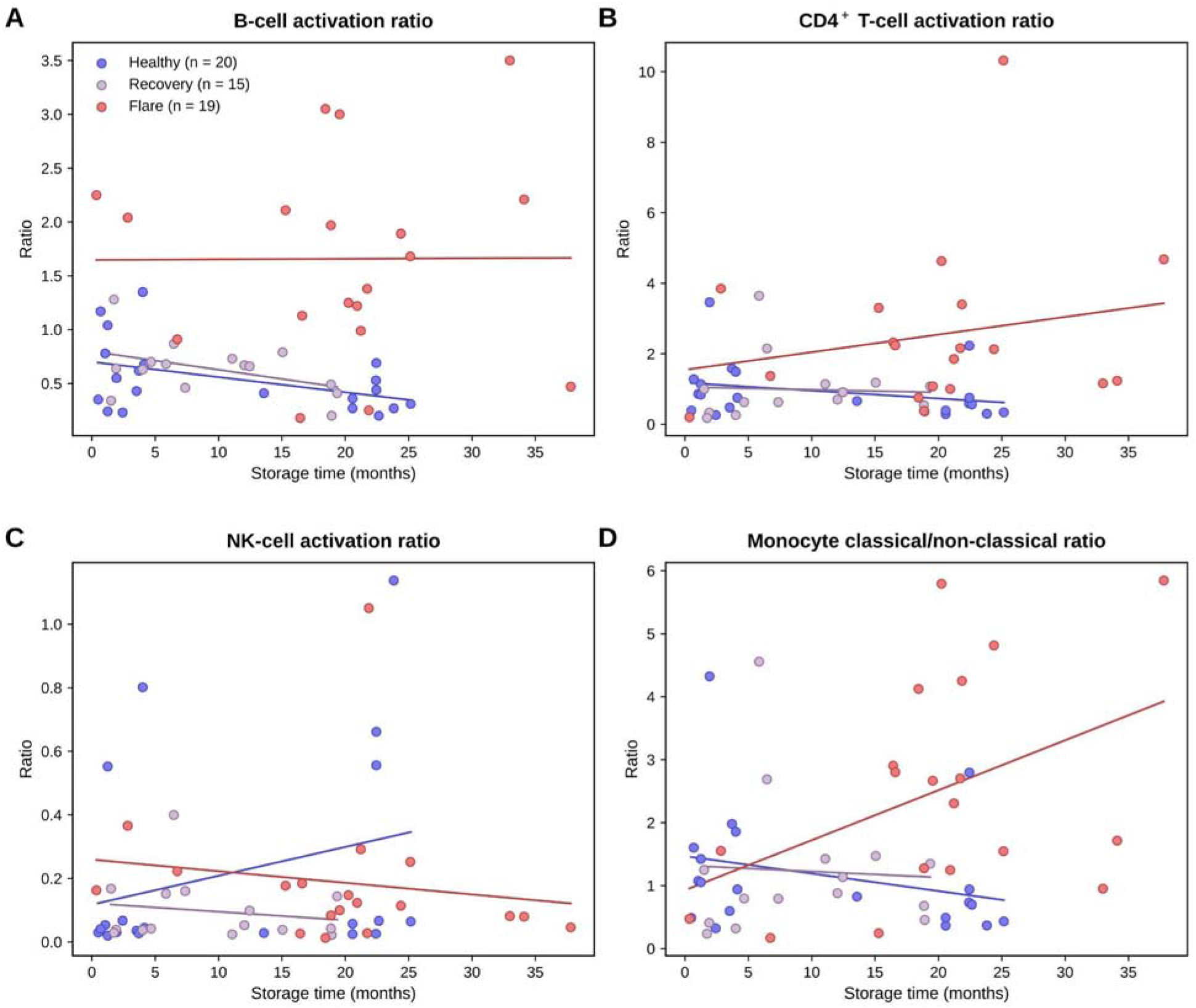
Sample storage time and immune-cell activation ratios. Storage time (months from blood draw to measurement on January 24, 2020) versus (A) B-cell activation ratio (CD19+CD20+CD69+ / CD19+CD20+IgD+CD27−), (B) CD4+ T-cell activation ratio (CD3+CD4+CD69+ / CD3+CD4+CD45RA+CD27−), (C) NK-cell activation ratio (CD56^bright^CD69+ / CD56^dim^), and (D) monocyte ratio (classical CD14+CD16+CCR2+ / non-classical CD14+CD16+CX3CR1+). Samples are from healthy controls (n = 20, blue), patients with PANS during recovery (n = 15, lavender), and patients with PANS during flare (n = 19, red). Storage time was not associated with any activation ratio within any group (Spearman correlation; healthy, recovery, and flare, respectively): (A) ρ = −0.34 (p = 0.14), −0.26 (p = 0.35), −0.06 (p = 0.79); (B) ρ = −0.33 (p = 0.16), 0.25 (p = 0.36), 0.23 (p = 0.34); (C) ρ = 0.36 (p = 0.12), −0.10 (p = 0.71), −0.24 (p = 0.32); (D) ρ = −0.31 (p = 0.18), 0.25 (p = 0.38), 0.39 (p = 0.10). Median storage time was 4.1 months (range, 0.5–25.2) for healthy controls, 7.4 months (range, 1.5–19.4) for recovery, and 20.2 months (range, 0.4–37.8) for flare. Each point represents one sample, and lines show linear fits within each group. Storage time was not associated with any activation ratio within any group (all p > 0.05). Each point represents one sample, and lines show linear fits within each group.

**Supplementary Table S1.** Patient Demographics and Clinical Characteristics at IBH Clinic Presentation.

|  | PANS (N=19) |  | Controls (N=20) |  |
|---|---|---|---|---|
| **Male, No. (%)** | 11 | 58% | 12 | 60% |
| **Race/Ethnicity, No. (%)** |  |  |  |  |
| White or European | 14 | 74% | 12 | 60% |
| Asian or Asian American | 1 | 5% | 2 | 10% |
| Hispanic or Latino (may include White) | 2 | 11% | 1 | 5% |
| Multiracial or Other | 2 | 11% | 5 | 25% |
| **Age at draw (first), mean (SD), years** | 8.77 | 2.34 | 9.51 | 2.42 |
| **Age at first-ever neuropsychiatric deterioration, mean (SD), years** | 7.57 | 2.16 |  |  |
| **Age at neuropsychiatric deterioration prompting referral to clinic, mean (SD), years** | 8.30 | 2.20 |  |  |
| **Age at PANS onset, mean (SD), years** | 7.93 | 2.37 |  |  |
| **Age at IBH Clinic presentation, mean (SD), years** | 8.52 | 2.24 |  |  |
| **Meets PANS criteria at or before IBH Clinic presentation, No. (%)** | 17 | 89% |  |  |
| **Global Impairment at intake, mean (SD)** | 46.47 | 20.04 |  |  |
| **CGI at intake, mean (SD)** | 35.11 | 14.62 |  |  |
| **WPI at intake, mean (SD)** | 2.68 | 2.96 |  |  |
| **SSS at intake, mean (SD)** | 4.79 | 3.08 |  |  |
| **CGAS at intake, mean (SD)** | 58.47 | 14.94 |  |  |
| **Percent Baseline at intake, mean (SD)** | 52.25 | 24.58 |  |  |
| Scores reflect first score available within 90 days of intake |  |  |  |  |
| **31-Item Symptoms at Intake, No. (%)** |  |  |  |  |
| Obsessions | 18 | 95% |  |  |
| Compulsions | 15 | 79% |  |  |
| Hoarding | 5 | 26% |  |  |
| Food refusal or avoidance | 8 | 42% |  |  |
| Urge to overeat; thinking about eating all the time | 2 | 11% |  |  |
| Fluid refusal or avoidance | 1 | 5% |  |  |
| Separation anxiety | 12 | 63% |  |  |
| Other anxiety, fears, phobias, or panic attacks | 14 | 74% |  |  |
| Mood swings or moodiness | 17 | 89% |  |  |
| Emotional lability | 9 | 47% |  |  |
| Suicidal ideation or behavior | 8 | 42% |  |  |
| Depression or sadness | 12 | 63% |  |  |
| Irritability | 16 | 84% |  |  |
| Aggressive behaviors, violence, or rage | 12 | 63% |  |  |
| Oppositional behaviors | 14 | 74% |  |  |
| Hyperactivity or impulsivity | 8 | 42% |  |  |
| Trouble paying attention | 11 | 58% |  |  |
| Baby talk | 3 | 16% |  |  |
| Other behavioral or developmental regression (e.g., poor self care, immature judgement for age) | 4 | 21% |  |  |
| Worsening of school performance | 6 | 32% |  |  |
| Worsening of handwriting, copying, or art | 2 | 11% |  |  |
| Cognitive symptoms (difficulty thinking, foggy brain, memory problems) | 7 | 37% |  |  |
| Pain (headaches, abdominal pain, body pain) | 13 | 68% |  |  |
| Sleep disturbance | 14 | 74% |  |  |
| Daytime wetting or bedwetting (enuresis) | 8 | 42% |  |  |
| Urinary frequency (uses restroom frequently) | 3 | 16% |  |  |
| Bothered by sounds, smells, textures, or lights (sensory amplification) | 12 | 63% |  |  |
| Hallucinations | 4 | 21% |  |  |
| Delusions or paranoid thoughts | 5 | 26% |  |  |
| Motor tics | 6 | 32% |  |  |
| Phonic or vocal tics | 5 | 26% |  |  |

**Supplementary Table S2.** Mass cytometry antibody panel.

| Metal | Target | Clone | Source | Catalog no. | Lot / conjugation date | Final concentration | Staining |
|---|---|---|---|---|---|---|---|
| 141Pr | CD7 | CD7-6B7 | In-house conjugate | — | 4/3/2018 | 10 µg/mL | Surface |
| 142Nd | CD19 | HIB19 | Standard BioTools | 3142001B | 2221601 | 0.5 µL/test | Surface |
| 143Nd | HLA-DR | L243 | Standard BioTools | 3143013B | — | 0.5 µL/test | Surface |
| 144Nd | CD69 | FN50 | Standard BioTools | 3144018B | — | 0.5 µL/test | Surface |
| 145Nd | CD4 | RPA-T4 | Standard BioTools | 3145001B | 2171601 | 0.5 µL/test | Surface |
| 146Nd | IgD | IA6-2 | Standard BioTools | 3146005B | 1941604 | 0.25 µL/test | Surface |
| 147Sm | CD20 | 2H7 | Standard BioTools | 3147001B | 2741602 | 1 µL/test | Surface |
| 148Nd | pSTAT4 | 38 | Standard BioTools | 3148006A | — | [ ] | Intracellular |
| 149Sm | CD25 (IL-2R) | 2A3 | Standard BioTools | 3149010B | 1271502 | 0.5 µL/test | Surface |
| 150Nd | pSTAT5 | 47 | Standard BioTools | 3150005A | — | [ ] | Intracellular |
| 151Eu | CD123 (IL-3R) | 6H6 | Standard BioTools | 3151001B | 961508 | 0.25 µL/test | Surface |
| 152Sm | γδ TCR | 11F2 | Standard BioTools | 3152008B | 3261605 | 0.5 µL/test | Surface |
| 153Eu | pSTAT1 | 4a | Standard BioTools | 3153005A | — | [ ] | Intracellular |
| 154Sm | CD45 | HI30 | Standard BioTools | 3154001B | — | 0.25 µL/test | Surface |
| 155Gd | CD27 | L128 | Standard BioTools | 3155001B | 931512 | 0.5 µL/test | Surface |
| 156Gd | p-p38 | D3F9 | Standard BioTools | 3156002A | — | [ ] | Intracellular |
| 157Gd | CD24 | ML-5 | In-house conjugate | — | 10/10/2017 | 10 µg/mL | Surface |
| 158Gd | pSTAT3 | 4 | Standard BioTools | 3158005A | — | [ ] | Intracellular |
| 159Tb | CD11c | Bu15 | Standard BioTools | 3159001B | 2591516 | 0.5 µL/test | Surface |
| 160Gd | CD14 | M5E2 | Standard BioTools | 3160001B | 1481502 | 1 µL/test | Surface |
| 161Dy | CCR2 | K036C2 | BioLegend | 357202B | — | 2.5 µg/mL | Surface |
| 162Dy | CD66b | 80H3 | Standard BioTools | 3162023B | — | 0.25 µL/test | Surface |
| 163Dy | CD56 (NCAM) | NCAM16.2 | Standard BioTools | 3163007B | 131506 | 0.5 µL/test | Surface |
| 164Dy | IgM | MHM-88 | BioLegend | 314527B | 1741606 | 2.5 µg/mL | Surface |
| 165Ho | pCREB | 87G3 | Standard BioTools | 3165009A | — | [ ] | Intracellular |
| 166Er | CD16 | B73.1 | eBioscience | 16-0167-85 | 3/1/2017 | 10 µg/mL | Surface |
| 167Er | CD38 | HIT2 | Standard BioTools | 3167001B | 1031705 | 0.5 µL/test | Surface |
| 168Er | CD8a | SK1 | Standard BioTools | 3168002B | 3421401 | 0.5 µL/test | Surface |
| 169Tm | CD45RA | HI100 | Standard BioTools | 3169008B | 1581616 | 0.5 µL/test | Surface |
| 170Er | CD3 | UCHT1 | Standard BioTools | 3170001B | 2581608 | 0.5 µL/test | Surface |
| 171Yb | pERK1/2 | D13.14.4E | Standard BioTools | 3171010A | — | [ ] | Intracellular |
| 172Yb | CX3CR1 | 2A9-1 | Standard BioTools | 3172017B | — | [ ] | Surface |
| 173Yb | KIR3DL1 | DX9 | BioLegend | 330102 | 12/2/2019 | [5 or 10] µg/mL | Surface |
| 174Yb | CD49d | 9F10 | Standard BioTools | 3174018B | 1671714 | 0.25 µL/test | Surface |
| 175Lu | pS6 | N7-548 | Standard BioTools | 3175009A | — | [ ] | Intracellular |
| 176Yb | CD127 (IL-7Rα) | [ ] | [ ] | — | — | 5 µg/mL | Surface |
| 209Bi | CD11b | ICRF44 | Standard BioTools | 3209003B | — | 0.5 µL/test | Surface |

**Supplementary Table S3.** Immune-cell ratios and secondary clinical outcomes.

| Outcome | Effect | Estimate | 95% CI | P value | FDR-adjusted P | Samples, n | Participants, n |
|---|---|---|---|---|---|---|---|
| **B-cell** |  |  |  |  |  |  |  |
| CY-BOCS total | Beta | 2.908 | -0.375 to 6.191 | 0.119 | 0.477 | 23 | 16 |
| YGTSS total | Beta | 5.871 | -3.744 to 15.485 | 0.257 | 0.513 | 13 | 11 |
| MOAS total | Beta | 2.097 | -0.121 to 4.314 | 0.084 | 0.205 | 34 | 19 |
| Columbia Impairment total | Beta | 4.318 | 0.567 to 8.069 | 0.040 | 0.161 | 28 | 18 |
| MOAS log(MOAS + 0.1) | Beta | 0.466 | -0.302 to 1.233 | 0.248 | 0.272 | 34 | 19 |
| Any aggression (MOAS > 0) | OR | 1.565 | 0.652 to 3.754 | 0.316 | 0.450 | 34 | 19 |
| **T-cell** |  |  |  |  |  |  |  |
| CY-BOCS total | Beta | 1.305 | -1.376 to 3.987 | 0.353 | 0.625 | 23 | 16 |
| YGTSS total | Beta | -0.634 | -6.746 to 5.477 | 0.843 | 0.843 | 13 | 11 |
| MOAS total | Beta | 1.085 | -0.696 to 2.866 | 0.244 | 0.244 | 34 | 19 |
| Columbia Impairment total | Beta | 0.359 | -2.537 to 3.254 | 0.810 | 0.810 | 28 | 18 |
| MOAS log(MOAS + 0.1) | Beta | 0.326 | -0.216 to 0.868 | 0.248 | 0.272 | 34 | 19 |
| Any aggression (MOAS > 0) | OR | 1.347 | 0.733 to 2.475 | 0.338 | 0.450 | 34 | 19 |
| **NK-cell** |  |  |  |  |  |  |  |
| CY-BOCS total | Beta | -4.723 | -20.107 to 10.662 | 0.557 | 0.625 | 23 | 16 |
| YGTSS total | Beta | -20.028 | -49.092 to 9.036 | 0.204 | 0.513 | 13 | 11 |
| MOAS total | Beta | 10.213 | -1.548 to 21.974 | 0.105 | 0.205 | 34 | 19 |
| Columbia Impairment total | Beta | -3.278 | -22.839 to 16.282 | 0.745 | 0.810 | 28 | 18 |
| MOAS log(MOAS + 0.1) | Beta | 3.318 | -0.363 to 7.000 | 0.088 | 0.272 | 34 | 19 |
| Any aggression (MOAS > 0) | OR | 715.589 | 0.009 to 55984478.669 | 0.253 | 0.450 | 34 | 19 |
| **Monocyte** |  |  |  |  |  |  |  |
| CY-BOCS total | Beta | 0.566 | -1.666 to 2.798 | 0.625 | 0.625 | 23 | 16 |
| YGTSS total | Beta | 1.363 | -3.461 to 6.188 | 0.591 | 0.788 | 13 | 11 |
| MOAS total | Beta | 1.115 | -0.377 to 2.607 | 0.154 | 0.205 | 34 | 19 |
| Columbia Impairment total | Beta | 1.582 | -0.864 to 4.028 | 0.217 | 0.433 | 28 | 18 |
| MOAS log(MOAS + 0.1) | Beta | 0.253 | -0.190 to 0.696 | 0.272 | 0.272 | 34 | 19 |
| Any aggression (MOAS > 0) | OR | 1.155 | 0.739 to 1.805 | 0.527 | 0.527 | 34 | 19 |

Beta estimates are per one-unit higher immune-cell ratio; MOAS log models report coefficients on the transformed scale. Odds ratios (OR) are shown for MOAS > 0. Approximate Wald 95% CIs are reported. Models include a participant random intercept. Benjamini–Hochberg FDR correction is within each outcome across the four immune-cell ratios.

**Supplementary Table S4.**
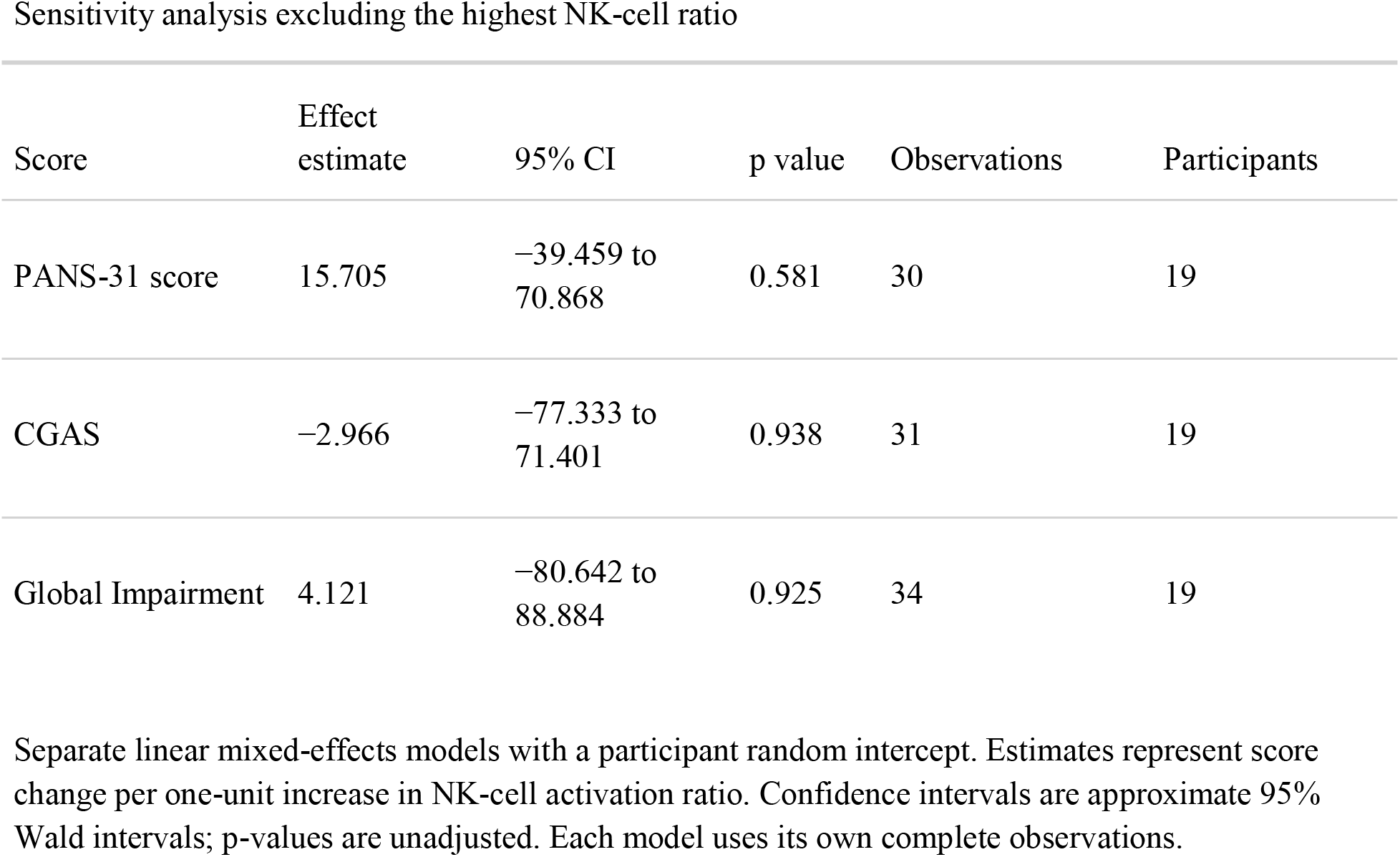
NK-cell activation ratio and psychiatric scores.

| Score | Effect estimate | 95% CI | p value | Observations | Participants |
|---|---|---|---|---|---|
| PANS-31 score | 15.705 | −39.459 to 70.868 | 0.581 | 30 | 19 |
| CGAS | −2.966 | −77.333 to 71.401 | 0.938 | 31 | 19 |
| Global Impairment | 4.121 | −80.642 to 88.884 | 0.925 | 34 | 19 |

Separate linear mixed-effects models with a participant random intercept. Estimates represent score change per one-unit increase in NK-cell activation ratio. Confidence intervals are approximate 95% Wald intervals; p-values are unadjusted. Each model uses its own complete observations.

**Supplementary Table S5.** Sensitivity analysis adjusting for flare status, immunomodulator use, and sex. Each covariate was included separately in linear mixed-effects models to assess the robustness of associations between immune-cell activation ratios and clinical severity.

| Outcome | Adjustment | Primary estimate (B cell ratio) | Adjusted estimate | Adjusted p value | % attenuation |
|---|---|---|---|---|---|
| **CGAS score** | **Flare status** | **-14.891** | **-9.371** | **0.010** | **37.1%** |
| CGAS score | Immunomodulator use | -16.597 | -16.054 | <0.001 | 3.3% |
| CGAS score | Sex | -14.891 | -14.566 | <0.001 | 2.2% |
| **Global impairment** | **Flare status** | **12.811** | **-0.663** | **0.847** | **94.8%** |
| Global impairment | Immunomodulator use | 14.236 | 12.594 | 0.003 | 11.5% |
| Global impairment | Sex | 12.811 | 12.715 | 0.004 | 0.7% |
| **PANS-31 score** | **Flare status** | **8.373** | **5.278** | **0.168** | **37.0%** |
| PANS-31 score | Immunomodulator use | 9.614 | 9.616 | 0.002 | 0.0% |
| PANS-31 score | Sex | 8.373 | 9.262 | 0.004 | -10.6% |

**Supplementary Table S6.**
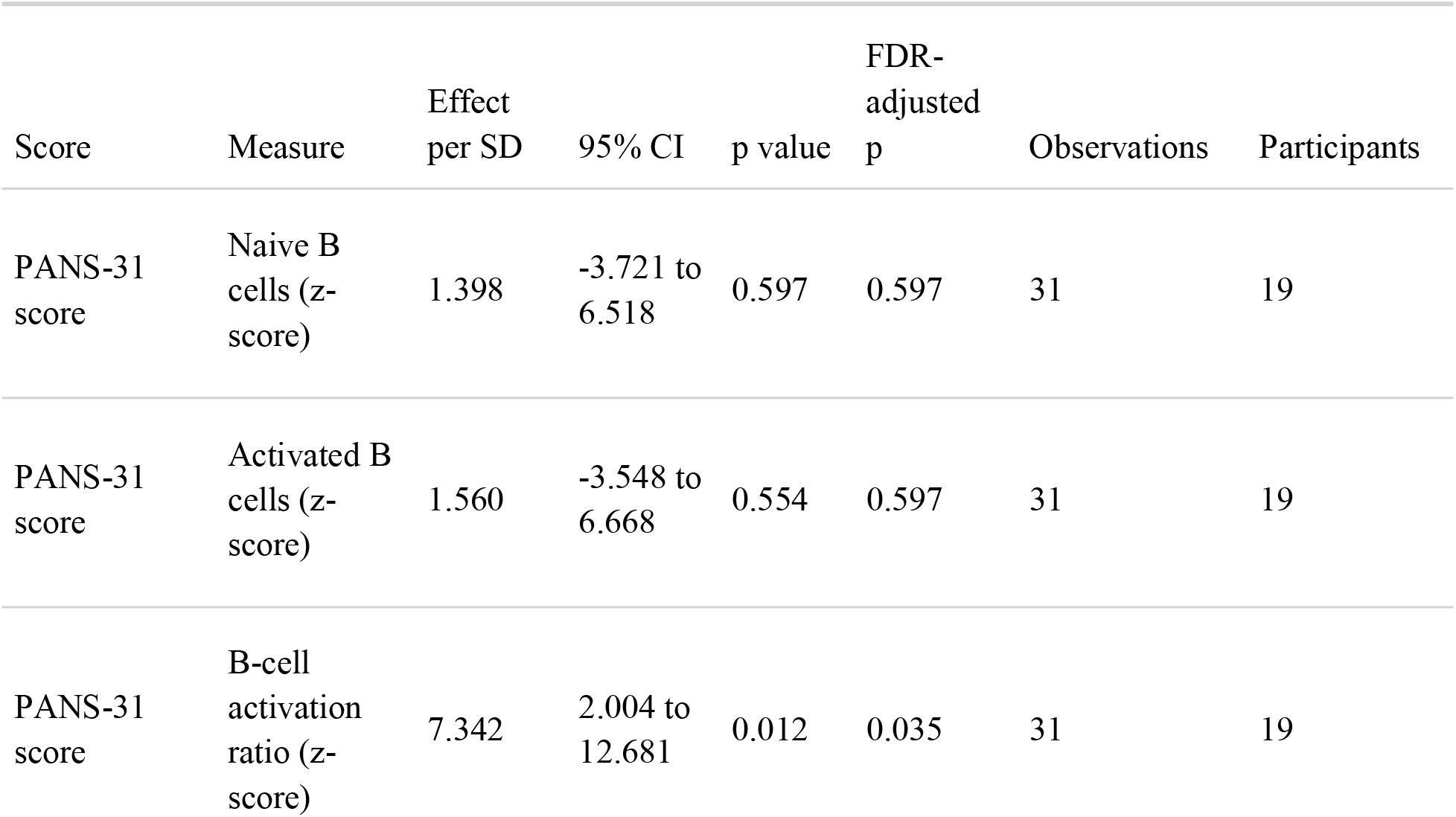
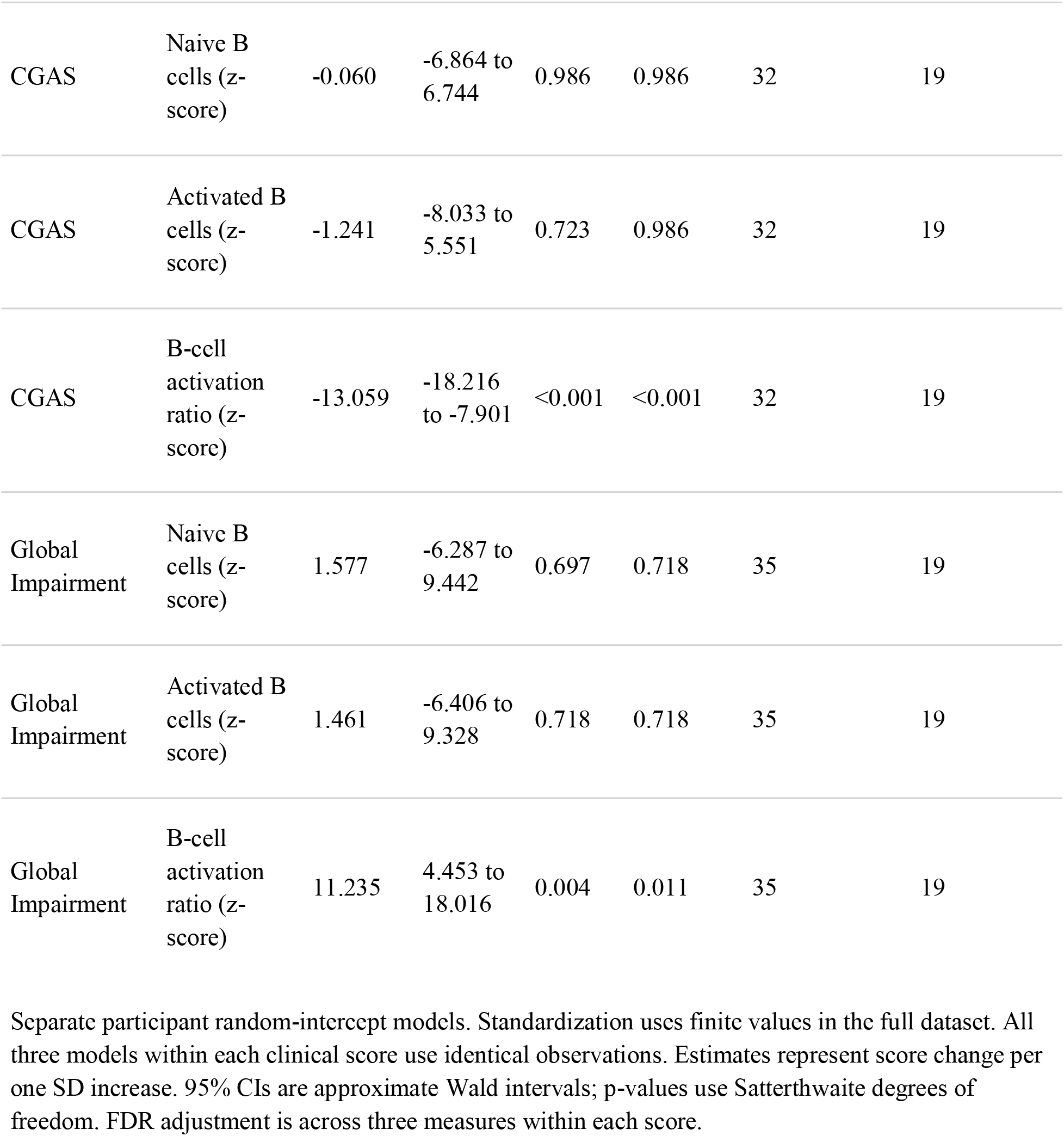
Standardized B-cell measures and clinical severity with outliers retained.

| Score | Measure | Effect per SD | 95% CI | p value | FDR-adjusted p | Observations | Participants |
|---|---|---|---|---|---|---|---|
| PANS-31 score | Naive B cells (z-score) | 1.398 | -3.721 to 6.518 | 0.597 | 0.597 | 31 | 19 |
| PANS-31 score | Activated B cells (z-score) | 1.560 | -3.548 to 6.668 | 0.554 | 0.597 | 31 | 19 |
| PANS-31 score | B-cell activation ratio (z-score) | 7.342 | 2.004 to 12.681 | 0.012 | 0.035 | 31 | 19 |
| CGAS | Naive B cells (z-score) | -0.060 | -6.864 to 6.744 | 0.986 | 0.986 | 32 | 19 |
| CGAS | Activated B cells (z-score) | -1.241 | -8.033 to 5.551 | 0.723 | 0.986 | 32 | 19 |
| CGAS | B-cell activation ratio (z-score) | -13.059 | -18.216 to -7.901 | <0.001 | <0.001 | 32 | 19 |
| Global Impairment | Naive B cells (z-score) | 1.577 | -6.287 to 9.442 | 0.697 | 0.718 | 35 | 19 |
| Global Impairment | Activated B cells (z-score) | 1.461 | -6.406 to 9.328 | 0.718 | 0.718 | 35 | 19 |
| Global Impairment | B-cell activation ratio (z-score) | 11.235 | 4.453 to 18.016 | 0.004 | 0.011 | 35 | 19 |

Separate participant random-intercept models. Standardization uses finite values in the full dataset. All three models within each clinical score use identical observations. Estimates represent score change per one SD increase. 95% CIs are approximate Wald intervals; p-values use Satterthwaite degrees of freedom. FDR adjustment is across three measures within each score.

**Supplementary Figure S4.**
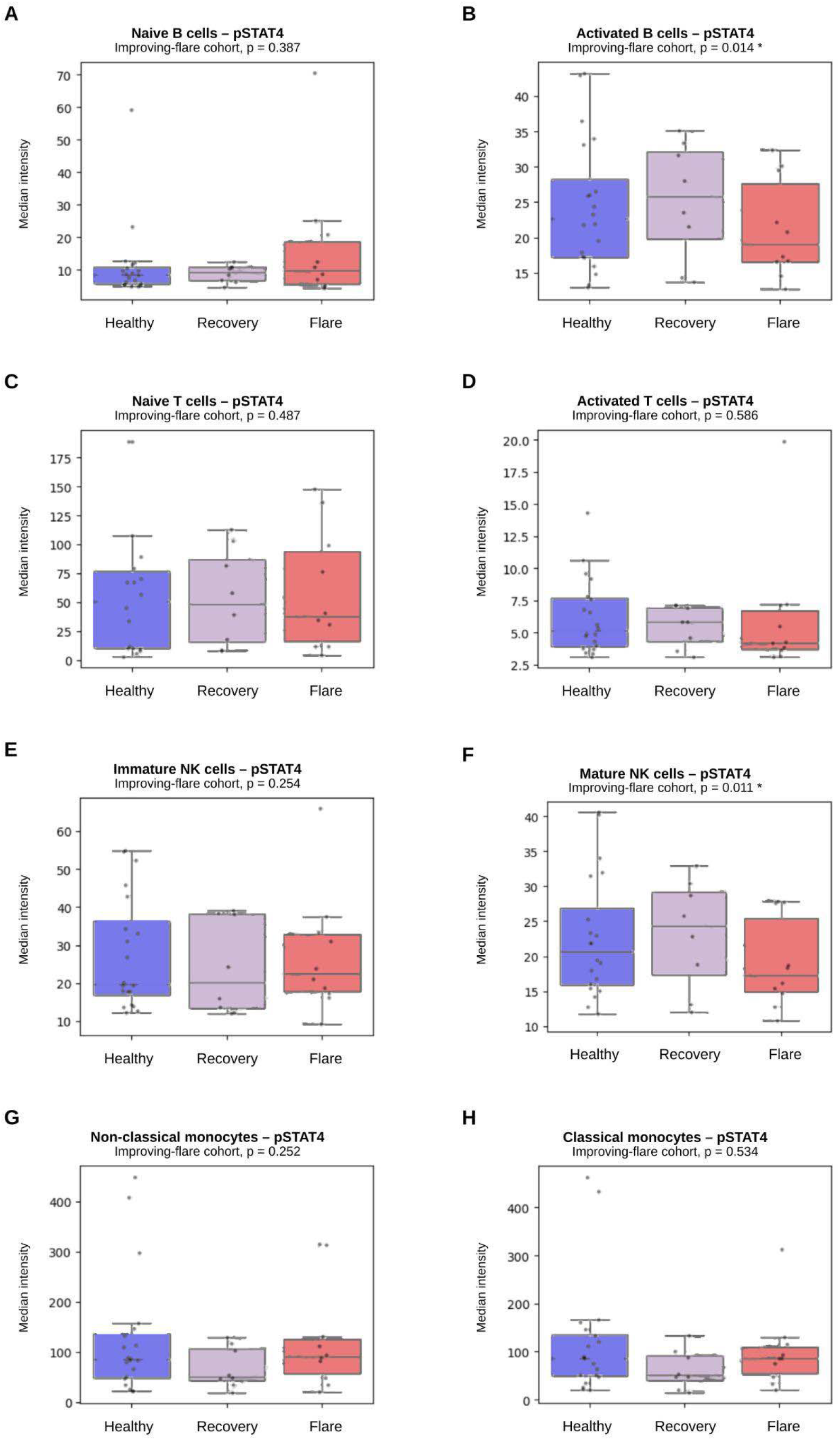
pSTAT4 signaling in the improving-flare cohort. Median intensity of pSTAT4 in healthy controls and in patients with PANS sampled during improving flare and subsequent recovery. Nominal differences were observed in activated B-cells (p = 0.014) and mature NK cells (p = 0.011)

**Supplementary Figure S5.**
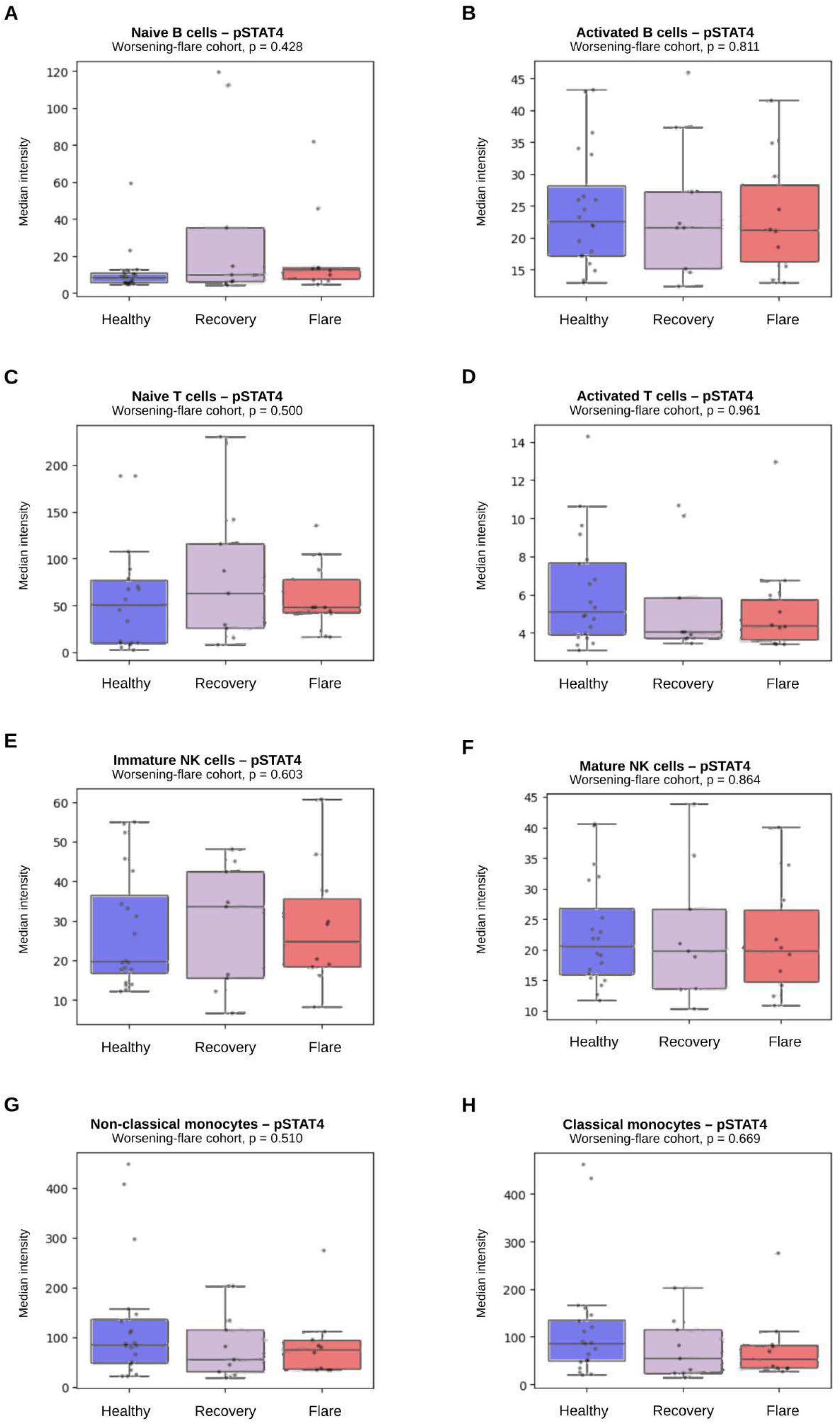
pSTAT4 signaling in the worsening-flare cohort. Median intensity of pSTAT4 in healthy controls and in patients with PANS sampled during worsening flare and subsequent recovery. No significant differences were observed.

**Supplementary Figure S6.**
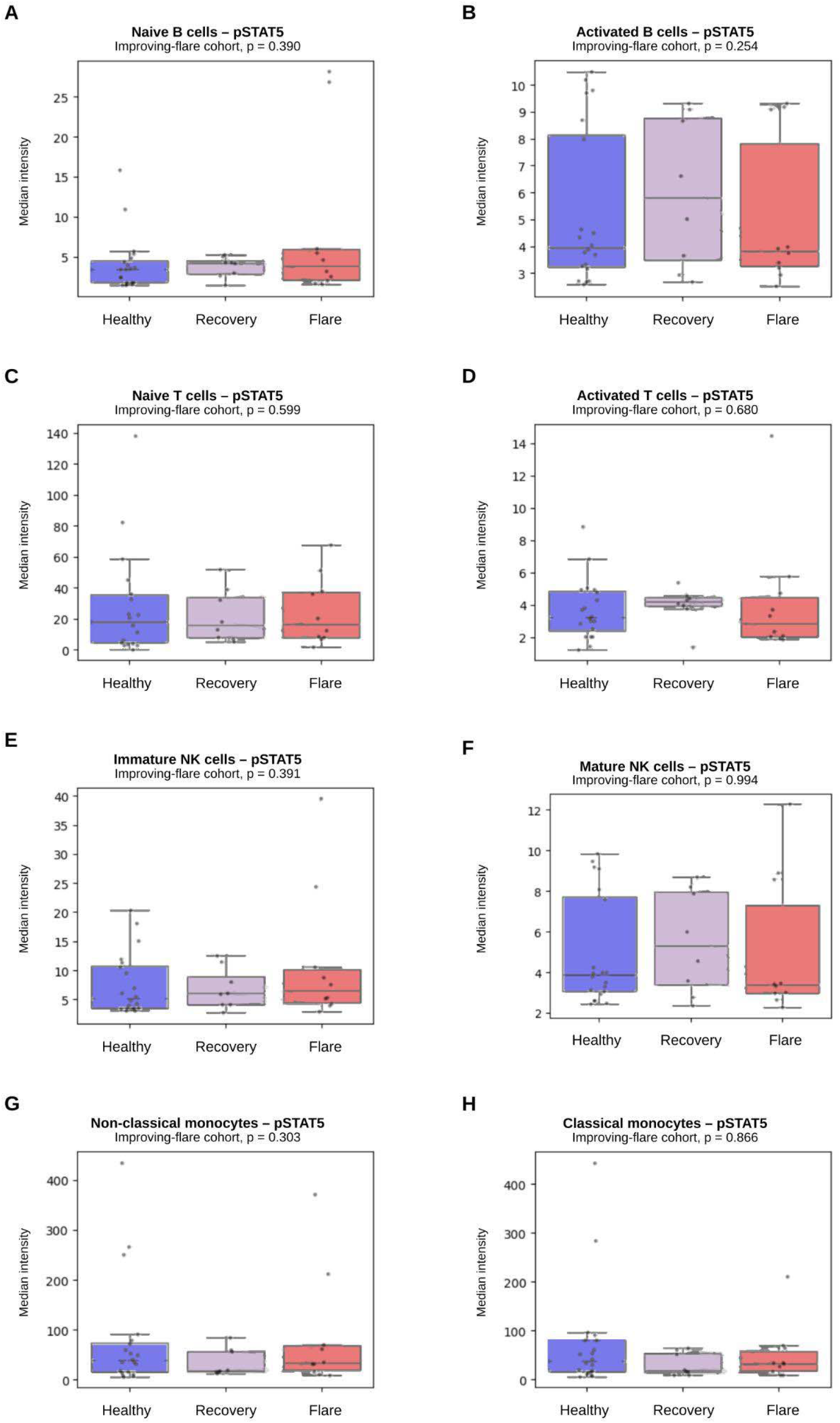
pSTAT5 signaling in the improving-flare cohort. Median intensity of pSTAT5 in healthy controls and in patients with PANS sampled during improving flare and subsequent recovery. No significant differences were observed.

**Supplementary Figure S7.**
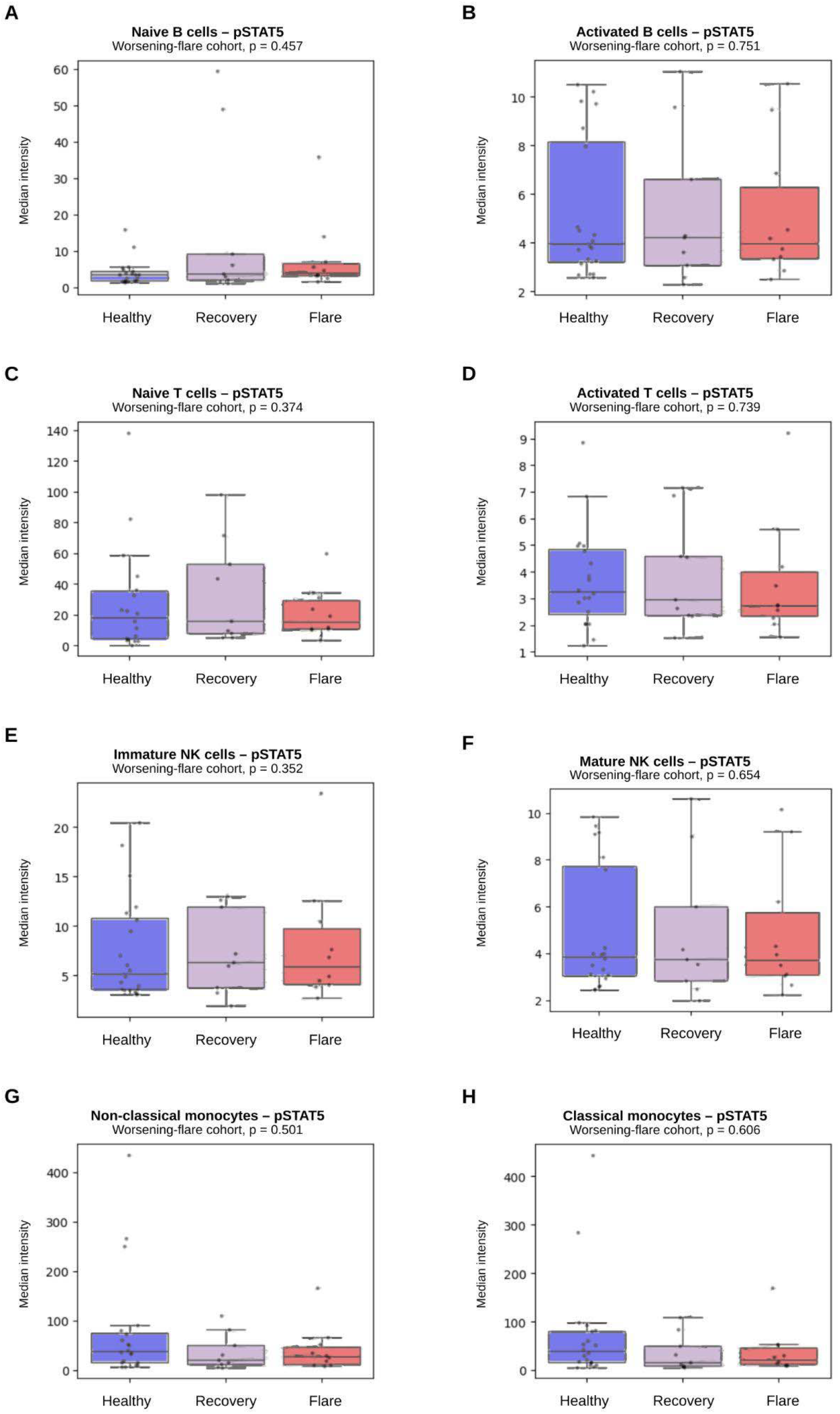
pSTAT5 signaling in the worsening-flare cohort. Median intensity of pSTAT5 in healthy controls and in patients with PANS sampled during worsening flare and subsequent recovery. No significant differences were observed.

**Supplementary Figure S8.**
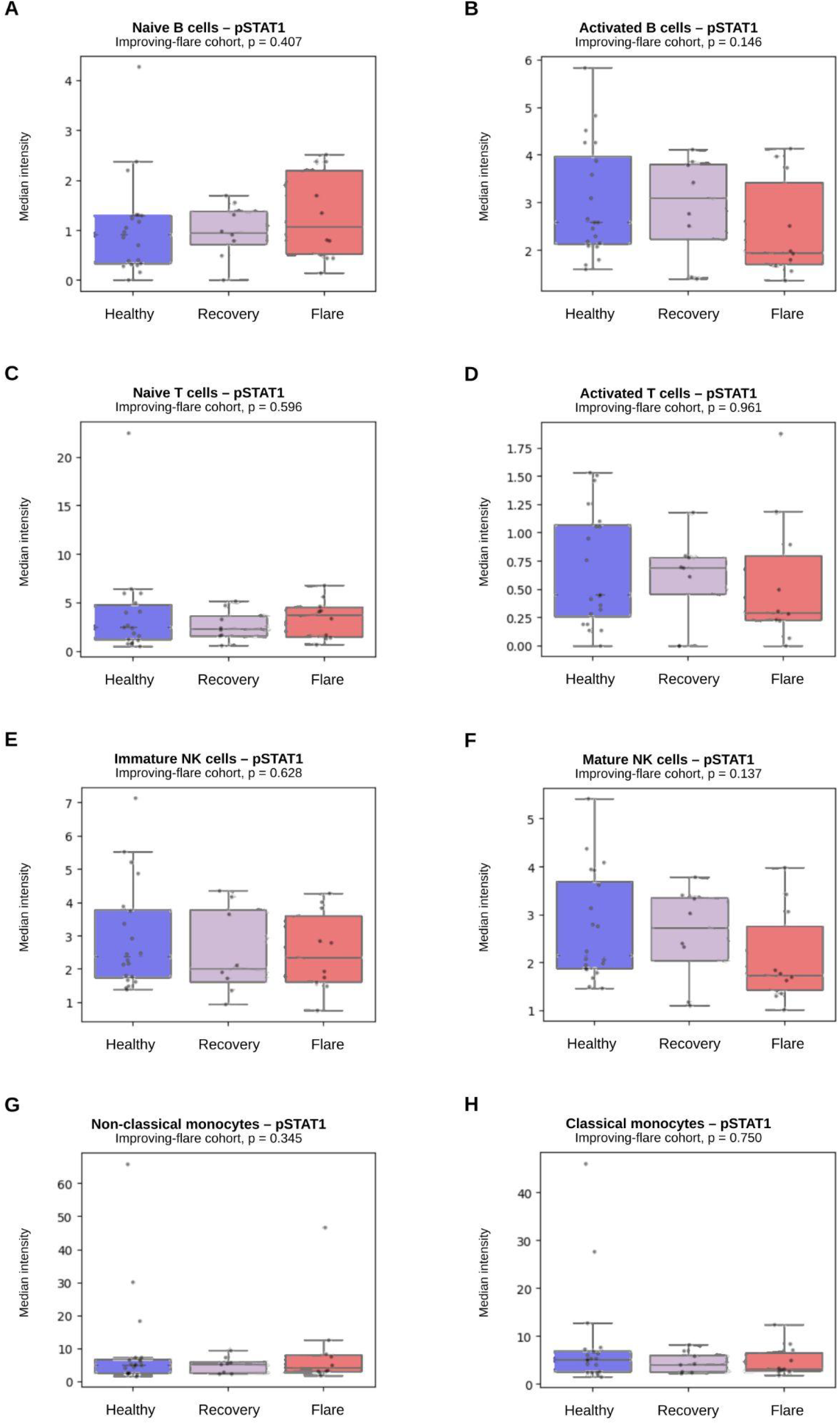
pSTAT1 signaling in the improving-flare cohort. Median intensity of pSTAT1 in healthy controls and in patients with PANS sampled during improving flare and subsequent recovery. No significant differences were observed

**Supplementary Figure S9.**
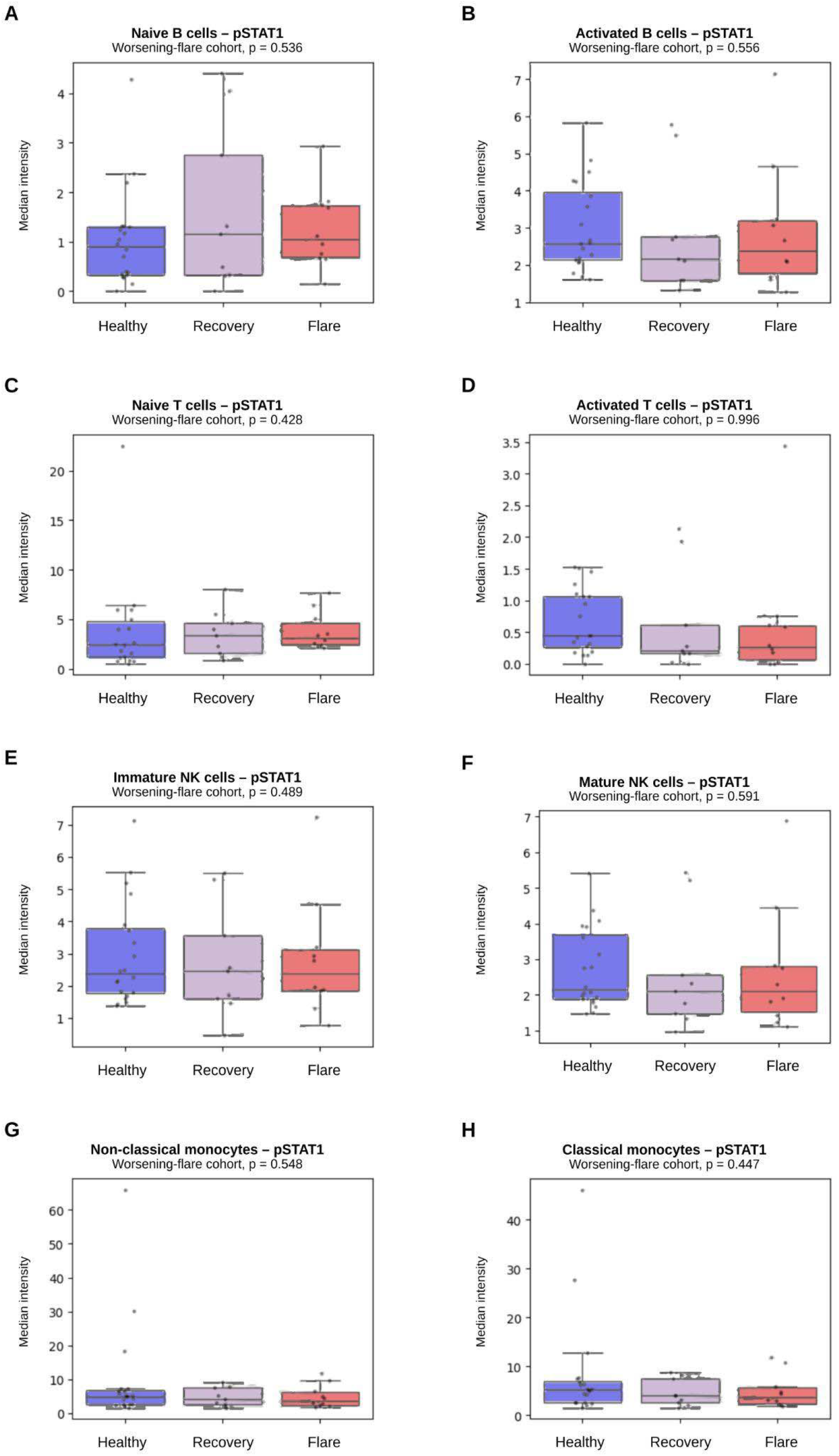
pSTAT1 signaling in the worsening-flare cohort. Median intensity of pSTAT1 in healthy controls and in patients with PANS sampled during worsening flare and subsequent recovery. No significant differences were observed.

**Supplementary Figure S10.**
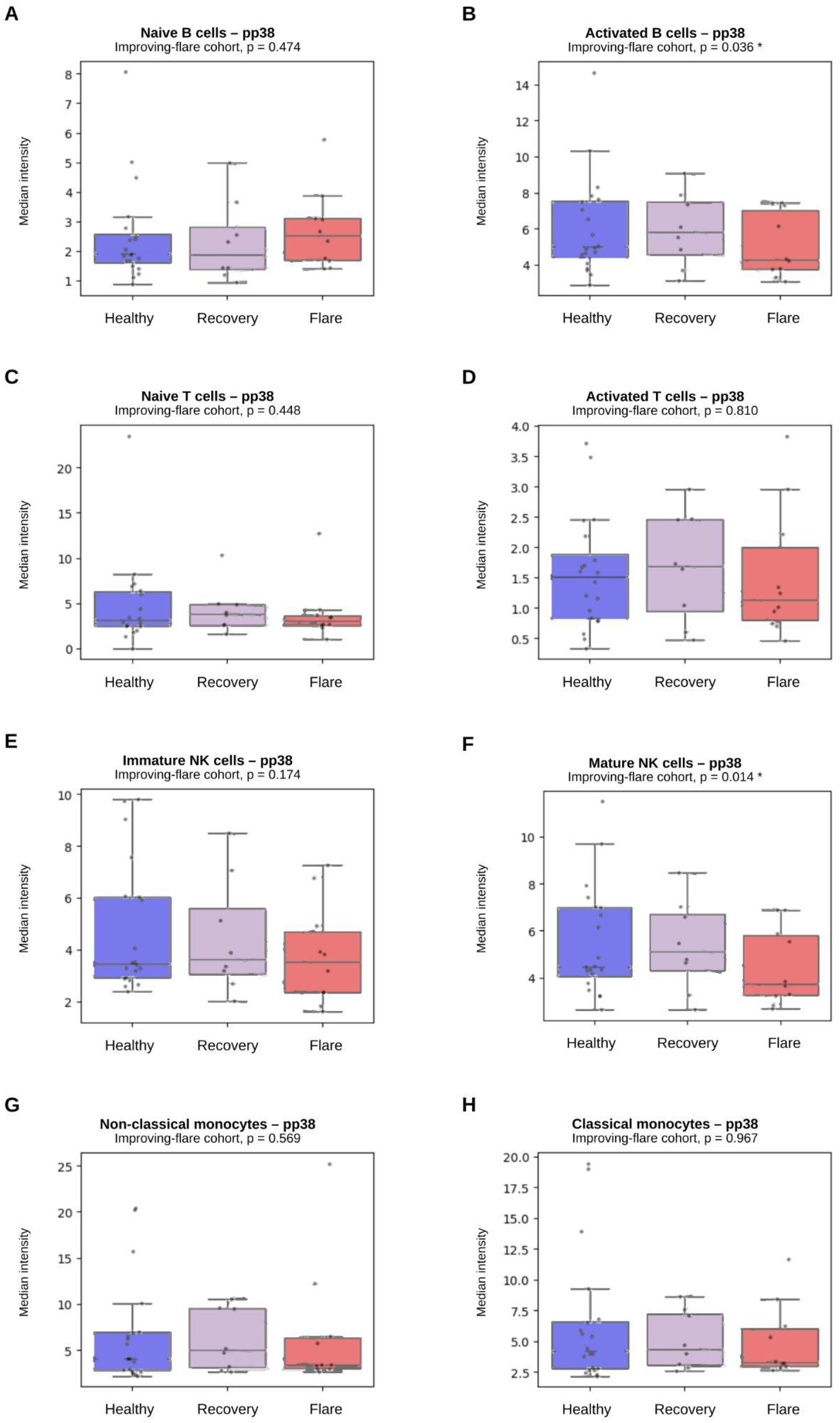
p38 MAPK (pp38) signaling in the improving-flare cohort. Median intensity of pp38 in healthy controls and in patients with PANS sampled during improving flare and subsequent recovery. Nominal differences were observed in activated B-cells (p = 0.036) and mature NK cells (p = 0.014).

**Supplementary Figure S11.**
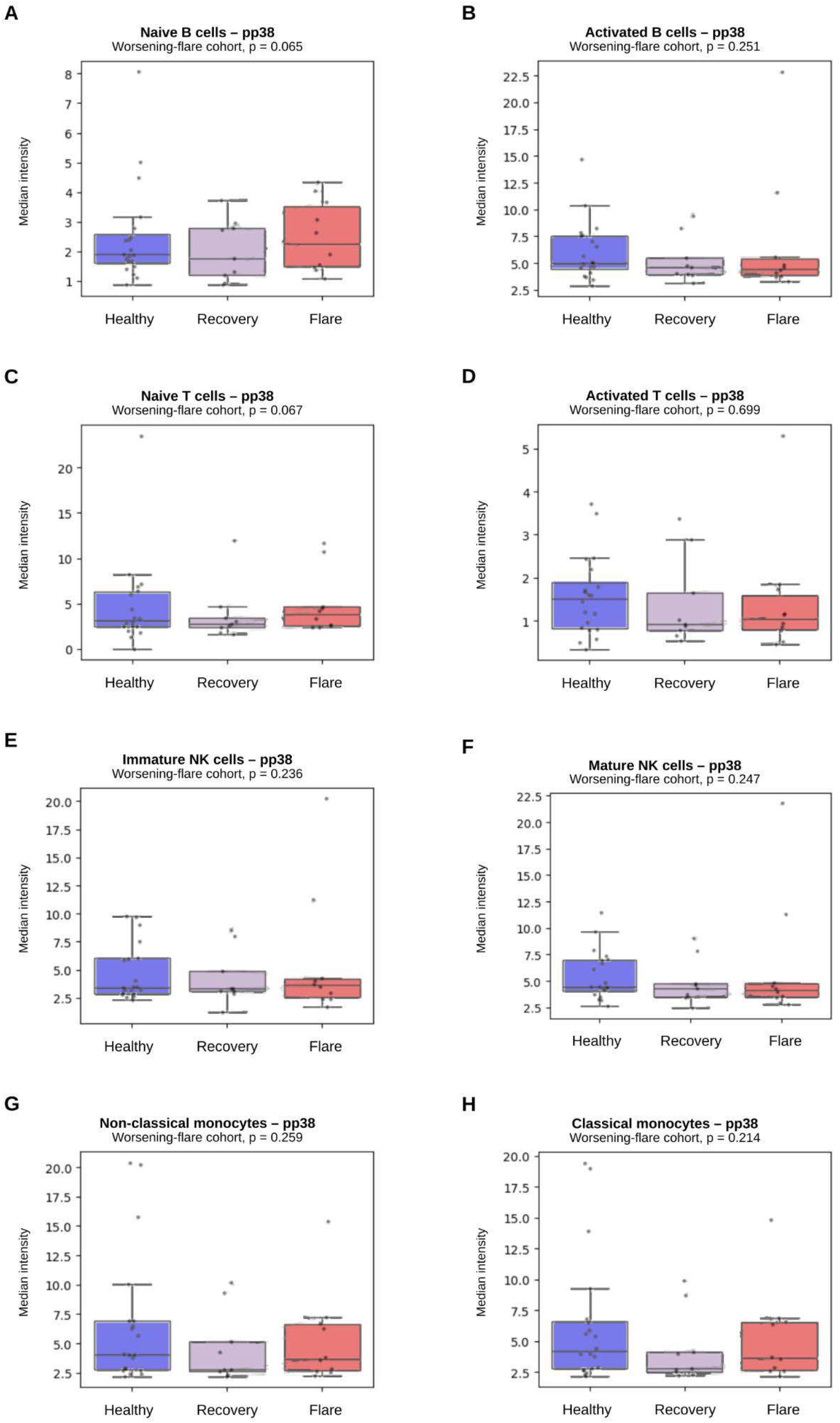
p38 MAPK (pp38) signaling in the worsening-flare cohort. Median intensity of pp38 in healthy controls and in patients with PANS sampled during worsening flare and subsequent recovery. No significant differences were observed.

**Supplementary Figure S12.**
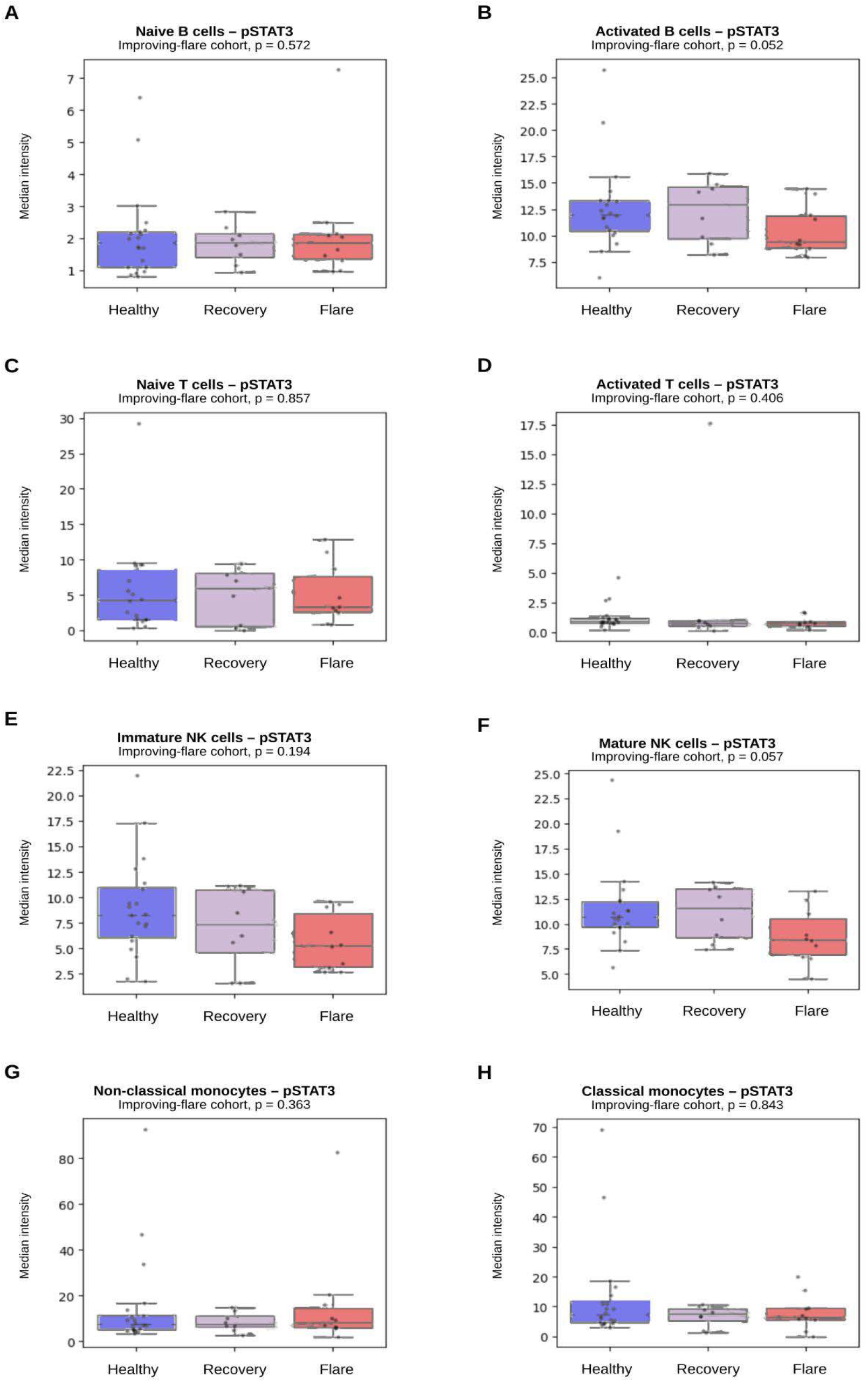
pSTAT3 signaling in the improving-flare cohort. Median intensity of pSTAT3 in healthy controls and in patients with PANS sampled during improving flare and subsequent recovery. No significant differences were observed.

**Supplementary Figure S13.**
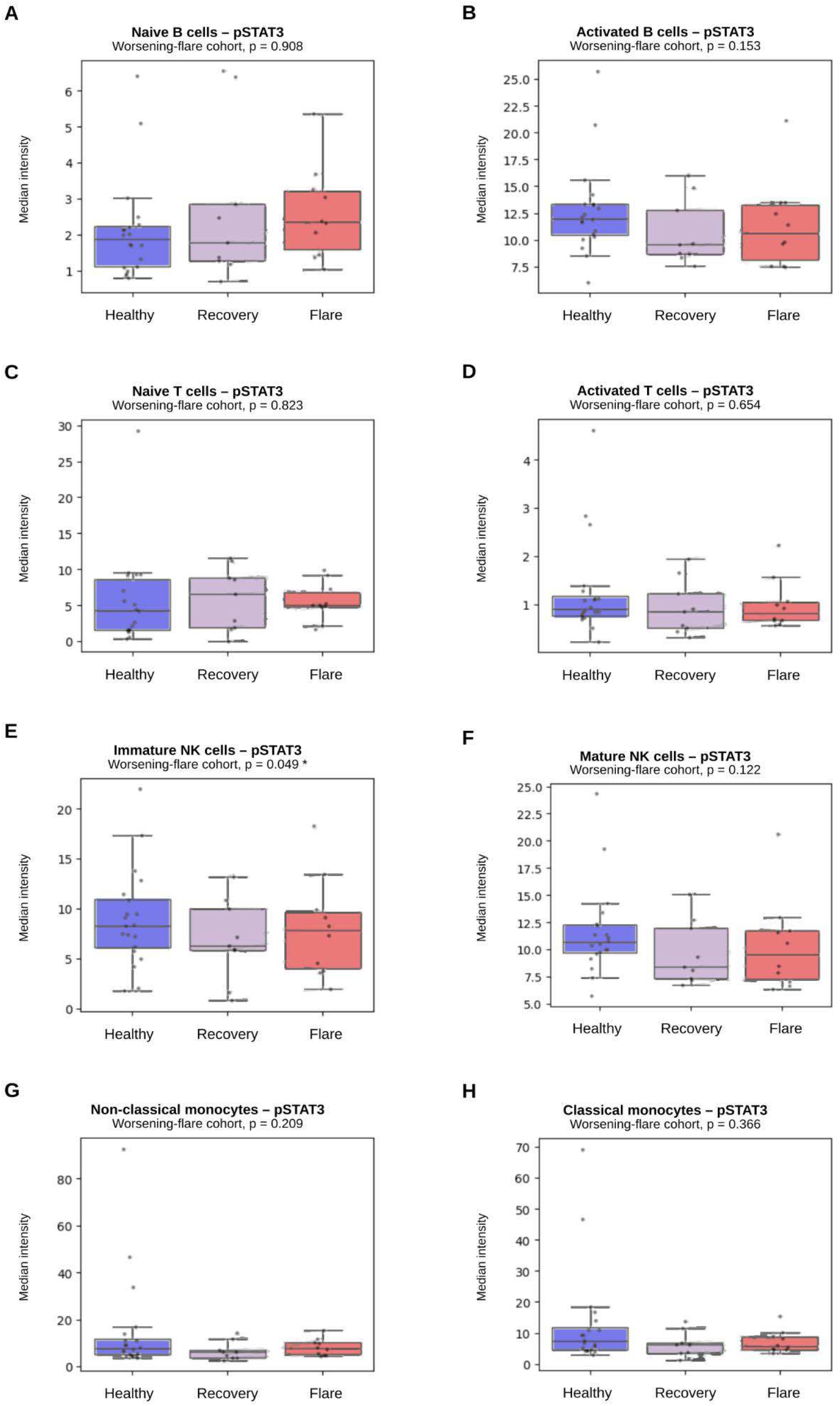
pSTAT3 signaling in the worsening-flare cohort. Median intensity of pSTAT3 in healthy controls and in patients with PANS sampled during worsening flare and subsequent recovery. A nominal difference was observed in immature NK cells (p = 0.049)

**Supplementary Figure S14.**
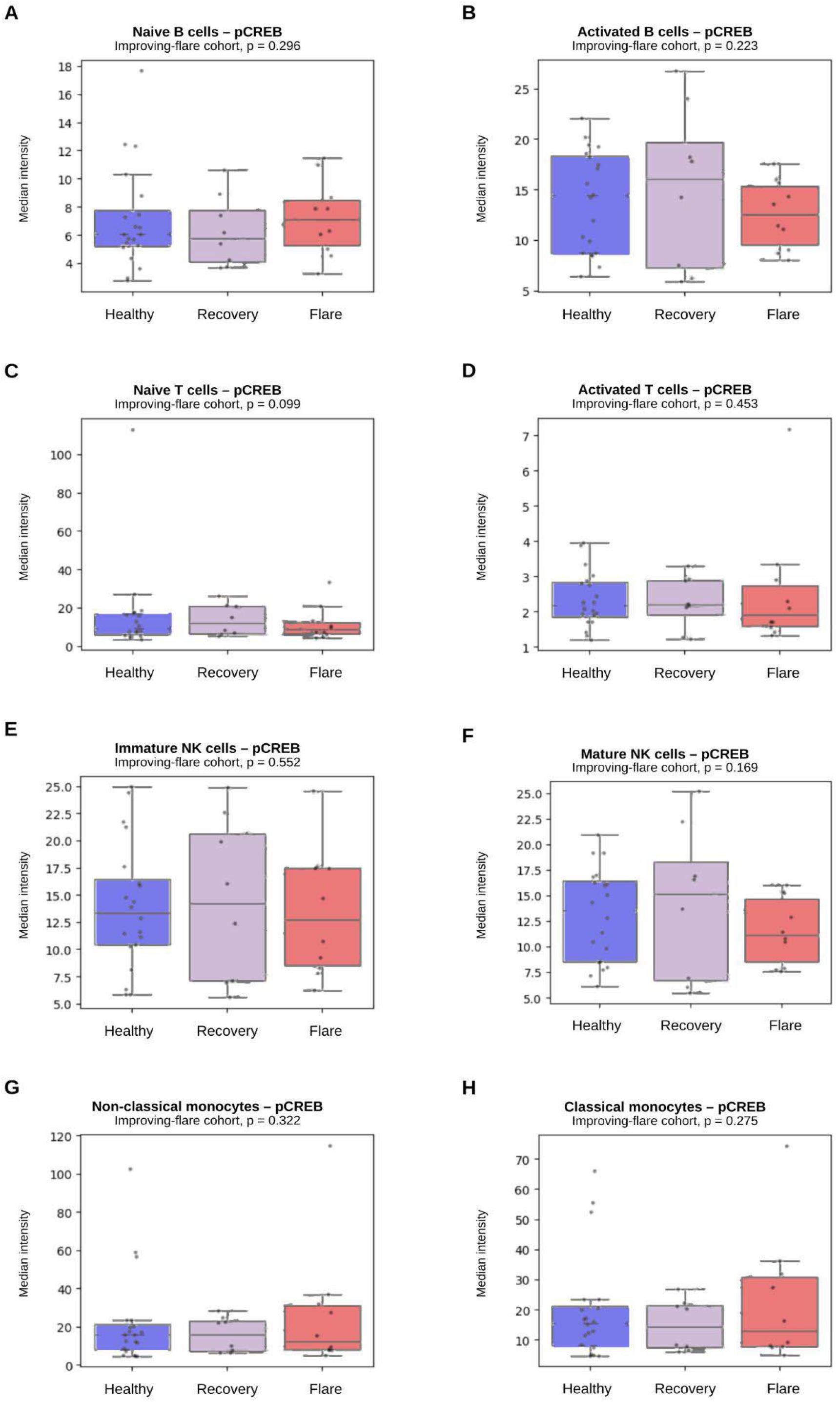
pCREB signaling in the improving-flare cohort. Median intensity of pCREB in healthy controls and in patients with PANS sampled during improving flare and subsequent recovery. No significant differences were observed.

**Supplementary Figure S15.**
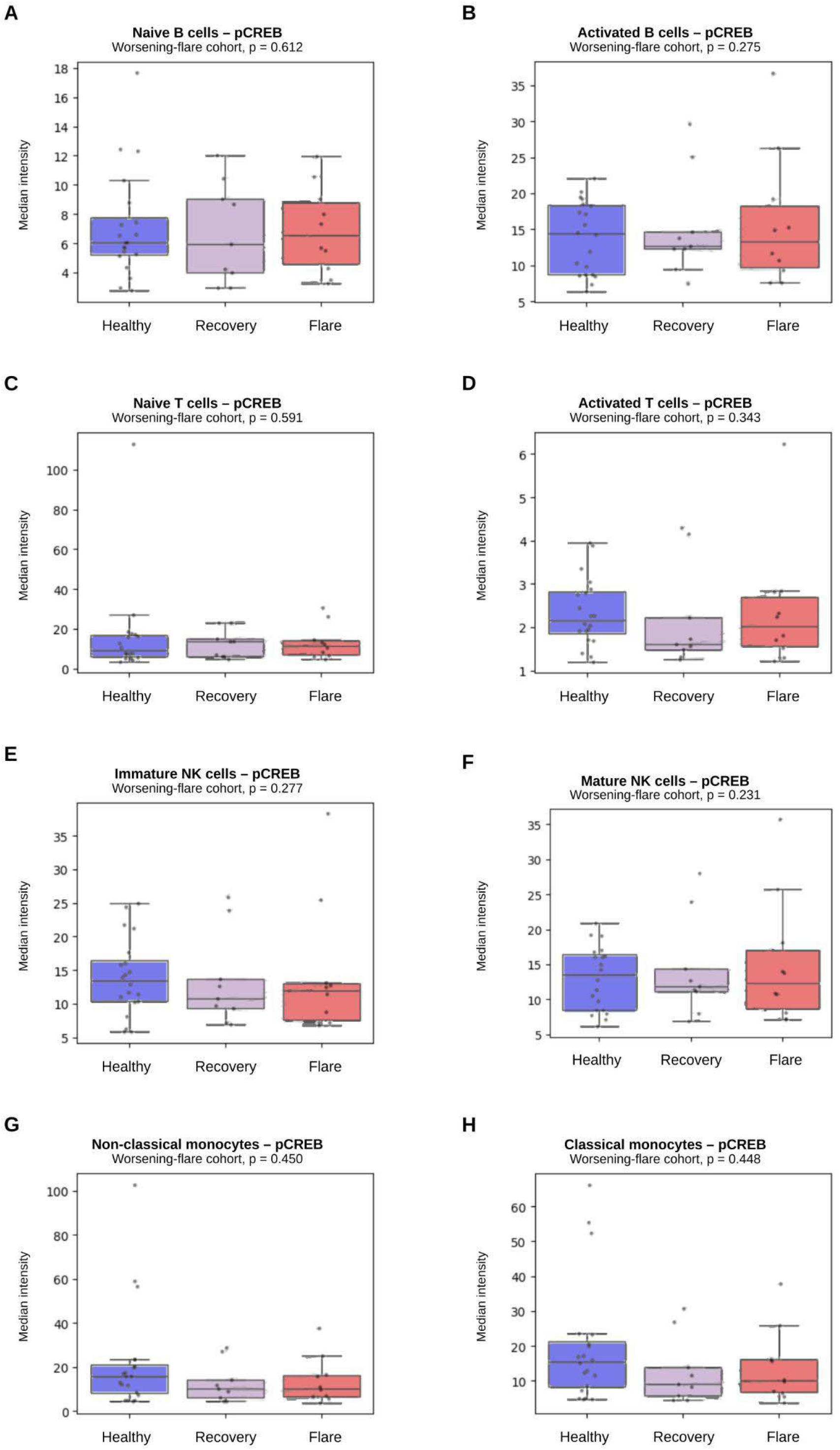
pCREB signaling in the worsening-flare cohort. Median intensity of pCREB in healthy controls and in patients with PANS sampled during worsening flare and subsequent recovery. No significant differences were observed.

**Supplementary Figure S16.**
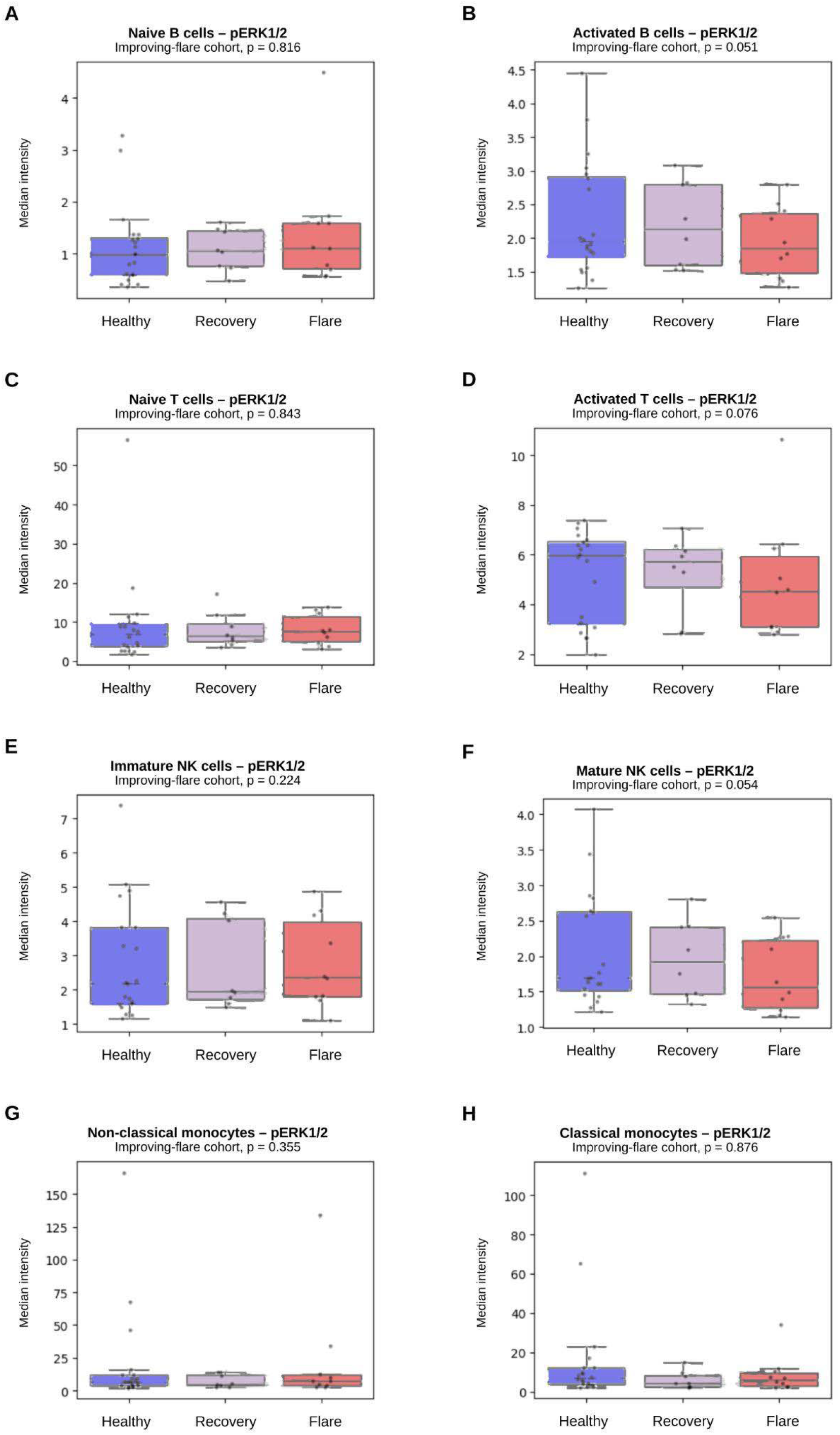
pERK1/2 signaling in the improving-flare cohort. Median intensity of pERK1/2 in healthy controls and in patients with PANS sampled during improving flare and subsequent recovery. No significant differences were observed.

**Supplementary Figure S17.**
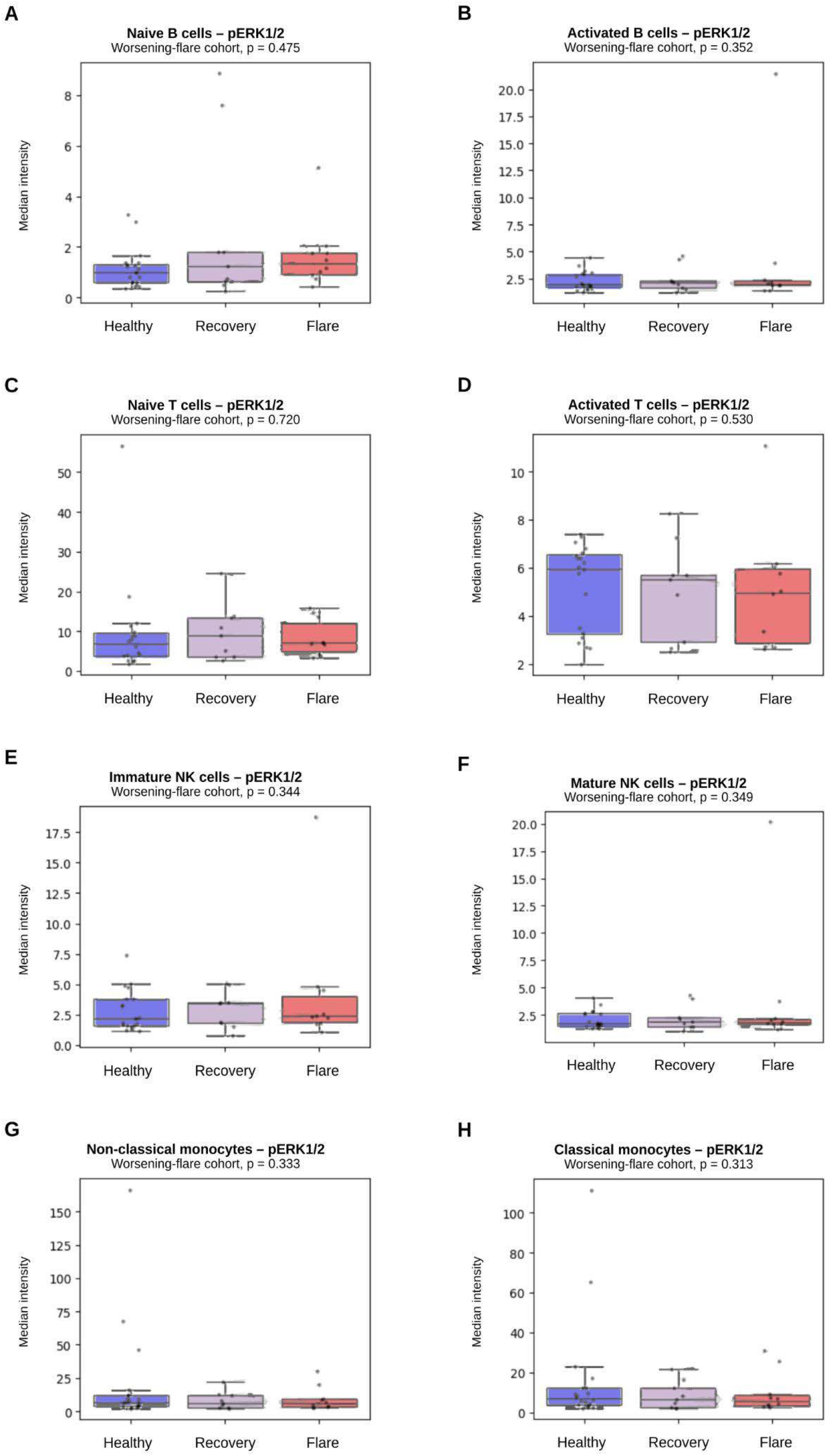
pERK1/2 signaling in the worsening-flare cohort. Median intensity of pERK1/2 in healthy controls and in patients with PANS sampled during worsening flare and subsequent recovery. No significant differences were observed.

**Supplementary Figure S18.**
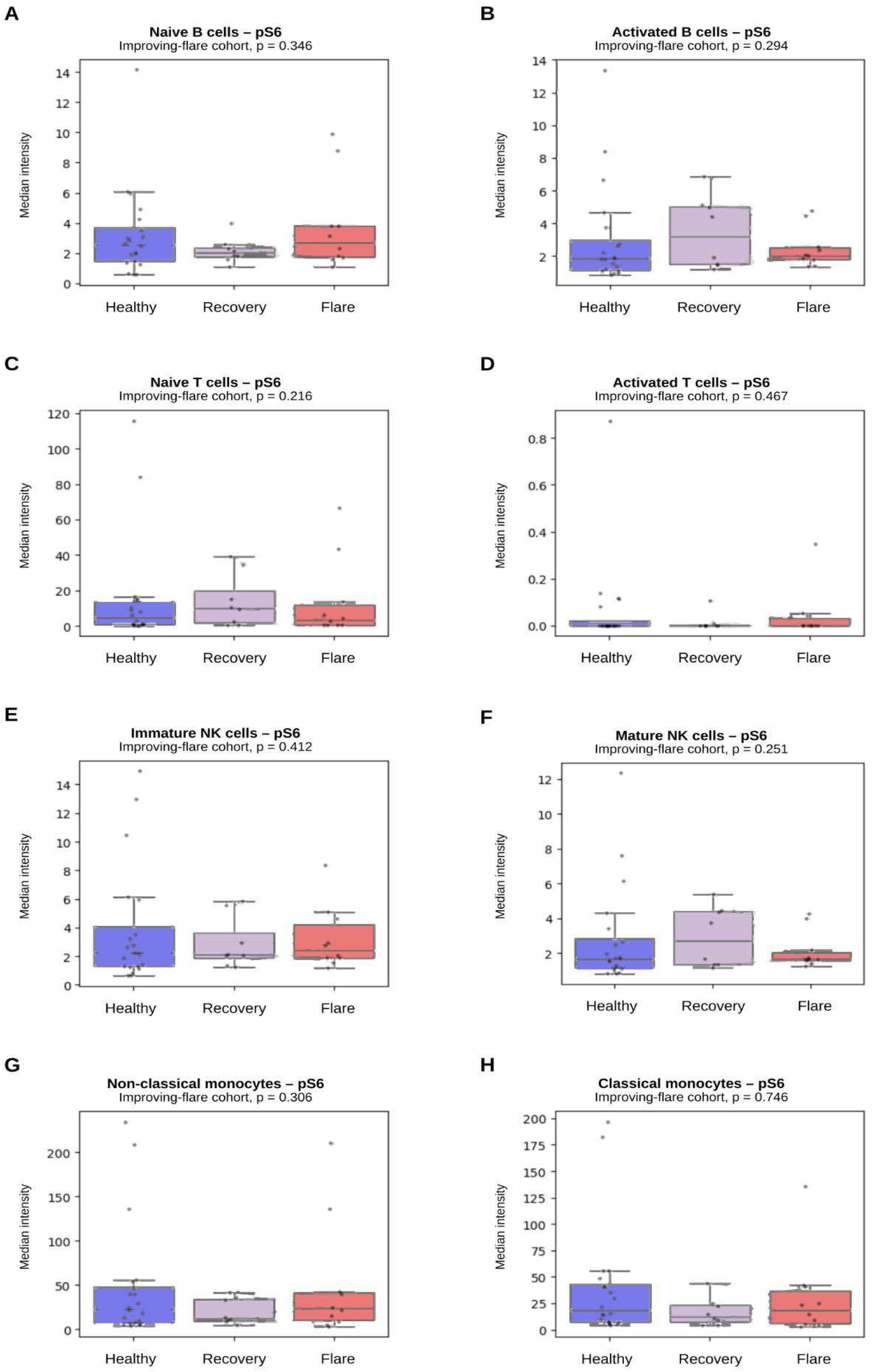
pS6 (mTOR) signaling in the improving-flare cohort. Median intensity of pS6 in healthy controls and in patients with PANS sampled during improving flare and subsequent recovery. No significant differences were observed.

**Supplementary Figure S19.**
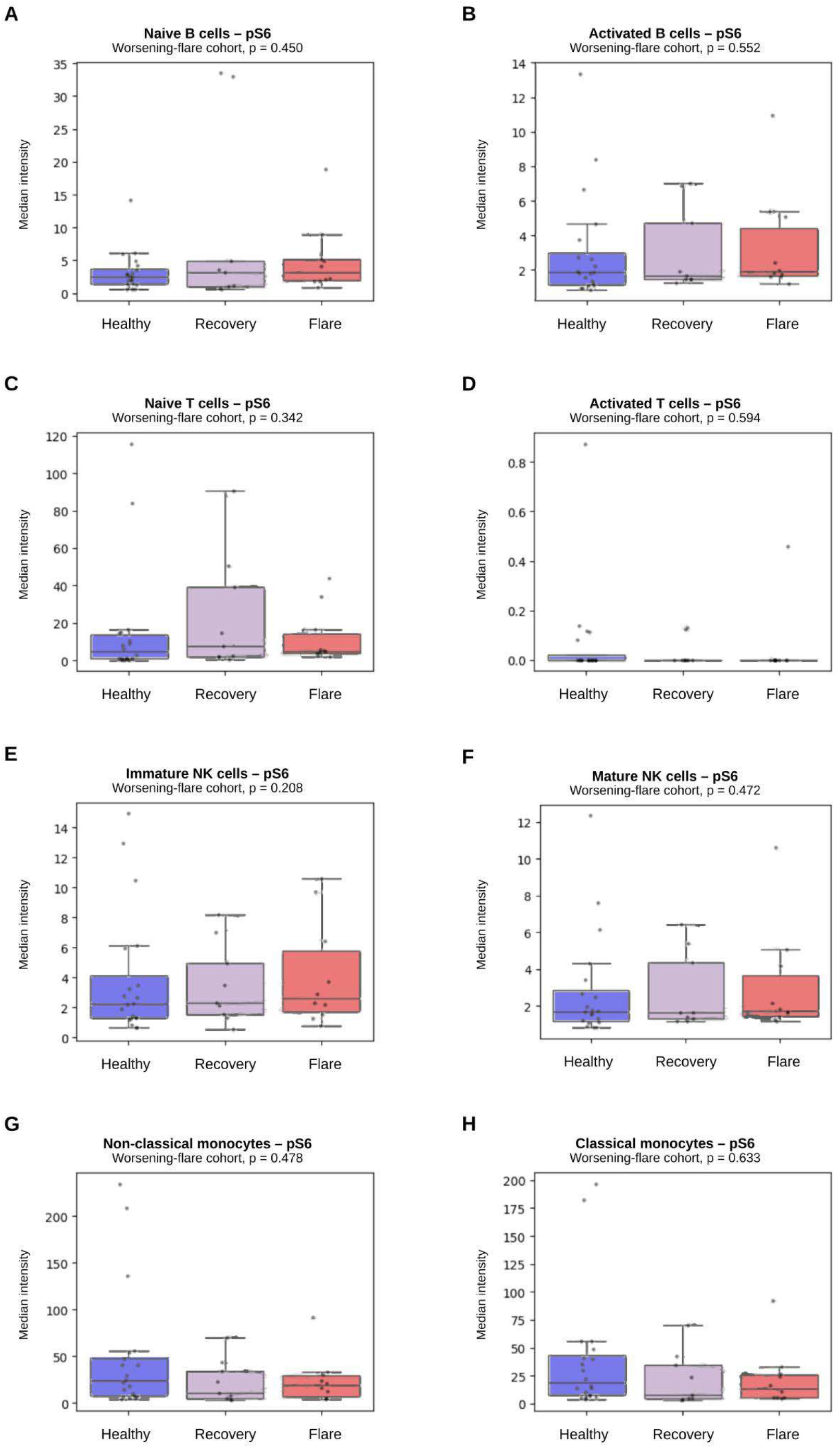
pS6 (mTOR) signaling in the worsening-flare cohort. Median intensity of pS6 in healthy controls and in patients with PANS sampled during worsening flare and subsequent recovery. No significant differences were observed

**Supplementary Figure S20.**
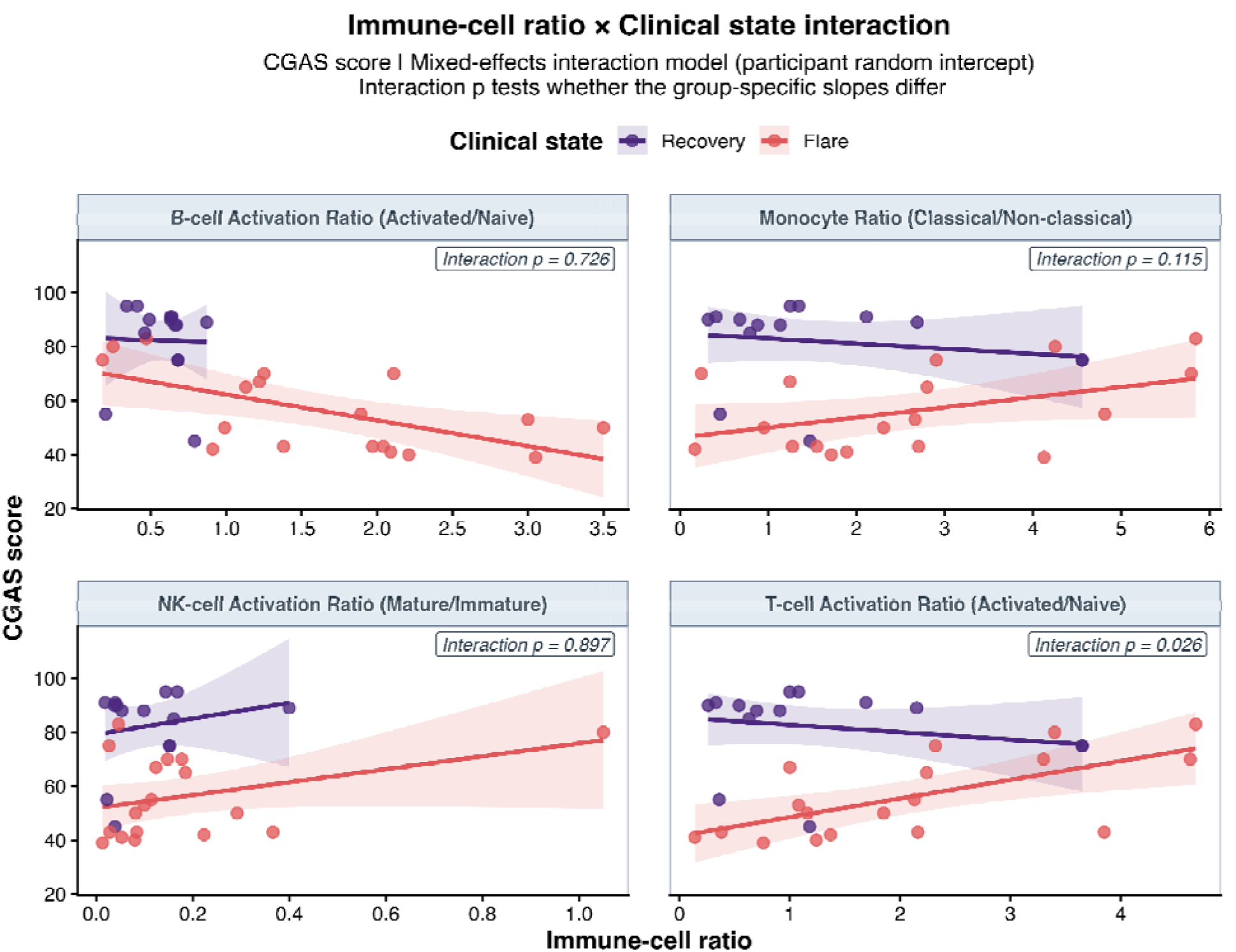
Interaction between immune-cell ratio and clinical state in relation to CGAS scores. Linear mixed-effects models included immune-cell ratio x clinical state interactions and participant-specific random intercepts. Points represent individual observations, lines indicate model-estimated associations, and shaded areas represent 95% confidence intervals.

**Supplementary Figure S21.**
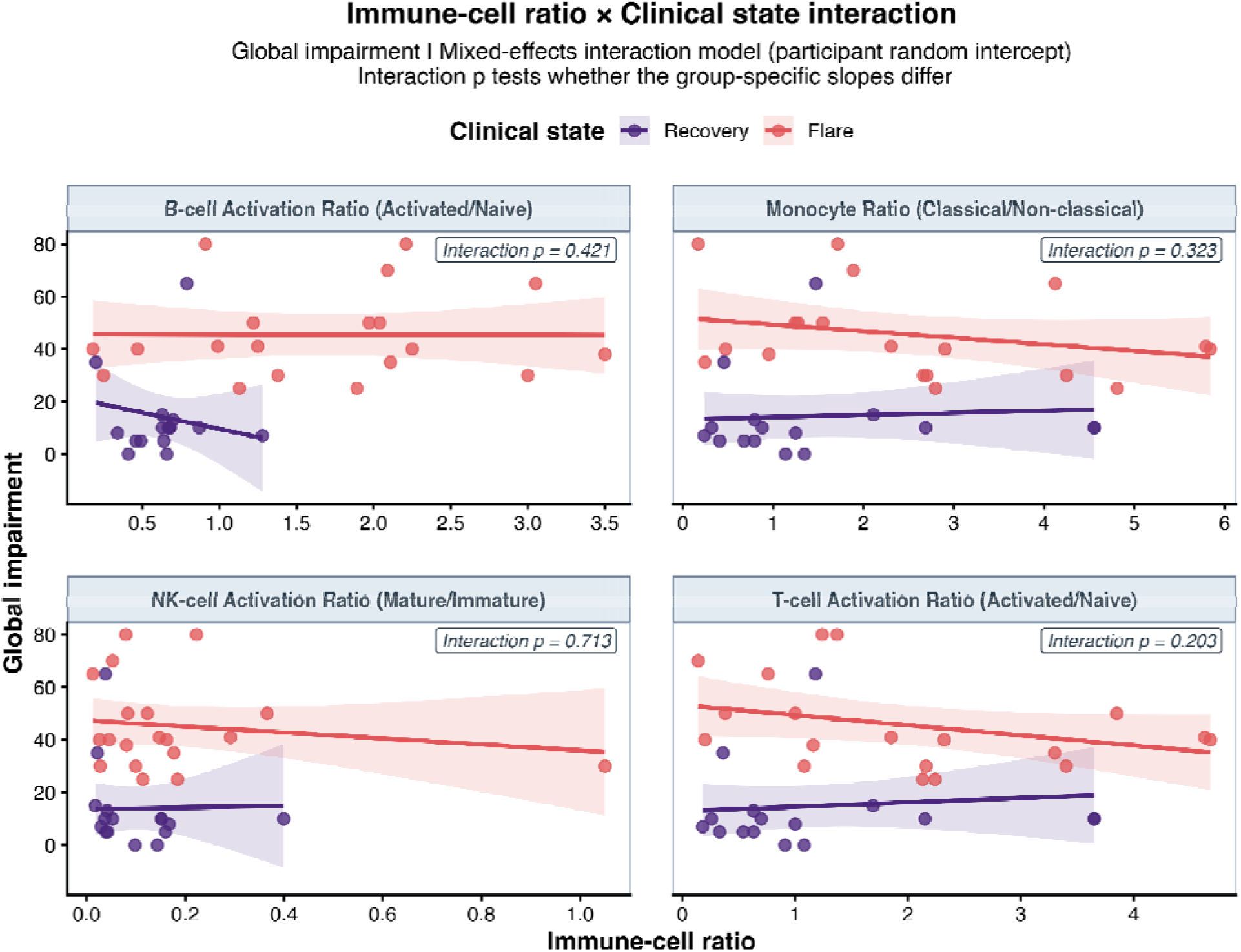
Interaction between immune-cell ratio and clinical state in relation to Global Impairment scores. Linear mixed-effects models included immune-cell ratio x clinical state interactions and participant-specific random intercepts. Points represent individual observations, lines indicate model-estimated associations, and shaded areas represent 95% confidence intervals.

**Supplementary Figure S22.**
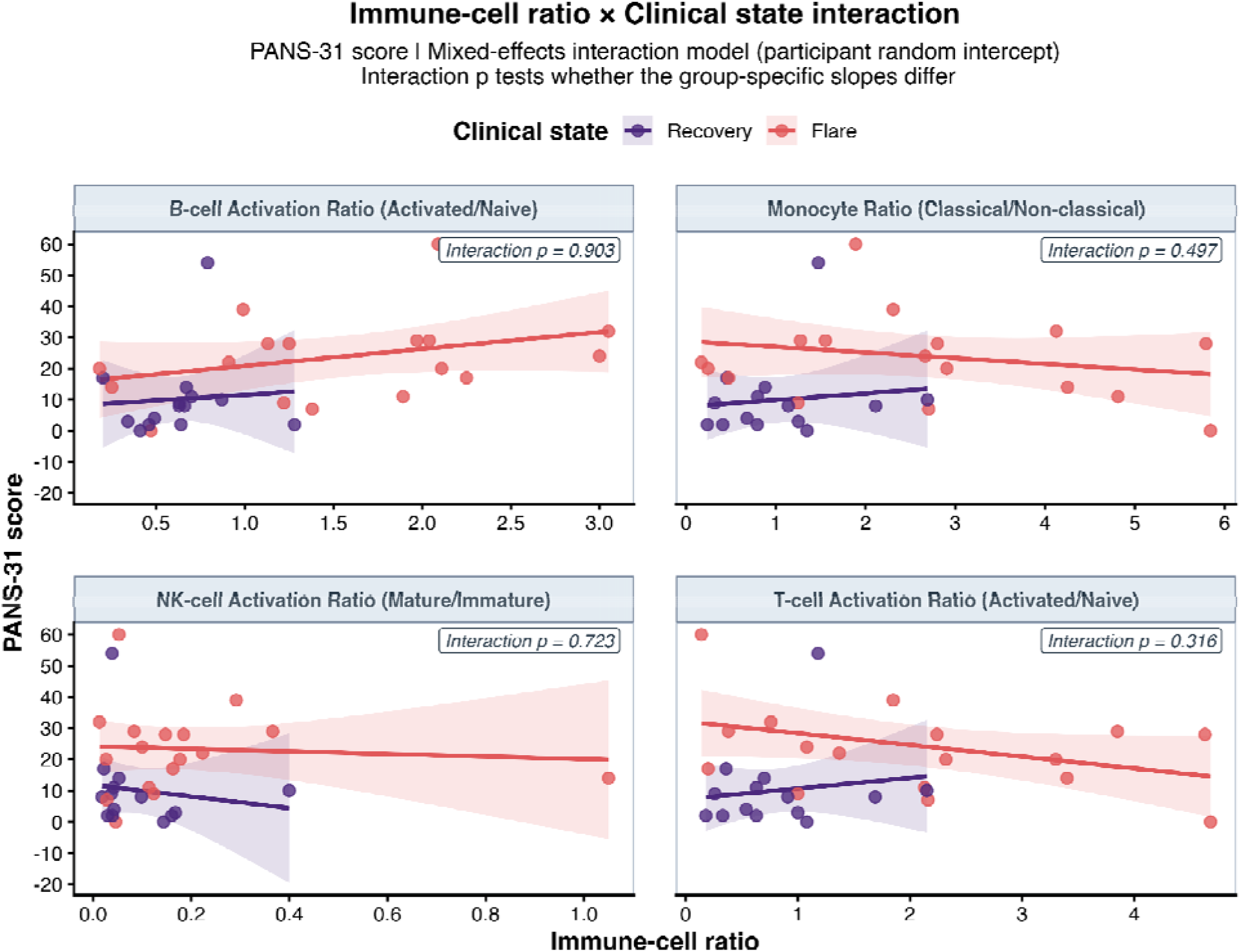
Interaction between immune-cell ratio and clinical state in relation to PANS-31 scores. Linear mixed-effects models included immune-cell ratio x clinical state interactions and participant-specific random intercepts. Points represent individual observations, lines indicate model-estimated associations, and shaded areas represent 95% confidence intervals.

**Supplementary Figure S23.**
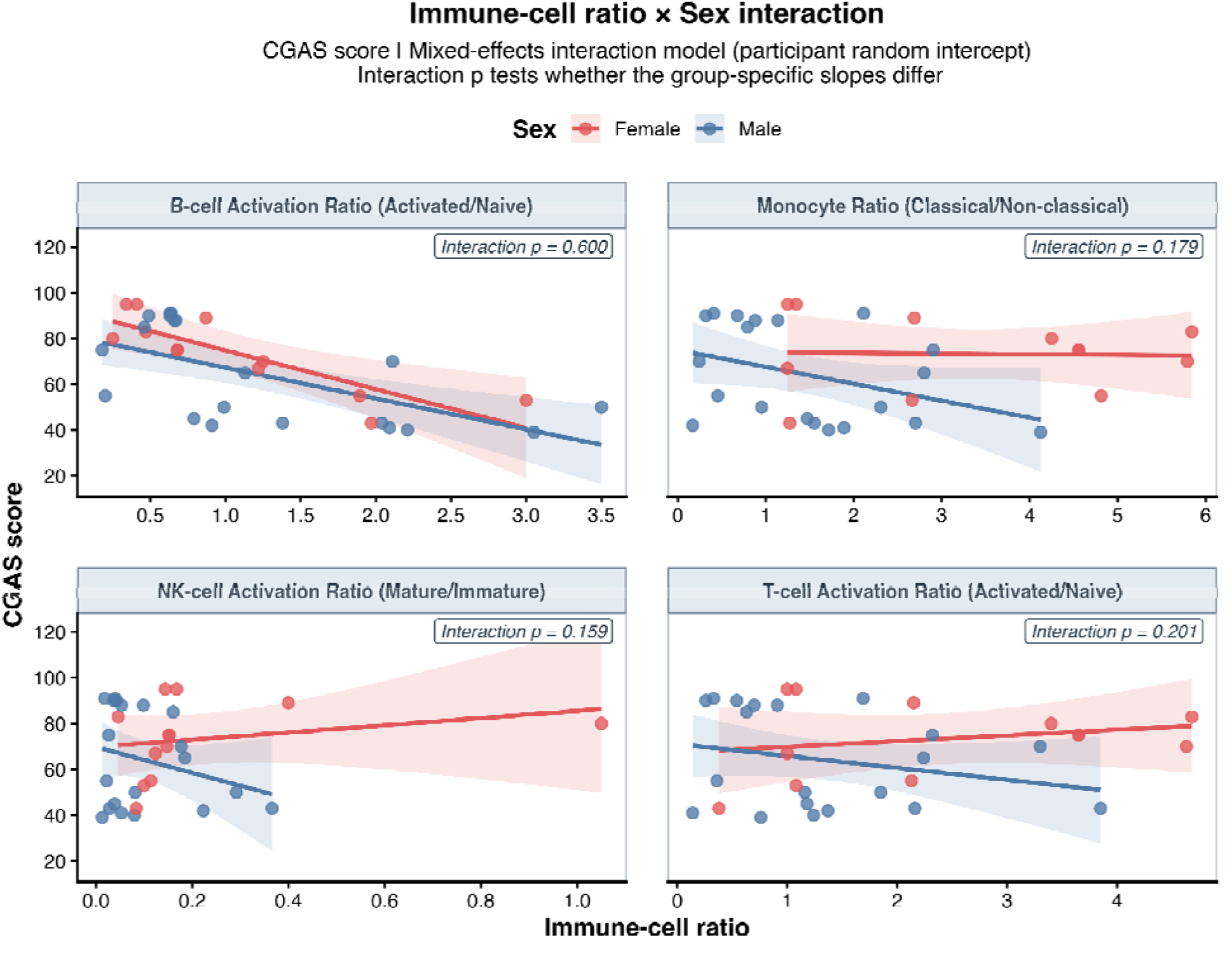
Interaction between immune-cell ratio and sex in relation to CGAS scores. Linear mixed-effects models included immune-cell ratio x sex interactions and participant-specific random intercepts. Points represent individual observations, lines indicate model-estimated associations, and shaded areas represent 95% confidence intervals.

**Supplementary Figure S24.**
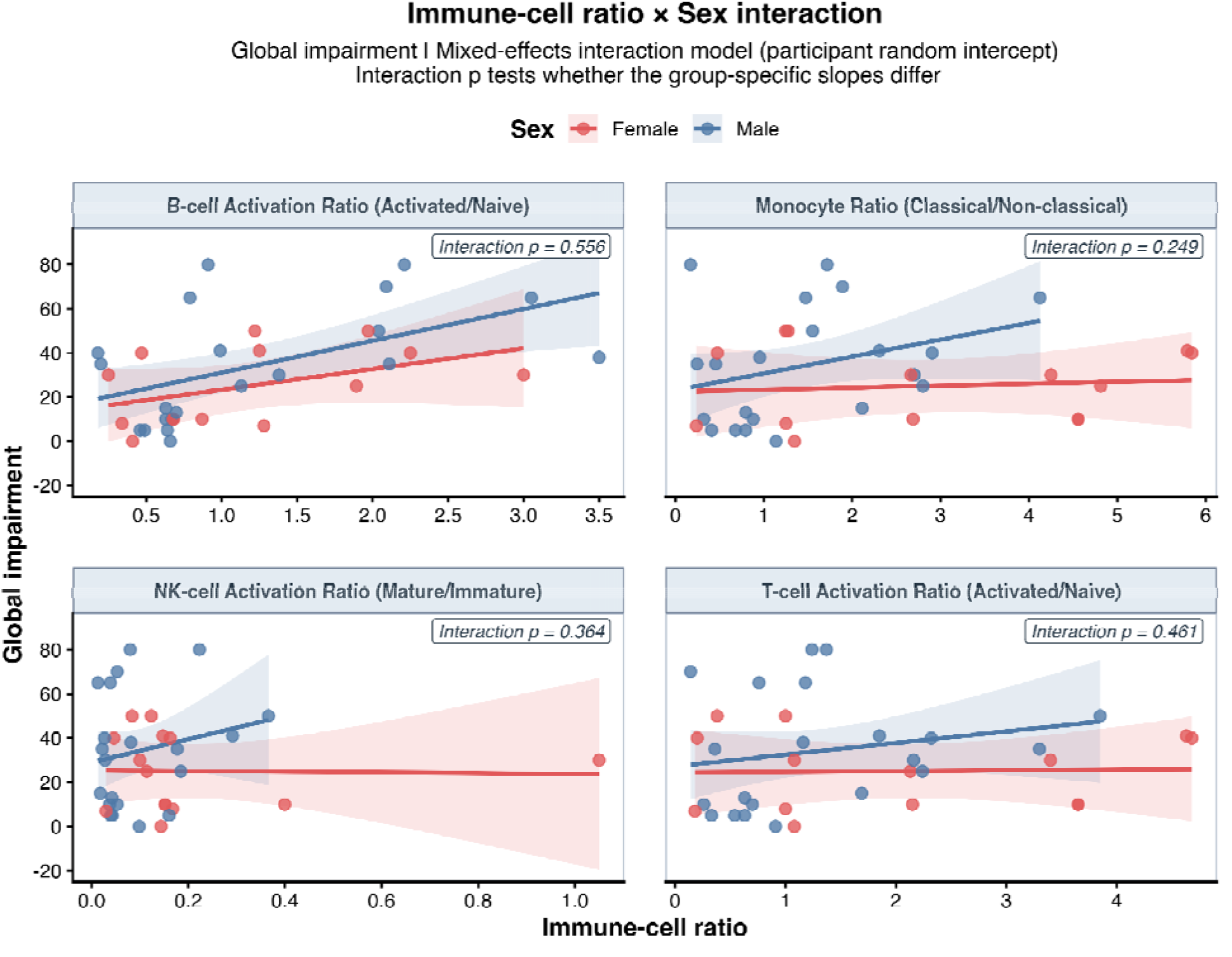
Interaction between immune-cell ratio and sex in relation to Global Impairment scores. Linear mixed-effects models included immune-cell ratio x sex interactions and participant-specific random intercepts. Points represent individual observations, lines indicate model-estimated associations, and shaded areas represent 95% confidence intervals.

**Supplementary Figure S25.**
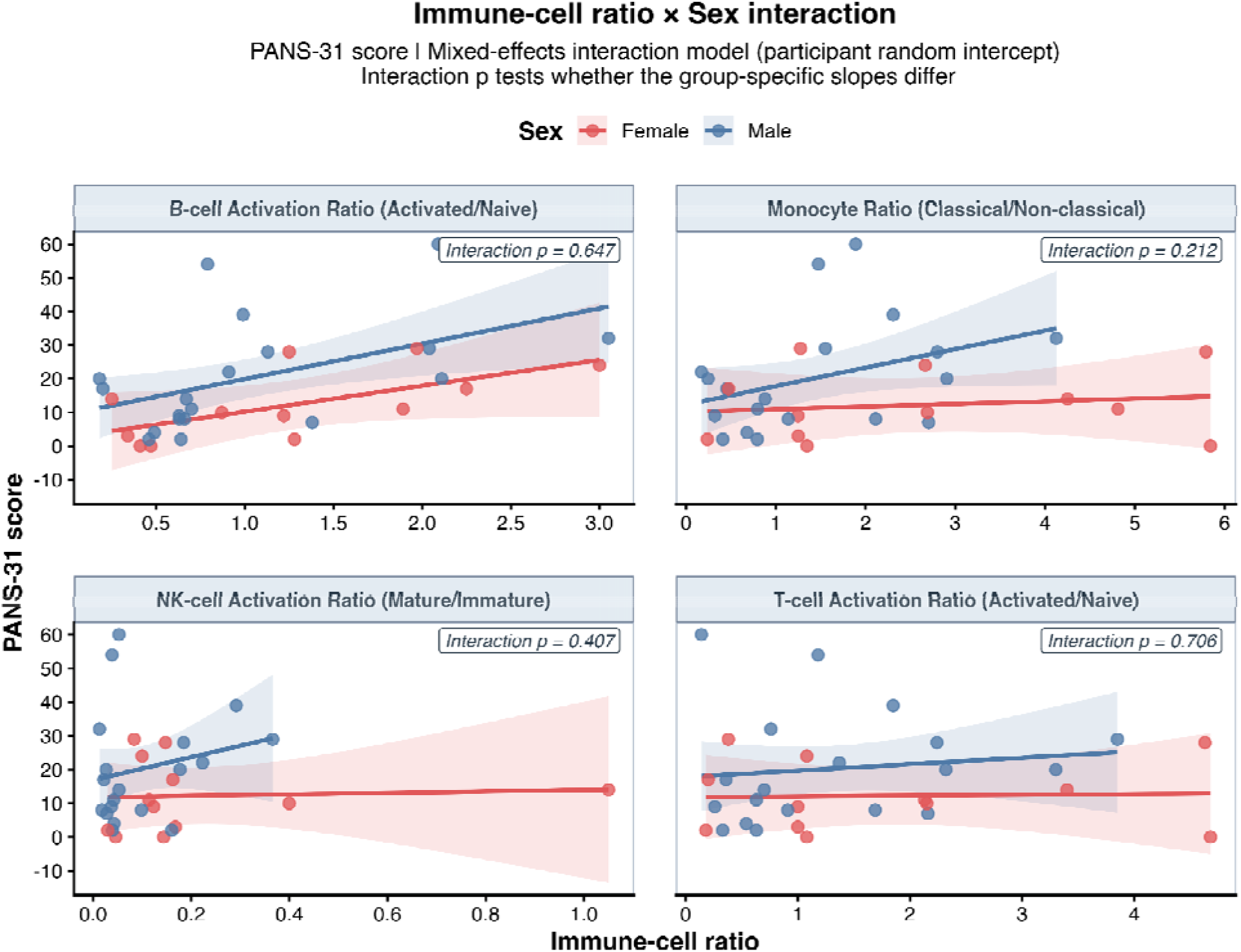
Interaction between immune-cell ratio and sex in relation to PANS-31 scores. Linear mixed-effects models included immune-cell ratio x sex interactions and participant-specific random intercepts. Points represent individual observations, lines indicate model-estimated associations, and shaded areas represent 95% confidence intervals.

**Supplementary Figure S26.**
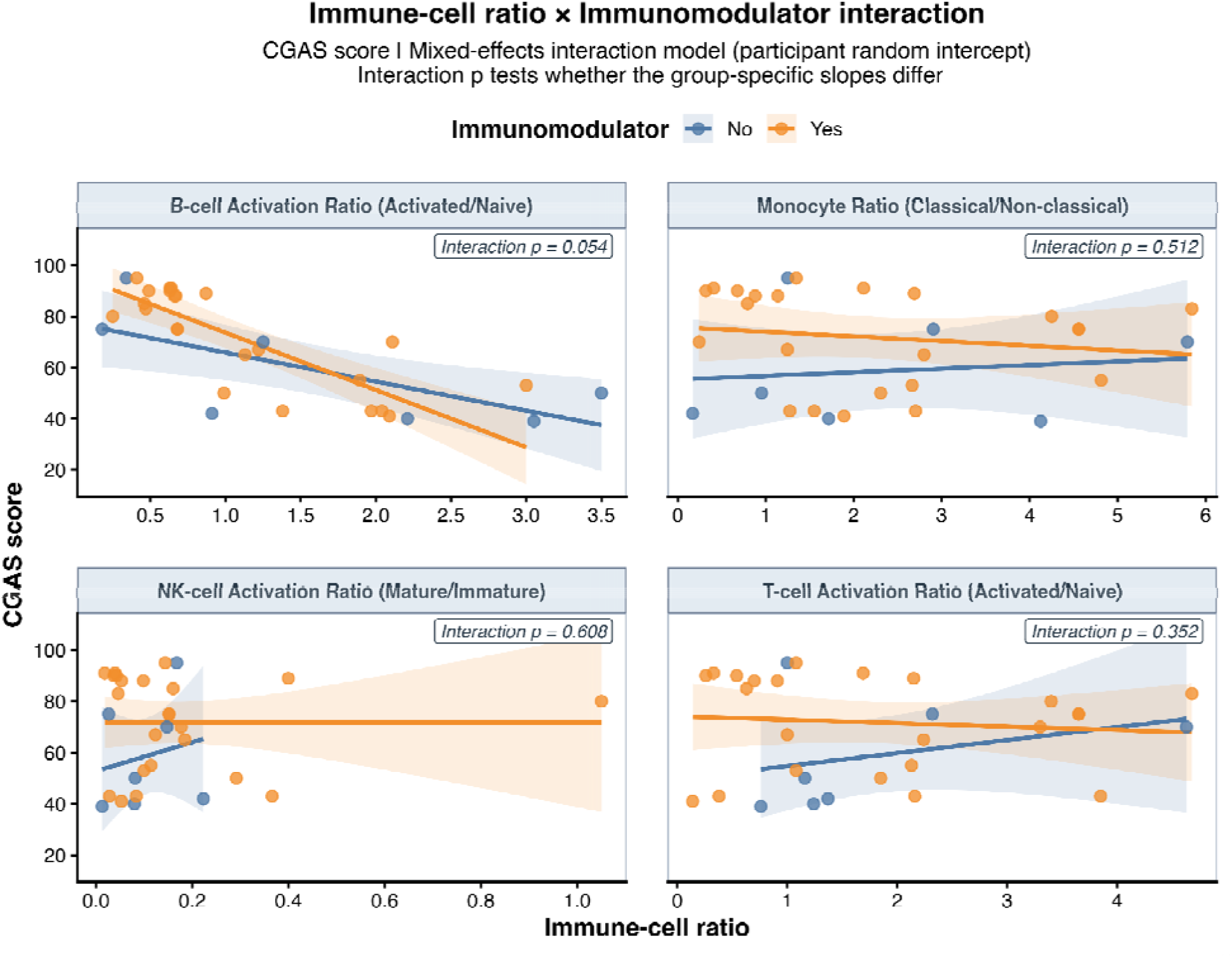
Interaction between immune-cell ratio and immunomodulator use in relation to CGAS scores. Linear mixed-effects models included immune-cell ratio x immunomodulator use interactions and participant-specific random intercepts. Points represent individual observations, lines indicate model-estimated associations, and shaded areas represent 95% confidence intervals.

**Supplementary Figure S27.**
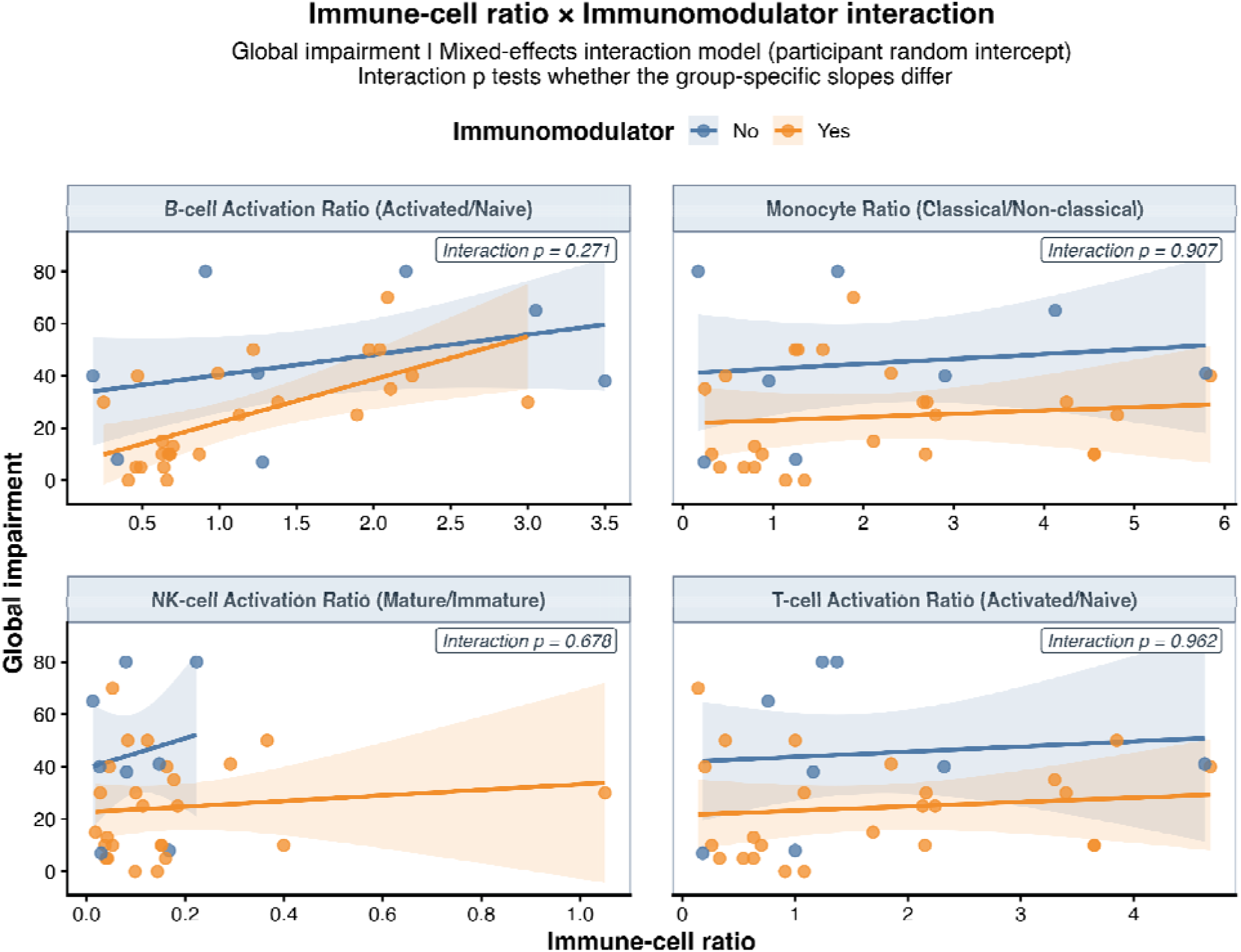
Interaction between immune-cell ratio and immunomodulator use in relation to Global Impairment scores. Linear mixed-effects models included immune-cell ratio x immunomodulator use interactions and participant-specific random intercepts. Points represent individual observations, lines indicate model-estimated associations, and shaded areas represent 95% confidence intervals.

**Supplementary Figure S28.**
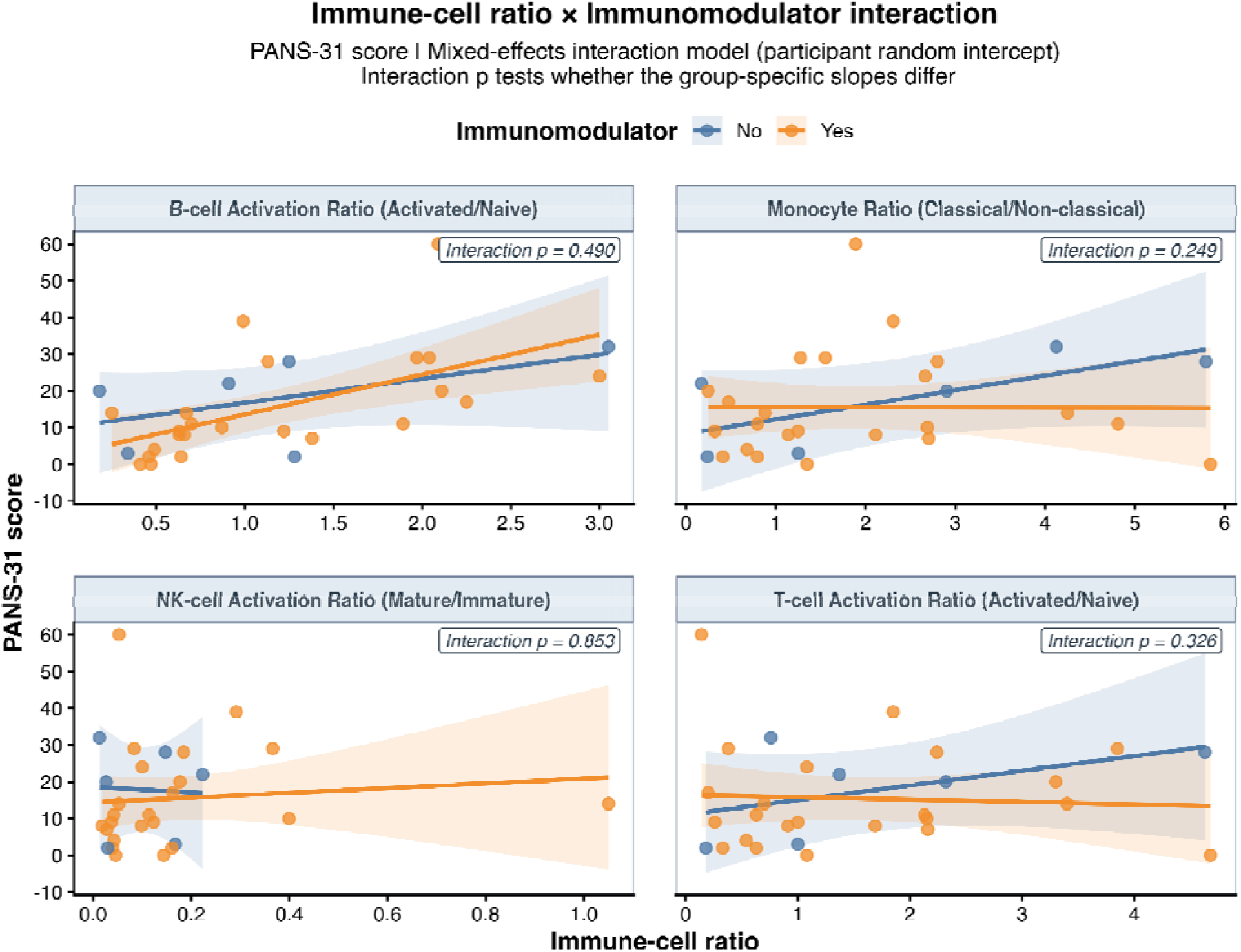
Interaction between immune-cell ratio and immunomodulator use in relation to PANS-31 scores. Linear mixed-effects models included immune-cell ratio x immunomodulator use interactions and participant-specific random intercepts. Points represent individual observations, lines indicate model-estimated associations, and shaded areas represent 95% confidence intervals.

## Notes

### Competing Interest Statement

The authors have declared no competing interest.

## References

1. Swedo, E., Leckman, J. & Rose, N. From research subgroup to clinical syndrome: Modifying the PANDAS criteria to describe PANS (pediatric acute-onset neuropsychiatric. Pediatr. Ther. https://api.semanticscholar.org/CorpusID:33224646} (2012).

2. Swedo, S. E. et al. Pediatric autoimmune neuropsychiatric disorders associated with streptococcal infections: clinical description of the first 50 cases. Am. J. Psychiatry 155, 264–271 (1998).

3. Carapetis, J. R., et al. Acute rheumatic fever and rheumatic heart disease. Nat. Rev. Dis. Primers 2, (2016).

4. Gewitz, M. H. et al. Revision of the Jones Criteria for the diagnosis of acute rheumatic fever in the era of Doppler echocardiography: a scientific statement from the American Heart Association. Circulation 131, 1806–1818 (2015).

5. Asbahr, F. R. et al. Obsessive-compulsive and related symptoms in children and adolescents with rheumatic fever with and without chorea: A prospective 6-month study. Am. J. Psychiatry 155, 1122–1124 (1998).

6. Teixeira, A. L., Vasconcelos, L. P., Nunes, M. do C. P. & Singer, H. Sydenham’s chorea: from pathophysiology to therapeutics. Expert Rev. Neurother. 21, 913–922 (2021).

7. Thomas, T. et al. Evaluation, diagnosis, and treatment of Sydenham chorea: Consensus guidelines. Pediatrics 156, (2025).

8. Orsini, A. et al. Psychopathological impact in patients with history of rheumatic fever with or without Sydenham’s chorea: A multicenter prospective study. Int. J. Environ. Res. Public Health 19, 10586 (2022).

9. Vreeland, A. et al. Postinfectious inflammation, autoimmunity, and obsessive-compulsive disorder: Sydenham chorea, pediatric autoimmune neuropsychiatric disorder associated with streptococcal infection, and pediatric acute-onset neuropsychiatric disorder. Dev. Neurosci. 45, 361–374 (2023).

10. Williams, K. A. & Swedo, S. E. Post-infectious autoimmune disorders: Sydenham’s chorea, PANDAS and beyond. Brain Res. 1617, 144–154 (2015).

11. Kirvan, C. A., Swedo, S. E., Heuser, J. S. & Cunningham, M. W. Mimicry and autoantibody-mediated neuronal cell signaling in Sydenham chorea. Nat. Med. 9, 914–920 (2003).

12. Kirvan, C. A., Swedo, S. E., Kurahara, D. & Cunningham, M. W. Streptococcal mimicry and antibody-mediated cell signaling in the pathogenesis of Sydenham’s chorea. Autoimmunity 39, 21–29 (2006).

13. Kirvan, C. A., Cox, C. J., Swedo, S. E. & Cunningham, M. W. Tubulin is a neuronal target of autoantibodies in Sydenham’s chorea. J. Immunol. 178, 7412–7421 (2007).

14. Brimberg, L. et al. Behavioral, pharmacological, and immunological abnormalities after streptococcal exposure: A novel rat model of Sydenham chorea and related neuropsychiatric disorders. Neuropsychopharmacology 37, 2076–2087 (2012).

15. Cox, C. J. et al. Brain human monoclonal autoantibody from Sydenham chorea targets dopaminergic neurons in transgenic mice and signals dopamine D2 receptor: Implications in human disease. J. Immunol. 191, 5524–5541 (2013).

16. Chain, J. L. et al. Autoantibody biomarkers for basal ganglia encephalitis in Sydenham chorea and pediatric autoimmune neuropsychiatric disorder associated with streptococcal infections. Front. Psychiatry 11, (2020).

17. Hoffman, K. L., Hornig, M., Yaddanapudi, K., Jabado, O. & Lipkin, W. I. A Murine model for neuropsychiatric disorders associated with group A β-hemolytic streptococcal infection. J. Neurosci. 24, 1780–1791 (2004).

18. Yaddanapudi, K. et al. Passive transfer of streptococcus-induced antibodies reproduces behavioral disturbances in a mouse model of pediatric autoimmune neuropsychiatric disorders associated with streptococcal infection. Mol. Psychiatry 15, 712–726 (2010).

19. Lotan, D. et al. Behavioral and neural effects of intra-striatal infusion of anti-streptococcal antibodies in rats. Brain Behav. Immun. 38, 249–262 (2014).

20. Dale, R. C. et al. Antibodies to surface dopamine-2 receptor in autoimmune movement and psychiatric disorders. Brain 135, 3453–3468 (2012).

21. Singer, H. S. et al. Neuronal antibody biomarkers for Sydenham’s chorea identify a new group of children with chronic recurrent episodic acute exacerbations of tic and obsessive compulsive symptoms following a streptococcal infection. PLoS One 10, e0120499 (2015).

22. Hesselmark, E. & Bejerot, S. Biomarkers for diagnosis of Pediatric Acute Neuropsychiatric Syndrome (PANS) – Sensitivity and specificity of the Cunningham Panel. J. Neuroimmunol. 312, 31–37 (2017).

23. Xu, J. et al. Antibodies from children with PANDAS bind specifically to striatal cholinergic interneurons and alter their activity. Am. J. Psychiatry 178, 48–64 (2021).

24. Xu, J. et al. Elevated antibody binding to striatal cholinergic interneurons in patients with pediatric acute-onset neuropsychiatric syndrome. Brain Behav. Immun. 122, 241–255 (2024).

25. Giedd, J. N., Rapoport, J. L., Garvey, M. A., Perlmutter, S. & Swedo, S. E. MRI assessment of children with obsessive-compulsive disorder or tics associated with streptococcal infection. Am. J. Psychiatry 157, 281–283 (2000).

26. Kumar, A., Williams, M. T. & Chugani, H. T. Evaluation of basal ganglia and thalamic inflammation in children with pediatric autoimmune neuropsychiatric disorders associated with streptococcal infection and Tourette syndrome. J. Child Neurol. 30, 749–756 (2015).

27. Cabrera, B. et al. Neuroanatomical features and its usefulness in classification of patients with PANDAS. CNS Spectr. 24, 533–543 (2019).

28. Zheng, J. et al. Association of pediatric acute-onset neuropsychiatric syndrome with microstructural differences in brain regions detected via diffusion-weighted magnetic resonance imaging. *JAMA Netw*. Open 3, e204063 (2020).

29. Zebrack, J. E. et al. Neurological soft signs at presentation in patients with pediatric acute-onset neuropsychiatric syndrome. *JAMA Netw*. Open 8, e250314 (2025).

30. Gaughan, T. et al. Rapid eye movement sleep abnormalities in children with pediatric acute-onset neuropsychiatric syndrome (PANS). J. Clin. Sleep Med. 12, 1027–1032 (2016).

31. Santoro, J. D., Frankovich, J. & Bhargava, S. Continued presence of period limb movements during REM sleep in patients with chronic static pediatric acute-onset neuropsychiatric syndrome (PANS). J. Clin. Sleep Med. 14, 1187–1192 (2018).

32. Congiu, P. et al. REM sleep atonia in patients with pediatric acute-onset neuropsychiatric syndrome: implications for pathophysiology. J. Clin. Sleep Med. 21, 757–764 (2025).

33. Gagliano, A. et al. Artificial Neural Networks Analysis of polysomnographic and clinical features in Pediatric Acute-Onset Neuropsychiatric Syndrome (PANS): from sleep alteration to ‘Brain Fog’. Nat. Sci. Sleep 13, 1209–1224 (2021).

34. Frick, L. R. et al. Differential binding of antibodies in PANDAS patients to cholinergic interneurons in the striatum. Brain Behav. Immun. 69, 304–311 (2018).

35. Dileepan, T. et al. Group A Streptococcus intranasal infection promotes CNS infiltration by streptococcal-specific Th17 cells. J. Clin. Invest. 126, 303–317 (2015).

36. Platt, M. P. et al. Th17 lymphocytes drive vascular and neuronal deficits in a mouse model of postinfectious autoimmune encephalitis. Proc. Natl. Acad. Sci. U. S. A. 117, 6708–6716 (2020).

37. Wayne, C. R. et al. Th17 effector cytokines induce shared and distinct microglial and endothelial cell responses in a mouse model for post-streptococcal encephalitis. Nat. Commun. 17, 9658 (2026).

38. Kim, M. L. et al. Dysregulated IL-1β-GM-CSF axis in acute rheumatic fever that is limited by hydroxychloroquine. Circulation 138, 2648–2661 (2018).

39. Middleton, F. M., McGregor, R., Webb, R. H., Wilson, N. J. & Moreland, N. J. Cytokine imbalance in acute rheumatic fever and rheumatic heart disease: Mechanisms and therapeutic implications. Autoimmun. Rev. 21, 103209 (2022).

40. Middleton, F. M. et al. CXCR3 is associated with T-cell-induced heart damage in acute rheumatic fever. Nat. Commun. 17, (2026).

41. Ma, M. et al. Development of autoimmune diseases among children with pediatric acute-onset neuropsychiatric syndrome. *JAMA Netw*. Open 7, e2421688 (2024).

42. Ma, M. et al. Arthritis in children with psychiatric deteriorations: A case series. Dev. Neurosci. 45, 325–334 (2023).

43. Gewitz, M. H. et al. Revision of the Jones criteria for the diagnosis of acute Rheumatic Fever in the era of Doppler echocardiography. Circulation 131, 1806–1818 (2015).

44. Brown, K. et al. Pediatric acute-onset neuropsychiatric syndrome response to oral corticosteroid bursts: An observational study of patients in an academic community-based PANS clinic. J. Child Adolesc. Psychopharmacol. 27, 629–639 (2017).

45. Spartz, E. J. et al. Course of neuropsychiatric symptoms after introduction and removal of nonsteroidal anti-inflammatory drugs: A pediatric observational study. J. Child Adolesc. Psychopharmacol. 27, 652–659 (2017).

46. Uziel, Y. et al. The use of naproxen in the treatment of children with rheumatic fever. J. Pediatr. 137, 269–271 (2000).

47. Hashkes, P. J. et al. Naproxen as an alternative to aspirin for the treatment of arthritis of rheumatic fever: a randomized trial. J. Pediatr. 143, 399–401 (2003).

48. Paz, J. A., Silva, C. A. A. & Marques-Dias, M. J. Randomized double-blind study with prednisone in Sydenham’s chorea. Pediatr. Neurol. 34, 264–269 (2006).

49. Eyre, M. et al. Treatments and outcomes among patients with Sydenham chorea. *JAMA Netw*. Open 7, e246792 (2024).

50. Yin, X. et al. Interferon-λ-neutralizing autoantibodies and common autoimmune disease autoantibodies in pediatric acute-onset neuropsychiatric syndrome. Front. Immunol. 17, (2026).

51. McGregor, R. et al. Mapping autoantibodies in children with acute rheumatic fever. Front. Immunol. 12, (2021).

52. McGregor, R., et al. PhIP-Seq uncovers marked heterogeneity in acute rheumatic fever autoantibodies. JCI Insight 11, (2026).

53. Rahman, S. S. et al. Sex-associated and disease state-dependent monocyte polarization and CNS-trafficking phenotypes in pediatric acute-onset neuropsychiatric syndrome (PANS). J. Neuroinflammation 22, 273 (2025).

54. Melamed, I. et al. IVIG response in pediatric acute-onset neuropsychiatric syndrome correlates with reduction in pro-inflammatory monocytes and neuropsychiatric measures. Front. Immunol. 15, 1383973 (2024).

55. Sigal, N. & Maecker, H. T. Mass cytometry assessment of cell phenotypes and signaling states in human whole blood. in Methods in Molecular Biology 113–128 (Springer US, New York, NY, 2022).

56. Fernandez, R. & Maecker, H. Cytokine-Stimulated Phosphoflow of Whole Blood Using CyTOF Mass Cytometry. Bio Protoc. 5, (2015).

57. Shaffer, D. A children’s global assessment scale (CGAS). Arch. Gen. Psychiatry 40, 1228 (1983).

58. Leibold, C., Thienemann, M., Farhadian, B., Willett, T. & Frankovich, J. Psychometric properties of the Pediatric Acute-onset Neuropsychiatric Syndrome Global Impairment Score in children and adolescents with Pediatric Acute-onset Neuropsychiatric Syndrome. J. Child Adolesc. Psychopharmacol. 29, 41–49 (2019).

59. Bernstein, G. A. et al. Psychometric properties of the PANS 31-Item Symptom Rating Scale. J. Child Adolesc. Psychopharmacol. 34, 157–162 (2024).

60. Mahendra, A., Misra, R. & Aggarwal, A. Th1 and Th17 predominance in the enthesitis-related arthritis form of juvenile idiopathic arthritis. J. Rheumatol. 36, 1730–1736 (2009).

61. Gaur, P., Misra, R. & Aggarwal, A. Natural killer cell and gamma delta T cell alterations in enthesitis related arthritis category of juvenile idiopathic arthritis. Clin. Immunol. 161, 163–169 (2015).

62. Gaur, P., Myles, A., Misra, R. & Aggarwal, A. Intermediate monocytes are increased in enthesitis-related arthritis, a category of juvenile idiopathic arthritis. Clin. Exp. Immunol. 187, 234–241 (2017).

63. Manion, K. et al. Different immunologic profiles are associated with distinct clinical phenotypes in longitudinally observed patients with systemic lupus erythematosus. Arthritis Rheumatol. 76, 726–738 (2024).

64. Dorner, B. G. et al. Selective expression of the chemokine receptor XCR1 on cross-presenting dendritic cells determines cooperation with CD8+ T cells. Immunity 31, 823–833 (2009).

65. Marsh-Wakefield, F. et al. Peripheral B-cell dysregulation is associated with relapse after long-term quiescence in patients with multiple sclerosis. Immunol. Cell Biol. 100, 453–467 (2022).

66. Nashi, E., Wang, Y. & Diamond, B. The role of B cells in lupus pathogenesis. Int. J. Biochem. Cell Biol. 42, 543–550 (2010).

67. Wiendl, H. et al. Corrigendum: Blood biomarker dynamics in people with relapsing multiple sclerosis treated with cladribine tablets: results of the 2-year MAGNIFY-MS study. Front. Immunol. 16, (2025).

68. Bugatti, S., Vitolo, B., Caporali, R., Montecucco, C. & Manzo, A. B cells in rheumatoid arthritis: From pathogenic players to disease biomarkers. Biomed Res. Int. 2014, 1–14 (2014).

69. Edwards, J. C. W. et al. Efficacy of B-cell–targeted therapy with rituximab in patients with rheumatoid arthritis. N. Engl. J. Med. 350, 2572–2581 (2004).

70. Caporali, R., Caprioli, M., Bobbio-Pallavicini, F., Bugatti, S. & Montecucco, C. Long term treatment of rheumatoid arthritis with rituximab. Autoimmun. Rev. 8, 591–594 (2009).

71. Sabatino, J. J., Jr, Pröbstel, A.-K. & Zamvil, S. S. B cells in autoimmune and neurodegenerative central nervous system diseases. Nat. Rev. Neurosci. 20, 728–745 (2019).

72. Kim, K. et al. Therapeutic B-cell depletion reverses progression of Alzheimer’s disease. Nat. Commun. 12, 2185 (2021).

73. Engler-Chiurazzi, E. B cells and the stressed brain: emerging evidence of neuroimmune interactions in the context of psychosocial stress and major depression. Front. Cell. Neurosci. 18, 1360242 (2024).

74. Han, V. X. et al. Epigenetic, ribosomal, and immune dysregulation in paediatric acute-onset neuropsychiatric syndrome. Mol. Psychiatry 30, 5389–5404 (2025).

75. Chang, Kiki et al. Clinical evaluation of youth with pediatric acute-onset neuropsychiatric syndrome (PANS): recommendations from the 2013 PANS Consensus Conference. Journal of child and adolescent psychopharmacology vol. 25,1 (2015).

76. Masterson, Erin E et al. Defining Clinical Course of Patients Evaluated for Pediatric Acute-Onset Neuropsychiatric Syndrome: Phenotypic Classification Based on 10 Years of Clinical Data. Developmental neuroscience vol. 47,4 (2025).

